# Early immune responses have long-term associations with clinical, virologic, and immunologic outcomes in patients with COVID-19

**DOI:** 10.1101/2021.08.27.21262687

**Authors:** Zicheng Hu, Kattria van der Ploeg, Saborni Chakraborty, Prabhu Arunachalam, Diego Martinez Mori, Karen B. Jacobson, Hector Bonilla, Julie Parsonnet, Jason Andrews, Haley Hedlin, Lauren de la Parte, Kathleen Dantzler, Maureen Ty, Gene S Tan, Catherine A. Blish, Saki Takahashi, Isabel Rodriguez-Barraquer, Bryan Greenhouse, Atul J. Butte, Upinder Singh, Bali Pulendran, Taia T. Wang, Prasanna Jagannathan

## Abstract

The great majority of SARS-CoV-2 infections are mild and uncomplicated, but some individuals with initially mild COVID-19 progressively develop more severe symptoms. Furthermore, mild to moderate infections are an important contributor to ongoing transmission. There remains a critical need to identify host immune biomarkers predictive of clinical and virologic outcomes in SARS-CoV-2-infected patients. Leveraging longitudinal samples and data from a clinical trial of Peginterferon Lambda for treatment of SARS-CoV-2 infected outpatients, we used host proteomics and transcriptomics to characterize the trajectory of the immune response in COVID-19 patients within the first 2 weeks of symptom onset. We define early immune signatures, including plasma levels of RIG-I and the CCR2 ligands (MCP1, MCP2 and MCP3), associated with control of oropharyngeal viral load, the degree of symptom severity, and immune memory (including SARS-CoV-2-specific T cell responses and spike (S) protein-binding IgG levels). We found that individuals receiving BNT162b2 (Pfizer–BioNTech) vaccine had similar early immune trajectories to those observed in this natural infection cohort, including the induction of both inflammatory cytokines (e.g. MCP1) and negative immune regulators (e.g. TWEAK). Finally, we demonstrate that machine learning models using 8-10 plasma protein markers measured early within the course of infection are able to accurately predict symptom severity, T cell memory, and the antibody response post-infection.

---

Although the clinical course of COVID-19 exhibits considerable heterogeneity, the great majority of severe acute respiratory syndrome-related coronavirus 2 (SARS-CoV-2) infections are uncomplicated with mild symptoms throughout the course of the illness. However, some initially mild infections progress to more severe and/or prolonged symptoms as well as sustained disability^1^. Moreover, mild infections are an important contributor to ongoing transmission. An improved understanding of host determinants of clinical, virologic, and immunologic outcomes of SARS-CoV-2 infection is essential to addressing the unmet clinical need for novel therapies for COVID-19.

The early host response to acute SARS-CoV-2 infection likely plays a critical role in determining disease outcome and generation of virus-specific memory immune responses. Nucleic acid pattern recognition receptors (PRRs) mediate the early detection and host response to viral infections, with RNA virus recognition thought to occur mainly in the endosomal and/or cytosolic compartment by two different PRRs: Toll-like receptors (TLRs) and RIG-I-Like Receptors (RLRs). Viral recognition by TLRs and RLRs typically triggers a signaling cascade leading to induction of pro-inflammatory cytokines and type I and type III interferons (IFN). Type I and Type III IFNs lead to induction of antiviral effectors (interferon-stimulated genes, ISGs) which provide both a cell-intrinsic state of viral resistance and help coordinate the generation of adaptive immune responses^2^. Importantly, SARS-CoV-2 proteins interfere with induction of these IFNs^2–4^, and deficient IFN production has been associated with increased disease severity and poor outcomes^5,6^. Most studies evaluating the early host immune response have been performed in patients with severe disease^5,7–13^; signatures associated with patient outcomes among those with more mild to moderate disease remain less well defined^14^.

Naturally acquired immunity to SARS-CoV-2 results in protection from reinfection, mediated in part by SARS-CoV-2-specific memory T cell and antibody responses. However, there is considerable heterogeneity in the T cell and antibody response following natural SARS-CoV-2 infection, providing valuable opportunities to identify key immune components that are associated with protective immunity. The BNT162b2 (Pfizer–BioNTech) vaccine has been widely used throughout the world and is highly effective in preventing SARS-CoV-2 infection, as well as protecting patients from severe symptoms after infections^15^. Comparing responses induced by vaccination with those induced by natural SARS-CoV-2 infection could potentially guide researchers to better understand determinants of protective immunity and improve vaccine design.

Recent advances have highlighted the power of the tools of systems biology to comprehensively define the innate and adaptive immune response to infections, including SARS-CoV-2^5,5,7,9–13^. Such analyses have used host transcriptomics and proteomics, and assessments of the adaptive immune response, coupled with computational approaches, to define molecular signatures that predict clinical and immunologic outcomes of infection.

In this paper, we utilized a multi-omics approach to define early infection signatures following SARS-CoV-2 infection that predict subsequent symptom severity, oropharyngeal viral load, and memory immune responses. We leveraged longitudinal samples collected from outpatients enrolled in a randomized controlled trial of a type III interferon, Peginterferon Lambda-1a (Lambda, NCT04331899)^16^. In this trial, outpatients with initially mild to moderate COVID-19 were recruited within 72 hours of diagnosis, and followed through 7 months post-infection. We observed sequential activation of immune pathways in initially mild to moderate COVID-19 patients within the first 2 weeks of symptom onset, including interferon responses, NK cell activation, T cell activation and B cell responses. We identified variations in plasma proteins, early interferon signaling, and downstream cytokines (MCP1, MCP-2 and MCP-3) that were associated with multiple patient outcomes, including symptom severity, viral load, memory T cell activity and S protein-binding IgG levels measured up to 7 months after enrollment. By comparing the immune response in COVID-19 patients to the response to COVID-19 mRNA vaccine, we show that the immune response after the first dose of vaccination largely mirrors the trajectory of immune response after SARS-CoV-2 infection, including induction of the negative immune regulatory TWEAK, while the response to the second dose of vaccine is characterized by rapid activation of adaptive immunity and an absence of neutrophil response. Finally, we demonstrate that a machine learning model is able to predict symptom severity, T cell memory response and antibody response accurately using 8-10 plasma protein markers measured early during infection.

## Results

### Transcriptomic and proteomic profiles correlate with the time to symptom onset in COVID-19 patients

We recruited 108 participants with initially mild to moderate COVID-19 at diagnosis into this study. The median age of participants was 37 years (range 18-71) with 57% male, and 62% of Latinx ethnicity (Supplementary Table 1). Eight (6.7%) participants were asymptomatic at baseline. Of those with symptoms, the median duration of symptoms prior to randomization was 5 days.

Subjects were randomized to receive a single dose of Peginterferon Lambda or placebo at their first visit and followed up to 7 months post-enrollment (Figure 1). The median duration of viral shedding post-enrollment was 7 days, and symptoms was 8 days, and this did not differ between participants randomized to Lambda compared with placebo^16^. To profile the immune response in the COVID-19 patients, we conducted whole blood RNA-sequencing and plasma protein profiling with multiplex Olink panels (inflammation and immune response panels, n=184 proteins) using blood samples collected at day 0 and day 5 after enrollment. We assessed SARS-CoV-2-specific CD4+ T cell responses by intracellular cytokine staining using PBMC collected at day 28 after enrollment. We also measured IgG binding titers against the SARS-CoV-2 full length spike protein (S) using plasma collected at day 0, day 5, day 28, and month 7 (Figure 1).

**Figure 1:**
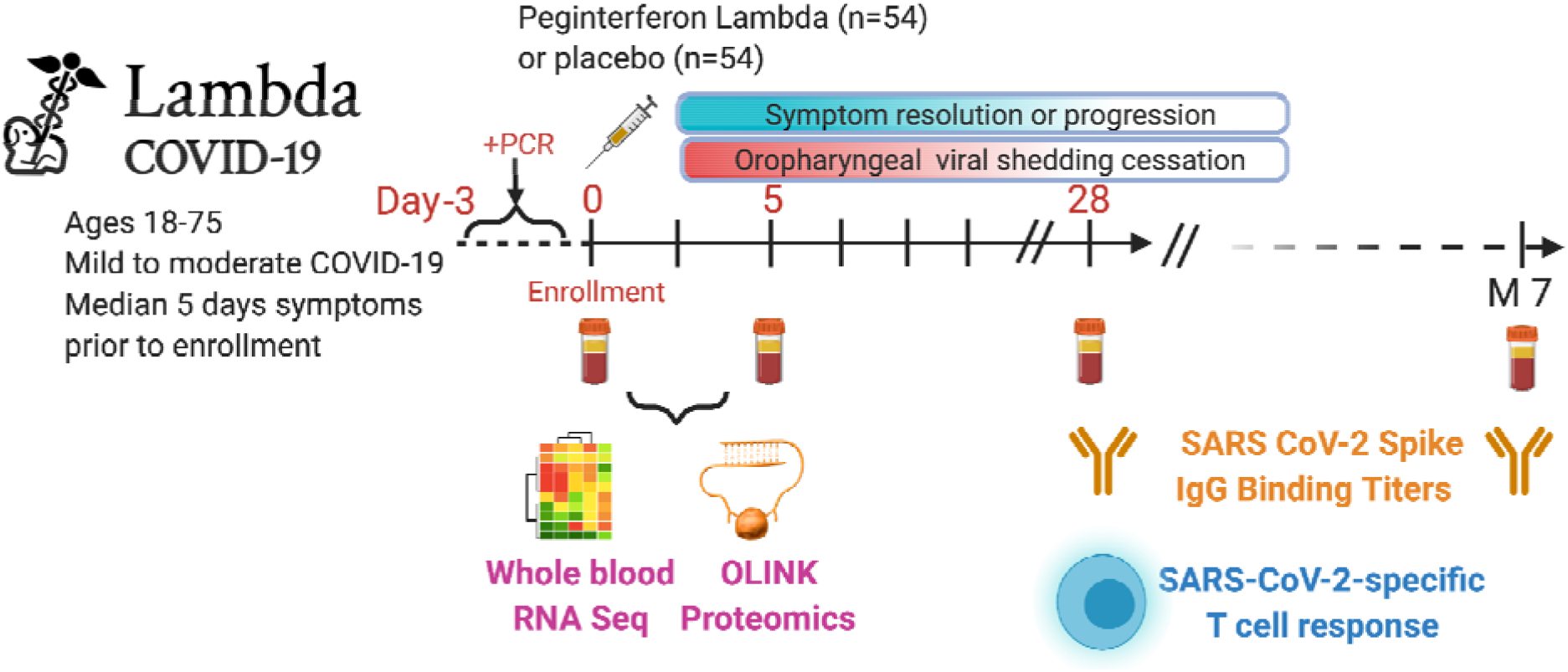
Study schema. Outpatients (n=108) with PCR-confirmed SARS-CoV-2 infection and swab obtained within 72 hours of randomization were enrolled in a Phase 2 clinical trial of subcutaneous Peginterferon lambda vs. placebo. In-person follow-up visits were conducted at day 1, 3, 5, 7, 10, 14, 21, 28,and month 7 post-enrollment, with assessment of symptoms and vitals, and collection of oropharyngeal swabs for SARS-CoV-2 testing. Blood obtained at Day 0 and 5 were evaluated by whole blood transcriptomics (RNA Sequencing), plasma proteomics (Olink), and SARS-CoV-2 specific antibodies. Clinical outcomes assessed included duration of symptoms and duration of virologic shedding. Immunologic outcomes assessed including SARS-CoV-2-specific T cell responses at day 28, and antibody responses at day 28 and month 7. Created with biorender.com.

We first examined antibody levels and transcriptomic profiles at day 0 and day 5 after enrollment in both patients randomized to Peginterferon Lambda and placebo. Based on the subject-reported symptom starting date, these samples were collected -1 to 20 days after symptom onset, with most of the samples collected within the first 2 weeks of the symptom onset (Figure 2A). As expected, we observed a positive correlation between the S protein binding IgG levels at enrollment and the time since symptom onset17 (Figure 2B). We performed principal component analysis of transcriptomic data and calculated the correlation between the first two principal components (PC) and other clinical variables. We found that PC1 had the strongest association with the time since symptom onset and the IgG titer, suggesting that these transcriptomic profiles capture the progression of the immune response in COVID-19 patients (Figure 2C-E). We also performed PCA analysis on the Olink data. Similar to results from the analysis of transcriptomics data, Olink data were associated with disease progression, as indicated by the high correlation between PC2 and the time since symptom onset (Figure 2F-H). We also observed an association between PC1 and age, which captures the impact of age on the plasma protein landscape in COVID-19 patients.

**Figure 2:**
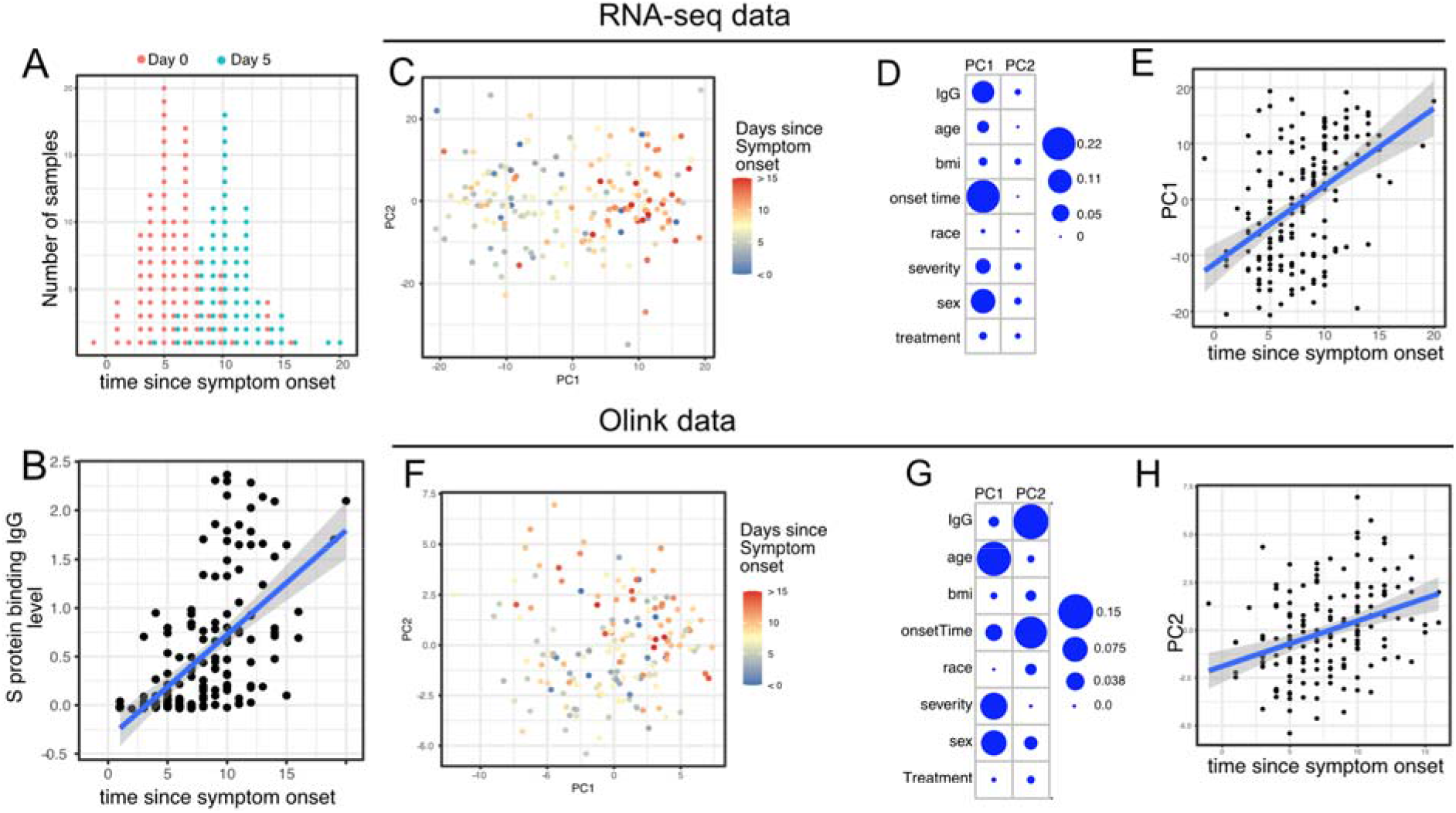
Transcriptomics and proteomics profiles correlate with the time to symptom onset in COVID-19 patients. (A) The distribution of RNA-seq sample collection time in respect to symptom onset. The colors of the dots represent the sample collection time from the enrollment. Asymptomatic cases are not shown. (B) Scatter plot showing the positive correlation between SARS-CoV-2 spike (S) protein binding IgG antibody level and the time since symptom onset. (C) PCA plot of the RNAseq samples. The colors of the dots represent the sample collection time from the enrollment. (D) the percent of the variances of PC1 and PC2 explained by different clinical variables. (E) scatter plot showing the positive correlation between PC1 of the RNA-seq data and the time since symptom onset. (F) PCA plot of the Olink proteomics data. The colors of the dots represent the sample collection time from the enrollment. (G) the percent of the variances of PC1 and PC2 explained by different clinical variables. (H) scatter plot showing the positive correlation between PC2 of the Olink data and the time since symptom onset.

We previously reported that Peginterferon Lambda treatment neither shortened the duration of SARS-CoV-2 viral shedding nor improved symptoms in outpatients with COVID-19^16^. PCA analysis revealed that transcriptional and proteomics profiles at day 5 post-treatment were not affected by Peginterferon Lambda treatment (Figure 2D, 2G, and Supplemental Figure 1). In addition, we found no significant differences in the T cell responses (at day 28 after enrollment) and antibody responses (at day 28 and month 7 after enrollment) between the two treatment arms (Supplemental Figure 1), as reported previously^17^. Taken together, Peginterferon Lambda treatment did not show noticeable effects on the immune response in COVID-19 outpatients.

Therefore, we combined the data from the control and treatment arms together for all downstream analysis.

### Trajectory analysis reveal sequential activation of immune pathways in COVID-19 patients

We next characterized the trajectory of early transcriptomic and proteomic responses using the RNAseq and Olink data as a function of time since symptom onset. To reduce the dimensionality and improve interpretability, we calculated the enrichment score of different immune pathways (based on Gene Ontology^18^) from the RNAseq data. We then combined pathway enrichment scores and Olink measurements into a single dataset for downstream statistical analysis. We fitted the data with quadratic regression to capture the non-linear dynamics of the pathways and proteins. We identified 38 immune pathways and 10 plasma proteins that varied as a function of time since symptom onset (False Discovery Rate (FDR) < 0.05, Figure 3A and Supplemental Table 2). Among them, 16 immune pathways or proteins showed nonlinear dynamics, as indicated by significant coefficients of the quadratic term (Supplemental Table 2).

**Figure 3:**
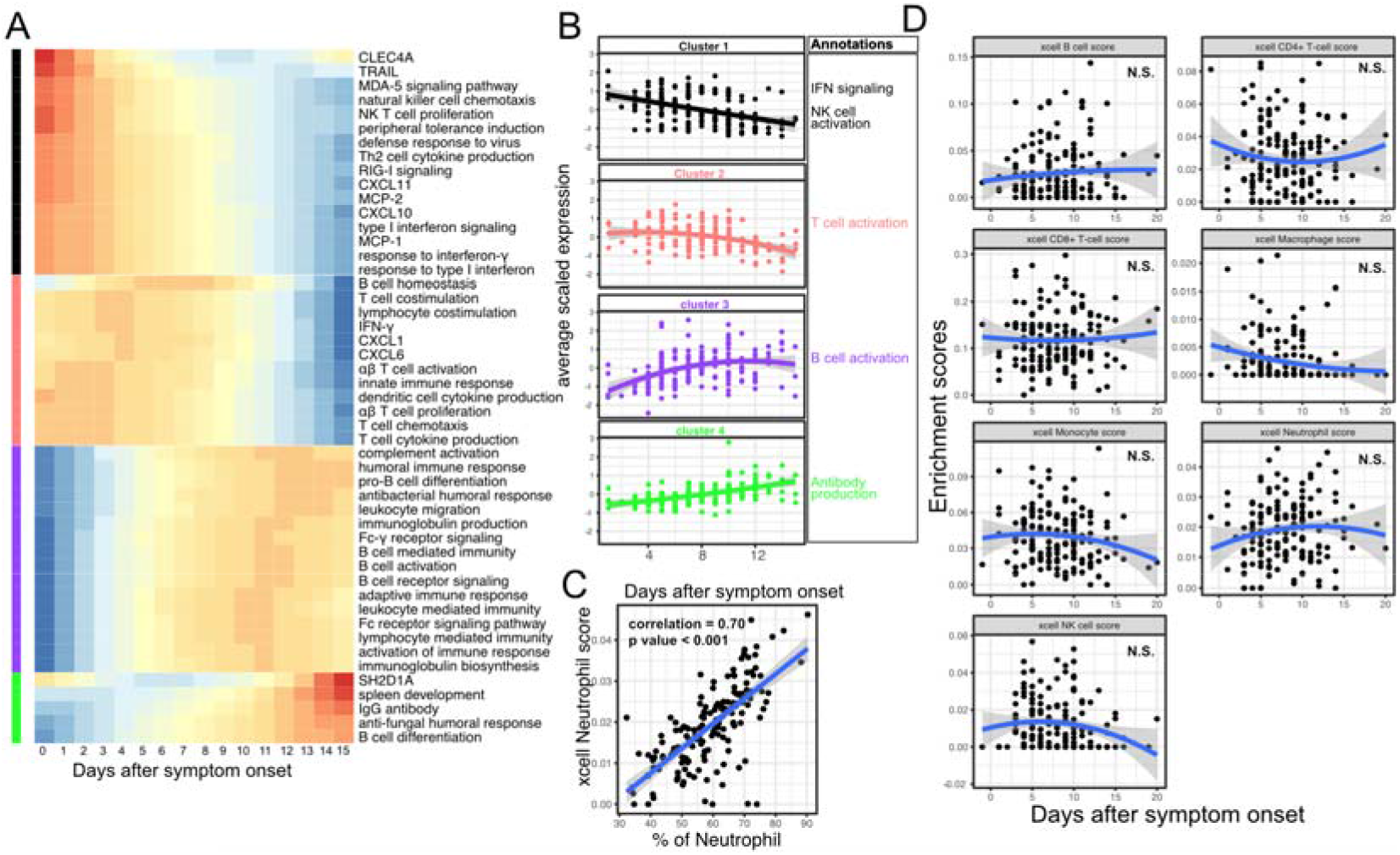
Trajectory analysis reveals sequential activation of immune pathways in COVID-19 patients. (A) The fitted expression level of immune pathways and plasma proteins at 0-15 days after symptom onset. The values are calculated by fitting quadratic regressions and are scaled to a mean of 0 and a standard deviation of 1. The color bar on the left side shows the clustering membership of the pathways and plasma proteins. (B) the average trajectory of the clusters. We scaled the expression level of each pathway and plasma proteins to a mean of 0 and a standard deviation of 1. We then calculated the average scaled expression of all members in the clusters. Each dot represents the mean expression in each blood sample. The lines represent the fitted quadratic regression. The grey areas represent the 95% confidence intervals. (C) We estimated the spearman correlation between the neutrophil enrichment score using the xCell. The plot shows the correlation between the xCell score and the counted neutrophil percentage in whole blood. (D) The relationship between xCell enrichment score and days after symptom onset.

We performed clustering analysis and identified four clusters based on the trajectory of the significant pathways and proteins (Figure 3A-B). Cluster 1 contains interferon-related pathways, natural killer cell activation pathways and proteins known to be activated by interferon signaling, including MCP-1, MCP-2, CXCL10 and CXCL11^19–22^. The trajectories in cluster 1 already reached the peak at the time of symptom onset and monotonically decreased over time. The trajectories in cluster 2 peaked at 1-5 days after symptom onset and contain Interferon-γ and pathways related to T cell activation. Interestingly, it also contains several myeloid cell attracting chemokines (CXCL1 and CXCL6) and the innate cell response pathway. Cluster 3 peaked between 10 to 14 days after the symptom onset and is characterized by pathways related to B cell activation. Cluster 4 trajectories monotonically increase after symptom onset and are characterized by the increasing S protein binding IgG level and related B cell differentiation pathways. The trajectory analysis revealed the sequential activation of interferon signaling, NK cells, myeloid cells, Interferon-γ, T cells, B cell and antibody production within the first 15 days of symptom onset.

To characterize how the composition of blood immune cells change over time, we used a previously established tool named xCell to estimate the enrichment score of the major immune cells^23^. As a positive control, we compared the neutrophil score with the neutrophil count data obtained from clinical lab tests and found high correlation between them (Figure 3C). Quadratic regression did not find significant associations between the major cell types and the time since symptom onset (Figure 3D). The results suggest that the trajectory of different immune pathways (Figure 3A) are mainly driven by the activation of corresponding immune cells rather than the composition change of major immune cell types.

### Variations in early immune responses are associated with disease severity in COVID-19 patients

We next sought to identify immune pathways and plasma proteins associated with symptom severity in COVID-19 outpatients. At the time of sample collection (day 0 and day 5 after enrollment), the majority of subjects showed either mild to moderate symptoms that subsequently resolved (n=100) or were asymptomatic (n=8). However, 8 patients later developed progressive and more severe symptoms and were hospitalized or presented to the emergency department (median 2 days to progression, range 1-13 days). We defined these individuals as severe COVID-19, and used regression models to identify immune pathways and plasma proteins to compare these participants with those who didn’t seek care at the hospital (mild/moderate COVID19), while controlling for days after symptom onset.

As two positive controls, we confirmed well documented findings that lymphocyte percentages were negatively correlated with symptom severity and neutrophil percentages were positively correlated with symptom severity (Figure 4A)^24^. In addition, our regression analysis identified 17 immune pathways and 24 plasma proteins that are significantly associated with symptom severity (FDR<0.05, Figure 4B-C, and Supplemental table 3). The proteins and pathways from cluster 1 (as identified above in Figure 3A) were significantly enriched (Fisher’s exact test, p < 0.001), including pathways related to interferon response, Rig-I signaling, NK cell activation and multiple protein markers known to be induced by interferon signaling (MCP-1, MCP-2 and CXCL11). The result highlights the association between early immune responses and symptom severity. Our regression analysis excluded the asymptomatic individuals, as their symptom onset date was unknown. To include the asymptomatic individuals, we performed one-way ANOVA analysis without adjusting for symptom onset time. The results from the ANOVA analysis were consistent with the regression analysis (Figure 4D).

**Figure 4:**
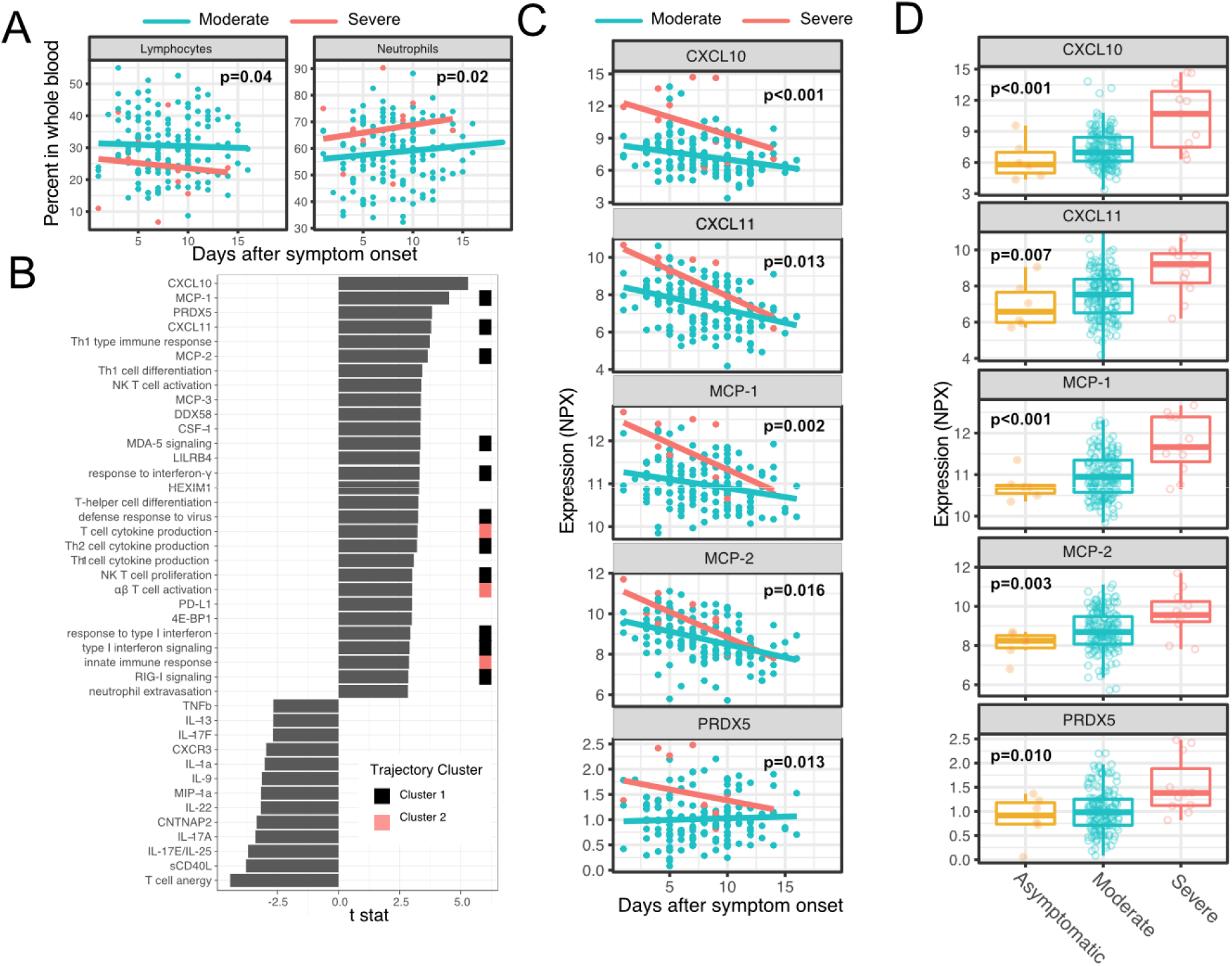
Variations in early immune responses are associated with disease severity in COVID-19 patients. (A) Scatter plot comparing the percentage of lymphocytes and neutrophils in whole blood between moderate and severe cases. The lines represent the fitted linear relationship between the percentages and the time after symptom onset. We fitted regression models to test the relationship between the immune measurements and the disease severity while controlling for the time after symptom onset. The p values for the disease severity are reported. (B) We fitted regression models to test the relationship between the immune measurements and symptom severity while controlling for the time after symptom onset. The bar plot shows the t score of the regression coefficient for symptom severity. The colored squares represent the clusters each immune measurement belongs to. The clusters are defined in Fig. 3A. (C) Scatter plot comparing the plasma protein levels between moderate and severe cases. The lines represent the fitted linear relationship between the percentages and the time after symptom onset. The top 5 significant proteins are shown. Data from asymptomatic cases are omitted, as their symptom onset time was unknown. (D) Box plots comparing the plasma protein levels between asymptomatic moderate and severe cases.

### Early proteomic and transcriptomic signatures show long-term association with virologic and immunologic outcomes

We examined associations between plasma proteins measured early in the course of infection and oropharyngeal viral load (measured by the area under the Ct curve from day 0 to 14 post enrollment). We identified 36 plasma proteins significantly associated with oropharyngeal viral load (top 10 significant proteins shown in Figure 5A, Supplemental table 3). Higher levels of several of these proteins were inversely correlated with viral load, including the cytosolic RNA sensor RIG-I (gene symbol DDX58), chemokines (CCL20 and CCL25), and other proteins (Keratin 19 {KRT19}, amphiregulin{AREG}) previously shown to be upregulated in COVID-19 patients^14,25^. Several immune transcriptomic pathways were also associated with viral control, including complement and B cell activation and the humoral immune response pathway (top 10 significant pathways shown in Figure 5B, Supplemental Table 3).

**Figure 5:**
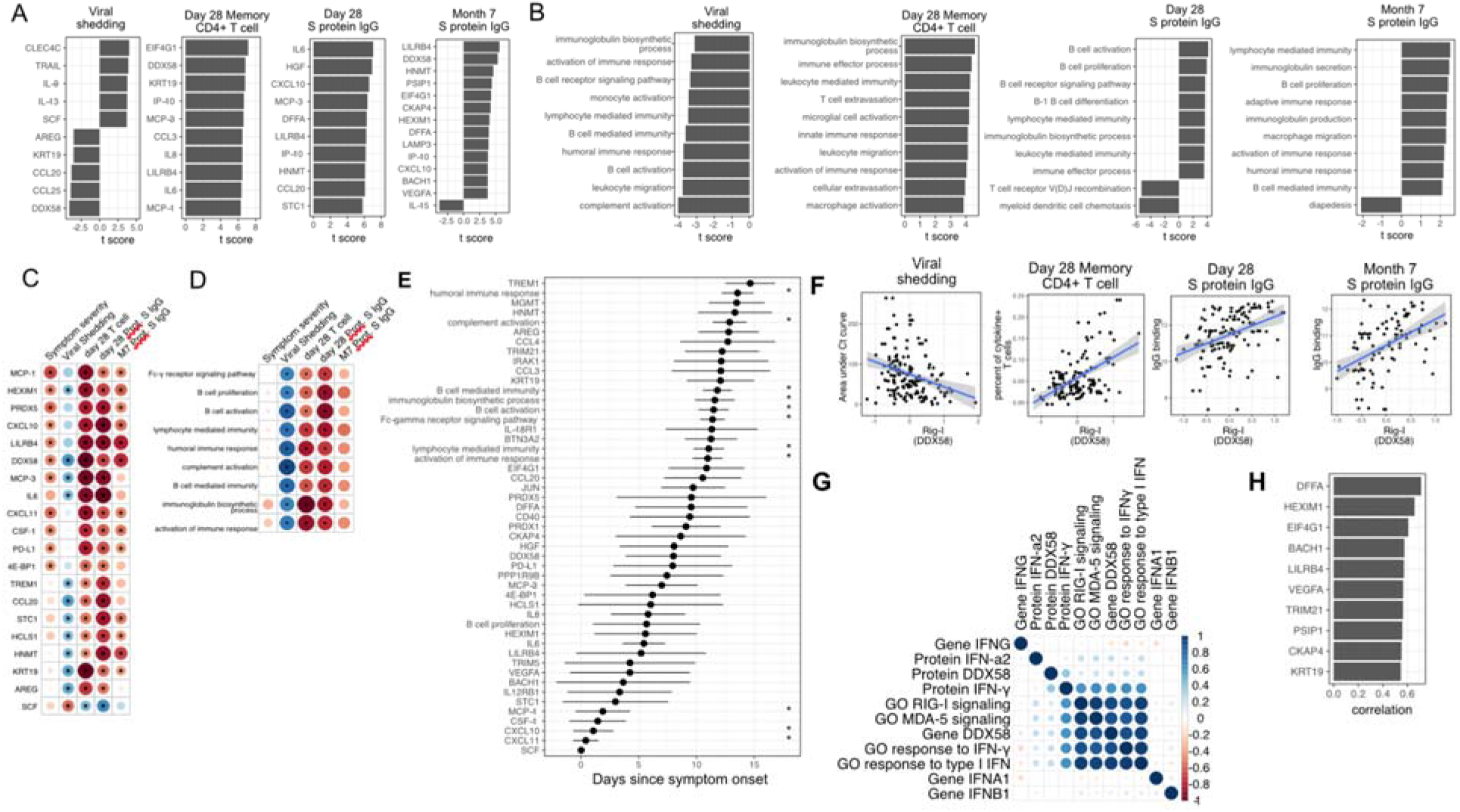
Plasma RIG-I is a biomarker for viral shedding, T cell activity, and spike (S)-binding IgG levels (A) The association between plasma proteins and viral shedding, memory T cell activity, and anti-S binding IgG levels. Memory CD4+ T cell activities are measured by the percent of cytokine positive T cells (TNF-α+ or TNFγ+ or IL21+) after spike protein stimulation. T cells are collected from patients 28 days after enrollment. Spike protein-binding IgG levels are measured 28 days or 7 months after enrollment. We fitted regression models to test the relationship between the immune measurements and viral shedding, memory T cell activity, and S protein binding IgG level while controlling for the time after symptom onset. The bar plot shows the t score of the regression coefficient for viral shedding, memory T cell activity, and anti-S binding IgG levels. (B) The association between immune pathways and viral shedding, memory T cell activity, and S protein binding IgG levels. (C-D) Association between plasma proteins and immune pathways that are associated with multiple outcomes. The heatmaps include immune measurements that are significantly associated (indicated by stars) with at least 3 outcomes. (E) The estimated time during which an immune measure reaches the maximum level. The error bars represent standard deviation derived from bootstrapping. The immune measures with stars are significantly associated with time. (F) Correlation between plasma RIG-I (DDX58) and viral shedding, memory T cell activity, and S protein binding IgG levels. (G) Correlation between plasma RIG-I protein and selected level of plasma proteins, genes, and pathways. (H) The top 10 plasma proteins correlated with plasma RIG-I protein.

We next measured associations between early plasma proteins, gene expression and subsequent immune memory, including cytokine-producing SARS-CoV-2-specific T cells measured 28 days post-enrollment (Supplemental Figure 2; Supplemental Table 3), and SARS-CoV-2-specific antibodies measured at 28 days and 7 months post-enrollment. We identified 87 plasma proteins that were significantly associated with SARS-CoV-2-specific T cell responses at day 28, and 91 and 13 plasma proteins significantly associated with S protein-binding IgG at day 28 days and month 7, respectively (top 10 significant proteins shown in Figure 5A, Supplemental table 3). Several proteins were associated with higher levels of SARS-CoV-2-specific T cells and the antibody response (Figure 5E), including RIG-I (gene symbol DDX58), chemokines (CXCL11), and other proteins (KRT19, AREG) also associated with control of viral load. Induction of early transcriptomic pathways were also associated with the development of SARS-CoV-2 specific T cells and S protein-binding IgG at day 28 (top 10 significant pathways shown in Figure 5B, Supplemental Table 3).

We further examined immune pathways and proteins associated with multiple clinical, virologic, and immune outcomes in COVID-19 patients. We identified 21 plasma proteins and 15 immune pathways that are correlated with three out of four aspects of the patient outcomes (Figure 5C and D). These include expected direct links (e.g. the correlation between immunoglobulin production[pathway and S protein-binding IgG) and more indirect links (e.g. the correlation between proinflammatory cytokines and S protein-binding IgG) between immune measurements. To establish a sequential order of pathway activation and protein expression, we fit a quadratic regression for each measurement, and then identified the time when the measurement reached maximal expression (Supplemental Figure 2A). This revealed 4 plasma proteins and 12 immune pathways whose expression were significantly associated with time. Among them, the interferon-gamma response, T cell cytokine production and several interferon induced cytokines (MCP1, MCP2, CXCL10 and CXCL11) reached maximal expression within the first 5 day after symptom onset (Figure 5E). Pathways related to B cell activation and antibody production reached maximum the latest, 10 to 15 days after symptom onset. A previous GWAS study has shown that symptom severity is associated with genetic variations near the interferon receptors (INFAR2) and CCR2 (receptor for MCP1, MCP2 and MCP3) loci. In addition, these variations were predicted to increase the expression of IFNAR2 and CCR2^26^. While the exact causal relationship cannot be established from our observational data, together our results suggest that the early interferon-related response and downstream CCR2 signaling shape later adaptive responses, and have long-term impact on the clinical, virological and immunological outcomes in COVID-19 patients.

Interestingly, plasma levels of RIG-I (gene symbol DDX58) were significantly associated with all examined virologic and immunologic outcomes (Figure 5F), as well as symptom severity (Figure 4B). Higher levels of plasma RIG-I were associated with less oropharyngeal viral load, more severe symptoms, increased SARS-CoV-2 specific T cell responses, and increased levels of S protein-binding IgG to SARS-CoV-2. Since RIG-I is a cytosolic PRR that, upon recognition of short viral double-stranded RNA during a viral infection, leads to upregulation of interferon signaling^27^, we explored associations between plasma RIG-I levels and related immune measurements, including the mRNA-level and protein-level expression of RIG-I and interferons, as well as RIG-I and interferon-related pathways. We found that the plasma RIG-I levels were modestly correlated with mRNA-level expression of RIG-I (correlation = 0.23, p value = 0.004, Figure 5G), as well as Rig-I signaling and Interferon related pathways (Figure 5C). Interestingly, we found a strong correlation between plasma level of RIG-I and plasma level of DFFA, an intracellular protein known to be involved in apoptosis (Figure 5H)^28,29^. In addition, the top 10 plasma proteins correlated with the plasma level of RIG-I are all intracellular proteins (Figure 5H). These data are consistent with the hypothesis that plasma RIG-I is associated with a cell death process that releases intracellular protein into the plasma. Furthermore, since plasma levels of RIG-I were not significantly associated with time (Fig 5E), these data suggest that plasma RIG-I levels might serve as a powerful and stable biomarker for predicting several clinical, viral and immunological outcomes in patients with COVID-19.

### Similar trajectories of immune responses induced by SARS-CoV-2 infection and COVID-19 mRNA vaccine

The BNT162b2 (Pfizer–BioNTech) vaccine has been widely used throughout the world and is highly effective in preventing SARS-CoV-2 infection, as well as protecting patients from severe symptoms after infections^15^. We leveraged a recently published Olink proteomics dataset from a BNT162b2 vaccine study^30^ to compare the immune response induced by COVID-19 vaccine and SARS-CoV-2 infections. Among the 66 protein markers shared between our SARS-CoV-2 infection dataset and the vaccination dataset, 8 proteins were significantly associated with time in the SARS-CoV-2 infection dataset, and 22 proteins were significantly associated with time in the vaccination dataset. It should be noted that the data collection in the vaccine study is highly synchronized, with well-defined vaccination day (day 0). In contrast, the time of infections in the SARS-CoV-2 infection dataset are only approximated by the self-reported time since symptom onset.

Comparison of the datasets reveals that the immune response after the first dose of vaccination (day 0 to day 21) largely mirrors the trajectory of immune response after SARS-CoV-2 infection. Early immune markers in the SARS-CoV-2 infection dataset, including IFNγ, MCP1, CXCL11, MCP2 and CXCL10 are upregulated within the first 7 days of the vaccination. Late immune markers in the SARS-CoV-2 infection dataset, including SLAMF1, TNFRSF9, CCL3, CCL4, TGFα and TNFSF14 are upregulated much later and show highest levels 21 days after the vaccination (Figure 6A). In contrast, the response after the second dose of vaccine (day 22 to day 28) is characterized by fast upregulation of both early and late protein markers (Figure 6A). Interestingly, three proteins that are significantly upregulated in SARS-CoV-2 patients were not induced after the second dose of vaccine, including TRAIL, CXCL1 and CXCL6. All three proteins are highly expressed in neutrophils^31^, and have been shown to regulate neutrophil recruitment (CXCL1 and CXCL6) or apoptosis (TRAIL) during inflammation^32–34^. The lack of TRAIL, CXCL1 and CXCL6 suggests an absence of neutrophil response to the second dose of vaccine.

**Figure 6:**
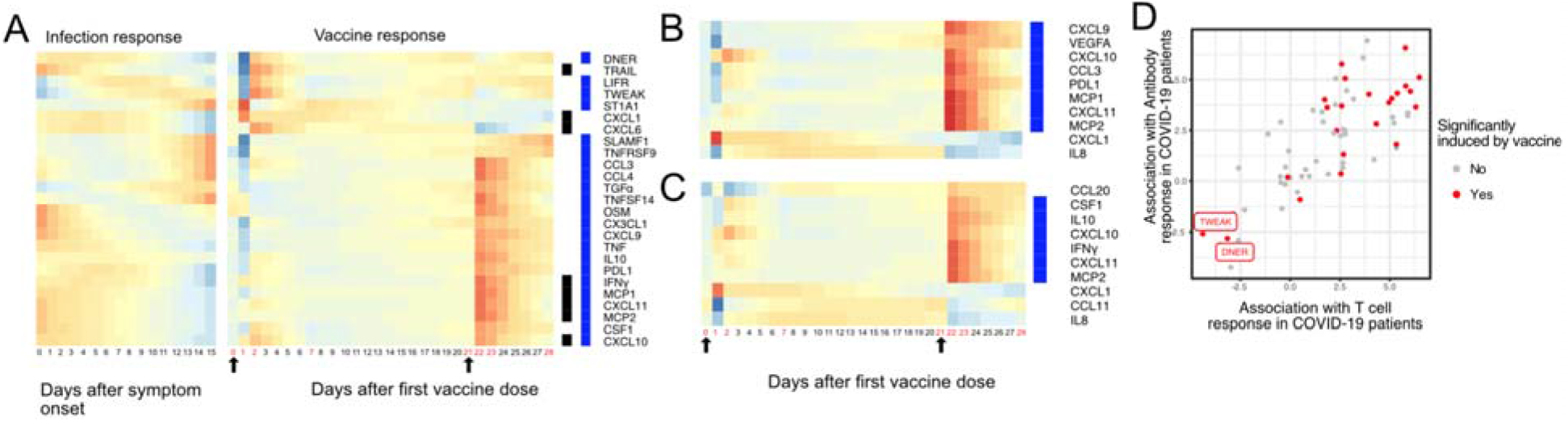
Comparing the immune response induced by SARS-CoV-2 infection and COVID-19 vaccine (BNT162b2). (A) The heat map shows the expression level of plasma proteins at 0-15 days after symptom onset in COVID-19 patients (left) and 0-28 days after vaccination in healthy individuals (right). The values from the SARS-CoV-2 dataset are calculated by fitting quadratic regressions and are scaled to a mean of 0 and a standard deviation of 1. The values from the vaccination dataset are computed by fitting using linear interpolation between the measured time points (days in red color) and are scaled to a mean of 0 and a standar deviation of 1. The black bar on the right side shows the protein markers that are significantly associated with time in COVID-19 patients (FDR<0.05). The blue bar on the right side shows the protein markers that are significantly associated with time after vaccination (FDR<0.05). The black arrows indicate the time of the first and the second doses of vaccination. (B) The vaccine response of the top 10 plasma proteins that are associated with CD4+ T cell response in SARS-CoV-2 patients. The blue bar on th right side shows the protein markers that are significantly associated with time after vaccination (FDR<0.05, p values are adjusted within the 10 protein set). (C) The vaccine response of the top 10 plasma proteins that are associated with spike (S)-binding IgG levels in SARS-CoV-2 patients. The blue bar on the right side shows the protein markers that are significantly associated with time after vaccination (FDR<0.05, p values are adjusted within the 10 protein set). (D) A scatter plot showing the plasma proteins profiled in both natural infection and vaccination studies. The X axis shows the association (t score of the regression coefficient) between plasma proteins and the SARS-CoV-2-specific memory CD4+ T cell response in natural infection. The Y axis shows the association (t score of the regression coefficient) between plasma proteins and the Spike protein binding IgG in natural infection. Points in red show if the protein is induced after vaccination.

Our analysis of the SARS-CoV-2 infection dataset reveals multiple plasma markers that are associated with T cell and antibody responses in COVID-19 patients (Figure 5). We next examined the trajectory of these proteins in response to the BNT162b2 vaccine. Among the top 10 proteins positively associated with CD4+ T cell response, 8 proteins are significantly induced by the vaccine (Figure 6B). Among the top 10 proteins positively associated with S protein-binding antibody levels, 6 proteins are significantly induced by the vaccine (Figure 6C). Similar to the result in Figure 6A, we found multiple neutrophil related proteins (CXCL1, IL8 and CCL11) to be induced by the first vaccine dose, but are absent in response to the second vaccine dose. Interestingly, we found that the vaccine also induced two proteins that are negatively associated with T cell and antibody response following natural infection, including TWEAK and DNER (Figure 6D). TWEAK has been known to attenuate the adaptive immunity by inhibiting STAT-1 and NF-κB^35^, suggesting that its induction could have a negative impact on the protective immunity against COVID-19. Taken together, our comparative analysis shows that the proteomic response of the BNT162b2 vaccine mirrors in many ways the proteomic response after SARS-CoV-2 infection. At the same time, we found important distinctions, including fast activation of adaptive immunity, an absence of neutrophil response in response to the second dose of vaccine and the induction of TWEAK that may negatively affect the adaptive response.

### Plasma proteins predict symptom severity, T cell response and Spike protein-binding IgG levels in COVID-19 patients

We performed predictive modeling to test if plasma proteins measured early following infection can accurately predict symptom severity, oropharyngeal viral load, and SARS-CoV-2 specific memory T cell and antibody responses manifested later in the study. We adopted a computation pipeline to select a small subset of predictive biomarkers from the 184 proteins measured by Olink assays. We used a leave-one-out cross validation strategy to iteratively evaluate the model performance. We used Random Forest for feature selection and for building the final model (Figure 7A). Based on results from cross-validation, we selected between 8 to 10 protein markers measured at early infection to predict each of the five outcomes. The final models achieved cross-validation AUC of 0.84, 0.66, 0.77, 0.84 and 0.75 for predicting symptom severity, oropharyngeal viral load, memory t cell activity, day 28 spike protein binding IgG levels and month 7 spike protein binding IgG levels, respectively (Figure 7B).

**Figure 7:**
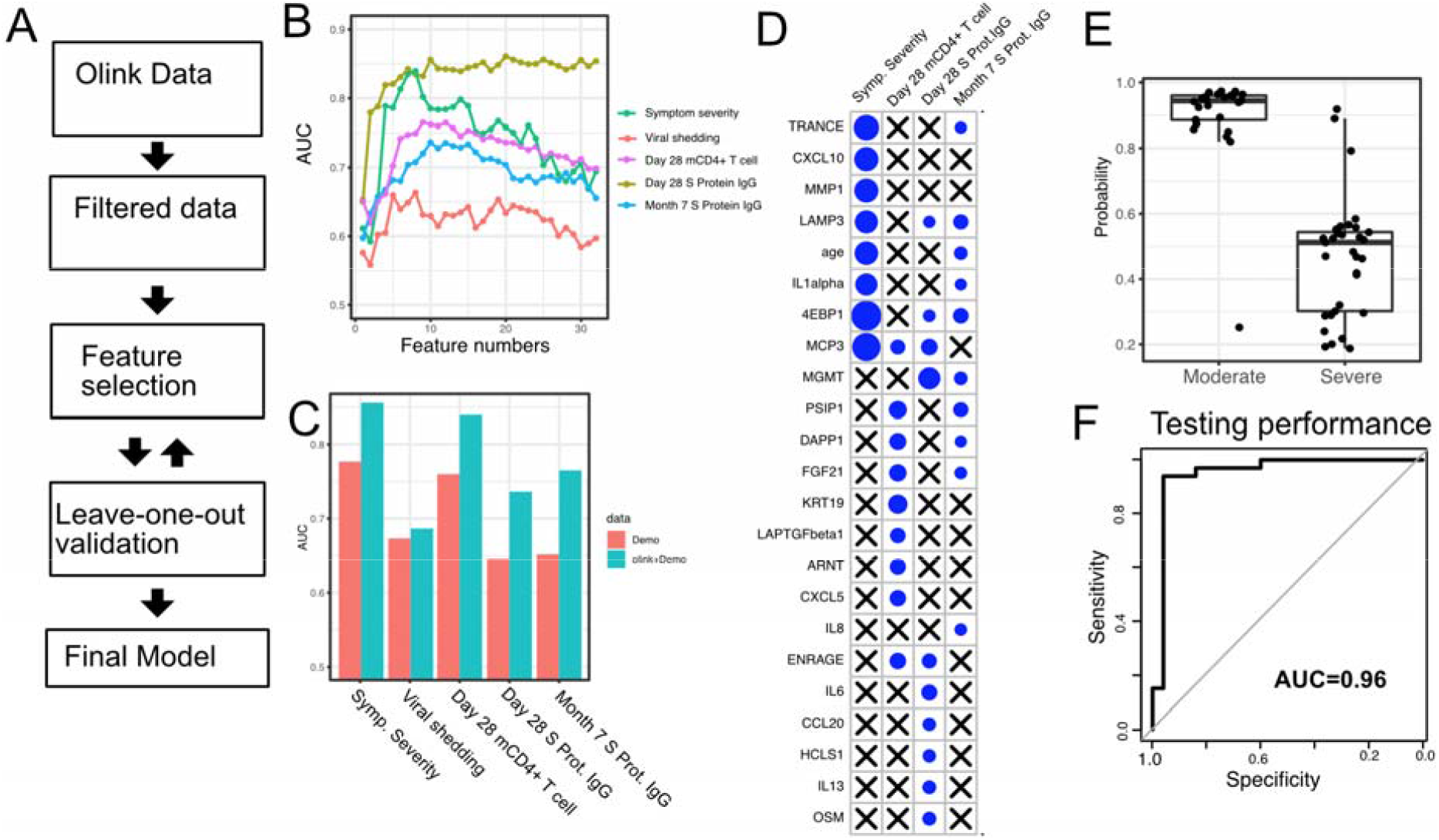
Plasma protein markers predict symptom severity, T cell response, and S protein-binding IgG level in COVID-19 patients. (A) machine-learning procedure for predicting COVID-19 patient outcomes using Olink proteomics data. (B) Random forest models were built to predict symptom severity, S protein-binding IgG level at 28 days and 7 months after enrollment, and cytokine+ memory CD4+ T cells 28 days after enrollment. The plot shows the leave-one-out cross-validation performance (measured by AUC of the receiver operator characteristics curve(ROC)) achieved by random forest models with different numbers of features. (C) The leave-one-out cross-validation performance of the best performing models and the models using demographical data (age and sex) only. (D) Feature importance of the final random forest models for predicting symptom severity, S protein-binding IgG level at 28 days and 7 months after enrollment, and cytokine+ memory CD4+ T cells at 28 days after enrollment. (E) We used the final model to predict severe cases in an independent dataset. The y axis of the plot shows the predicted probability. The x-axis shows the observed severity. (F) The performance of the final model measured by the ROC curve.

We compared the final models to baseline models that use only demographic (age and gender) data. The selected protein markers substantially improved the prediction of symptom severity, spike protein binding IgG levels at day 28 and month 7, and memory T cell responses at day 28. On the other hand, protein markers did not improve the prediction for oropharyngeal viral load.

We further tested if our model can accurately predict symptom severity in an independent dataset. We identified a published dataset that characterized the plasma proteins from 58 COVID-19 patients (26 moderate cases and 34 severe cases)^36^. Our model was able to accurately identify severe cases in the independent dataset, achieving an AUC of 0.96. The individuals in the test dataset already manifested severe symptoms while our training dataset was collected before the severe symptom were shown, potentially explaining the higher model performance in the test dataset than in the training dataset.

## Discussion

In this study, we longitudinally characterized the early immune response in patients who initially presented with mild to moderate COVID-19. With transcriptomic and proteomic profiling, we reveal a sequential activation of interferon signaling, NK cells, T cells and B cells within 2 weeks of symptom onset. We also identified associations between early immune profiles and later clinical, virologic, and immunologic outcomes. These data suggest that variations in plasma RIG-I levels, early interferon signaling, and related cytokines (MCP1, MCP-2 and MCP-3) are associated with multiple aspects of patient outcomes, including symptom severity, viral shedding, and the SARS-CoV-2 specific T cell and antibody response measured up to 7 months after enrollment. We also observed that the immune response after the first dose of SARS-CoV-2 mRNA vaccination largely recapitulates the trajectory of immune response after SARS-CoV-2 infection while the response to the second dose of vaccine is characterized by fast upregulation of both early and late protein markers and absence of a neutrophil response. Finally, we demonstrate that a machine learning model is able to predict symptom severity, T cell memory response and antibody response accurately using 8-10 plasma protein markers.

We observed that high plasma RIG-I levels were associated with greater disease severity, T cell activity, and the antibody response, suggesting that plasma RIG-I is a biomarker for increased immune activity in COVID-19 patients. High plasma RIG-I levels were also associated with lower SARS-CoV-2 viral loads, suggesting a potential role of this protein in restricting early virus replication. RIG-I has been shown to be critically important in the response to several RNA viruses, including influenza virus, typically via interactions with the adapter protein mitochondrial antiviral-signaling protein (MAVS) and downstream Type I and Type III interferon upregulation. RIG-I was recently shown to play an important role in both sensing SARS-CoV-2 RNA and inhibiting SARS-CoV-2 replication in human lung cells, but not via downstream MAVS induction^37^. Rather, interactions between the RIG-I helicase domain and SARS-CoV-2 RNA induced an inhibitory effect on viral replication, independent of downstream interferon upregulation^37^. This may explain the rather modest correlations observed between plasma RIG-I and RIG-I signaling and interferon related pathways. In contrast, we observed significant correlations between plasma RIG-I levels and plasma levels of DFFA, an intracellular protein known to be involved in cell death^29^, as well as other intracellular proteins, suggesting that plasma RIG-I levels may reflect increased cellular apoptosis. This hypothesis is consistent with a recent report which observed significant associations between gene expression signatures of apoptosis in plasmacytoid dendritic cells with increased disease severity^9^.

Our analysis also suggests that the variations in the early immune response shape the long-term outcome of COVID-19 patients. However, the cause of variations in the early responses are not fully understood. We observed that higher expression of three CCR2 ligands (MCP1, MCP2 and MCP3) were associated with multiple patient outcomes in COVID-19 patients, including increased disease severity and higher T cell activity and S protein-binding IgG levels. In addition MCP1 and MCP3 are negatively associated with oropharyngeal viral load. Our result is consistent with previous studies that show CCR2 signaling is associated with symptom severity in multiple viral infections, including SARS-Cov-2 and influenza^38,39^. A previous GWAS study has also identified an association between COVID-19 symptom severity and genetic variations that leads to increased CCR2 expression (receptor to interferon induced MCP1, MCP2 and MPC3)^26^. On the other hand, mouse studies show that CCR2 is essential for the survival of mice after pathogen challenge^40–42^. Our study also shows that the CCR2 ligands are associated with positive outcomes in patients with COVID-19, including reduced oropharyngeal viral load, increased memory T cell and IgG antibody response. Taken together, the results demonstrated the complex role of CCR2 signaling in regulating immune response. While it’s essential for an effective immune response, it also leads to severe symptoms and tissue damage. Therapeutic strategies to balance the positive and negative effects of CCR2 may benefit the management of COVID-19 patients.

Although naturally acquired SARS-CoV-2 infection results in protective antibody and T cell immune responses, reinfections can occur, and the precise determinants driving susceptibility to reinfection remain unclear^43^. The BNT162b2 (Pfizer–BioNTech) vaccine has been shown to be highly effective in preventing SARS-CoV-2 infection^15^, although breakthrough cases have been increasingly reported since its approval^44^. Comparing the vaccine response with the immune response of natural infection may shed light on determinants of protective immunity to SARS-CoV-2, and potential ways to improve COVID-19 vaccines. Our analysis reveals that the proteomic response of the BNT162b2 vaccine mirrors in many ways the proteomic response after SARS-CoV-2 infection, including the induction of protein markers that are positively associated with T cell and antibody response in COVID-19 patients. At the same time, we found important distinctions. In particular, proteomic signatures of a neutrophil response were absent in response to the second dose of vaccine. Multiple studies have demonstrated the importance of neutrophils in protection against both viral and bacterial reinfection^45,46^, although it remains unclear whether a neutrophil response may be beneficial, or detrimental, following vaccination. In addition, we found that the vaccine also induced proteins that are negatively associated with T cell and antibody response following natural infection, including TWEAK^35^. Further studies will be required to determine whether inhibiting TWEAK could potentially improve vaccine efficacy.

Our study has some limitations. First, while we identified multiple associations between early immune measures and the outcome of COVID-19 patients, we did not establish causal relationships between them. Future studies are needed to perturb key immune pathways in the early immune response and test their effect on the patient outcomes. Second, our study measured the immune response during the first 2 weeks of symptom onset in COVID-19 patients. Earlier immune responses between the initial infection and symptom onset have not been characterized. This is due to the difficulty to detect pre-symptomatic COVID-19 infection. Routine SARS-COV-2 monitoring in a select cohort will be required to acquire samples prior to and immediately after the infection in order to assess whether pre-infection signatures predict outcomes in COVID-19 patients. Third, our analysis focused on individual plasma proteins (based on olink data) and immune related Gene Ontology pathways (based on RNA-seq data). We used the Gene Ontology-based pathways to provide a high-level overview of the immune response in COVID-19 patients. Caution should be taken when interpreting the Gene Ontology pathways results, as the pathways are manually curated gene lists from literature and subject to publication bias, curation errors and over-simplification of biological processes. We encourage others to investigate the immune response of individual genes of interest using our shared RNA-seq data. Finally, we have created machine learning models to predict multiple outcomes in COVID-19 patients, including symptom severity, T cell response and antibody responses. While we are able to validate the model for predicting symptom severity in an independent dataset, additional datasets are needed to validate our model for predicting T cell and antibody responses.

In this study, we identified multiple biomarkers for predicting clinical and immunological outcomes in COVID-19 patients, including plasma level of RIG-I and the CCR2 ligands (MCP1, MCP2 and MCP3). In addition, we demonstrate that machine learning models using 8-10 biomarkers are highly effective in predicting these outcomes. The models can potentially be used to identify high-risk COVID-19 patients who will develop life-threatening symptoms, and to predict the degree of immune memory development. In addition, these biomarkers and models could also help explain variations in the response to COVID-19 vaccines, and to further identify differences between natural infection and vaccine-induced immunity.

## Data Availability

The will be available upon request.

## Acknowledgements

Support for the study was provided from NIH/NIAID (U01 AI150741-01S1 to ZH, ST, IRB, BG, TW, and PJ), the Stanford’s Innovative Medicines Accelerator, and NIH/NIDA DP1DA046089. The Lambda clinical trial was funded by anonymous donors to Stanford University, and Peginterferon Lambda provided by Eiger BioPharmaceuticals. The funders had no role in data collection and analysis or the decision to publish.

We thank all study participants who participated in this study, the study team for their tireless work, and Thanmayi Ranganath, Nancy Q. Zhao, Aaron J. Wilk, Rosemary Vergara, Julia L. McKechnie, Giovanny J. Martínez-Colón, Arjun Rustagi, Geoff Ivison, Ruoxi Pi, Madeline J. Lee, Taylor Hollis, Georgie Nahass, Kazim Haider, and Laura Simpson for assistance with processing samples. We also thank our colleagues at Stanford University Occupational Health and at San Mateo Medical Center for participant referrals. The Stanford REDCap platform (http://redcap.stanford.edu) is developed and operated by Stanford Medicine Research IT team. The REDCap platform services at Stanford are subsidized by a) Stanford School of Medicine Research Office, and b) the National Center for Research Resources and the National Center for Advancing Translational Sciences, National Institutes of Health, through grant UL1 TR001085.

## Methods

### Lambda Study Design and Oversight

Data and samples were obtained from a Phase 2, single-blind, randomized placebo-controlled trial to evaluate the efficacy of Lambda in reducing the duration of viral shedding in outpatients. The trial was conducted within the Stanford Health Care System. Adults aged 18-65 years with an FDA emergency use authorized reverse transcription-polymerase chain reaction (RT-PCR) positive for SARS-CoV-2 within 72 hours from swab to the time of enrollment were eligible for participation in this study. We included both symptomatic and asymptomatic patients based on the previous finding that the detected infectious virus were similar in samples from asymptomatic and symptomatic persons^44^. Symptomatic individuals were eligible given the presence of mild to moderate symptoms without signs of respiratory distress. Asymptomatic individuals were eligible if infection was the initial diagnosis of SARS-CoV-2 infection. Exclusion criteria included current or imminent hospitalization, respiratory rate >20 breaths per minute, room air oxygen saturation <94%, pregnancy or breastfeeding, history of decompensated liver disease, recent use of interferons, antibiotics, anticoagulants or other investigational and/or immunomodulatory agents for treatment of COVID-19, and prespecified lab abnormalities. Full eligibility criteria are provided in the study protocol. The protocol was amended on June 16^th^, 2020 after 54 participants were enrolled but before results were available to include adults up to 75 years of age and eliminate exclusion criteria for low white blood cell and lymphocyte count. The trial was registered at ClinicalTrials.gov (NCT04331899). The study was performed as an investigator-initiated clinical trial with the FDA (IND 419217), and approved by the Institutional Review Board of Stanford University.

### Participant Follow Up and Sample Collection

Participants completed a daily symptom questionnaire using REDCap Cloud version 1.5. In-person follow-up visits were conducted at day 1, 3, 5, 7, 10, 14, 21, and 28, with assessment of symptoms and vitals, and collection of oropharyngeal swabs (FLOQ Swabs; Copan Diagnostics). Peripheral blood was collected at enrollment, day 5, day 28, and month 7 post randomization. Whole blood was collected in Paxgene Tubes, and remaining blood was processed for plasma and peripheral blood mononuclear cells.

### Clinical Laboratory procedures

Laboratory measurements were performed by trained study personnel using point-of-care CLIA-waived devices or in the Stanford Health Care Clinical Laboratory. Oropharyngeal swabs were tested for SARS-CoV-2 in the Stanford Clinical Virology Laboratory using an emergency use authorized, laboratory-developed, RT-PCR. Centers for Disease Control and Prevention guidelines identify oropharyngeal swabs as acceptable upper respiratory specimens to test for the presence of SARS-CoV-2 RNA, and detection of SARS-CoV-2 RNA swabs using oropharyngeal swabs was analytically validated in the Stanford virology laboratory.

### Whole blood transcriptomics

Whole blood transcriptomics were performed at Novogene Corporation, Inc. Briefly, whole blood samples collected in Paxgene Tubes were first treated with Proteinase K, and then RNA extraction performed using Quick-RNA MagBead Kit (R2132) on KingFisher followed by sample quality control checks using a Qubit and Bioanalyzer 2100. Libraries were prepared using ZymoSeq RiboFree Total RNA Library Kit (R3000). Sequencing took place on a Nova6000 on an S4 lane, 30M paired reads, PE 150.

### Whole blood transcriptomic data analysis

The transcript-level count data and transcript per million (TPM) data was calculated using Kallist^47^ (v0.46.2) and human cDNA index produced using kallisto on Ensembl v96 transcriptomes. We identified all gene ontology terms that are the child term of immune system process (GO:0002376). We removed highly redundant gene ontology terms by grouping terms with >80% overlap of genes and manually selected the representative terms within each group. For each RNA-seq sample, we calculated the single-sample enrichment score of each gene ontology term using the fgsea R package^48^. The enrichment scores of the gene ontology terms were normally distributed across samples and are treated as variables, similar to individual protein markers, in the downstream analysis.

### Plasma protein profiling using Olink panels

We measured proteins in plasma using Olink multiplex proximity extension assay (PEA) inflammation panel and immune response panel (Olink proteomics, www.olink.com) according to the manufacturer’s instructions. The PEA is a dual-recognition immunoassay, where two matched antibodies labeled with unique DNA oligonucleotides simultaneously bind to a target protein in solution. This brings the two antibodies into proximity, allowing their DNA oligonucleotides to hybridize, serving as a template for a DNA polymerase-dependent extension step. This creates a double-stranded DNA “barcode” unique for the specific antigen and quantitatively proportional to the initial concentration of target protein. The hybridization and extension are immediately followed by PCR amplification and the amplicon is then finally quantified by microfluidic qPCR.

### T Cell Assays

SARS-CoV-2 specific T cell peptide pools were purchased from Miltenyi Biotec (PepTivator® SARS-CoV-2 Prot_S, Prot_S1, Prot N, and Prot M) and resuspended in DMSO. These PepTivator® reagents are pools of lyophilized peptides of 15 amino acid length with 11 amino acid overlap, covering immunodominant sequence domains of the Spike (S and S1) (aa sequence 1-1273), Nucleocapsid (N) or Membrane (M) proteins of SARS-CoV-2.

Antigen-specific T cell responses were measured using an intracellular cytokine staining assay. Briefly, cryopreserved PBMCs were thawed, counted, and resuspended in complete RPMI (RPMI (Corning) supplemented with 10% FBS (Gibco), 100 IU Penicillin (Corning), 100 ug/ml Streptomycin (Corning), 1 mM Hepes (Corning) and 2 mM L-glutamine (Corning)). The cells were plated in 96-well U bottom plates at 1×10e6 PBMCs per well and then rested overnight at 37°C in a CO2 incubator. The following morning, cells were cultured in presence of either SARS-CoV-2 peptides (1 μg/ml), PMA (300 ng/ml) and Ionomycin (1.5 μg/ml) as positive control, or media as a negative control for 6 hours at 37°C. All conditions were in the presence of brefeldin A (BD Pharmingen), monensin (BD Pharmingen), and CD107a. After a 6-hour incubation, cells were washed and surface stained for CCR7 for 15 min at 37°C, followed by the remaining surface stain for 30 min at room temperature (RT) in the dark. Thereafter, cells were washed twice with PBS containing 0.5% BSA and 2 mM EDTA, then fixed/permeabilized (FIX & PERM® Cell Permeabilization Kit, Invitrogen) and stained with intracellular antibodies for 20 min at RT in the dark. A complete list of antibodies are listed in Supplementary Methods. All samples were analyzed on an Attune NXT flow cytometer and analyzed with FlowJo X (Tree Star) software.

### Antibody Assays

IgG antibody titers against the SARS-CoV-2 full length spike protein were assessed by enzyme-linked immunosorbent assay (ELISA)27. Briefly, 96 Well Half-Area microplates (Corning (Millipore Sigma)) plates were coated with antigens at 2 μg/ml in PBS for 1h at RT. Next, the plates were blocked for an hour with 3% non-fat milk in PBS with 0.1% Tween 20 (PBST). Plasma was diluted fivefold starting at 1:50 in 1% non-fat milk in PBST. 25 μl of the diluted plasma was added to each well and incubated for 2h at RT. Following primary incubation, 25 μl of 1:5000 diluted horse radish peroxidase (HRP) conjugated anti-Human IgG secondary antibodies (Southern Biotech) were added and incubated for 1h at RT. The plates were developed by adding 25 μl/well of the chromogenic substrate 3,3′,5,5′-tetramethylbenzidine (TMB) solution (Millipore Sigma). The reaction was stopped with 0.2N sulphuric acid (Sigma) and absorbance was measured at 450nm (iD5 SPECTRAmax, Molecular Devices). The plates were washed 5 times with PBST between each step and an additional wash with PBS was done before developing the plates. All data were normalized between the same positive and negative controls and the binding AUC were calculated using GraphPad PRISM (Version 9).

### Quantifying oropharyngeal viral load

We identified the cycle threshold (Ct) value using the fluorescence vs cycle data reported from RT-PCR scanner. We subtract the Ct value from the detect limit (Ct=41) to quantify the viral shedding in each OP swap. We plotted the viral shedding in each visit versus time, and calculated the area under the curve using numerical integration based on the trapezoid rule.

### Analysis of Olink data from the vaccine study

The olink data from the mRNA vaccine study was previously obtained and published ^30^.We tested if the level of the proteins are significantly altered after vaccination using ANOVA (expression ∼ time), where time is treated as a categorical variable to account for non-linear behavior of the proteins. P values from the ANOVA models are adjusted using the False Discovery Rate (FDR) method^49^. To visualize the trajectory of the proteins, we imputed the protein level in each day using linear interpolation with the ‘approx’ function in R.

### Statistical analysis

Principal component analysis was conducted by applying the prcomp function in base R to the whole Olink dataset or the top 500 genes with the highest variance. To access the association between the principal components and clinical data, we fitted regression models (PC ∼ clinical variable). The percent of variances explained by the clinical variable is used to measure the association.

We accessed the association between the expression of immune pathways or protein markers with time using the regression model (expression ∼ time + time^2^). It should be noted that our study contains repeated measures of the same individuals in two time points (0 and 5 days after enrollment). While including subjects as random effects in the regression model allows the model to adjust for individual differences, it resulted in near-singular fits of the data for many of the immune measurements. To avoid model over-fitting, we decided to only include the fixed effects (time) in our model. To find significant associations, we compared the model with the base model (expression ∼ 1) and used the F test to calculate the p value. We adjusted the p value using the False Discovery Rate (FDR) method. We performed a parallel analysis using mixed-effect models [expression ∼ time + time^2^ + subject ID(random effect)] to fit the data and found that all significant (FDR<0.05) variables identified using the fixed effect model were also significant in the mixed-effect model (Supplemental Table 2).

We estimated the enrichment score of the major immune cell types using the xCell package^23^. The association between the xCell scores and time were tested using the same regression method described above.

We tested the association between immune measurements and symptom severity using regression models (measurements ∼ symptom severity + time + time^2^) and the lm function in R. The p value of the symptom severity term is adjusted using the FDR method. Similar regression models were used to test the association between immune measurements and other outcomes, including the oropharyngeal viral load, the memory CD4+ T cell activity and S protein binding IgG levels. To test between immune measurements and symptom severity without adjusting the time to symptom onset, we performed a one-way anova analysis using the lm function in R (measurements ∼ symptom severity).

To estimate the time in which an immune measurement reaches the maximum level, we first fit a quadratic regression model (measurement ∼ time + time^2^). We then identified the day (between 0 - 15 after symptom onset) in which the fitted regression model reached the maxim. We repeated the process 100 times to estimate the variance of the time.

### Predictive modeling

We used the protein measurements (measured by Olink assays) to predict the clinical, virological and memory T cell activity and IgG antibodies. Since the outcomes are a mixture of categorical (symptom severity) and continuous (viral load, T cell and antibody responses) variables, we framed the all prediction tasks as classification problems by dichotomizing the continuous variables using median as cutoffs. To prevent overfitting, we selected 30 proteins with the highest variance as input data, as the highly variable proteins best capture the inter-subject difference across the COVID-19 patients. We further select features using random forest and leave-one-out validation. In step 1, we train a random forest model using data from all samples but one left out sample. In step 2, we rank the feature importance of the 30 protein markers based on the gini index reported by the random forest model. In step 3, We train reduced random forest models with 1-29 most important proteins. In step 4, we predict the outcomes using the data from the left out sample. We repeat steps 1-4 until we predict the outcome of all samples. We calculate the model performance using the area under the receiver operator characteristic curve (AUC). The variable combinations that give rise to the highest AUC are selected as the optimal model. The optimal model for predicting symptom severity was tested using Olink data from two independent studies.

### Data availability

The RNA-sequencing data, Olink, clinical, virological, and immunological, as well as the machine learning models, are available upon request.

All codes for data analysis are available upon request.

## References

1. Wu, Z. & McGoogan, J. M. Characteristics of and Important Lessons From the Coronavirus Disease 2019 (COVID-19) Outbreak in China: Summary of a Report of 72[314 Cases From the Chinese Center for Disease Control and Prevention. JAMA 323, 1239–1242 (2020).

2. Park, A. & Iwasaki, A. Type I and Type III Interferons - Induction, Signaling, Evasion, and Application to Combat COVID-19. Cell Host Microbe 27, 870–878 (2020).

3. Konno, Y. et al. SARS-CoV-2 ORF3b Is a Potent Interferon Antagonist Whose Activity Is Increased by a Naturally Occurring Elongation Variant. Cell Rep 32, 108185 (2020).

4. Thorne, L. G. et al. Evolution of enhanced innate immune evasion by the SARS-CoV-2 B.1.1.7 UK variant. bioRxiv 2021.06.06.446826 (2021) doi:10.1101/2021.06.06.446826.

5. Arunachalam, P. S. et al. Systems biological assessment of immunity to mild versus severe COVID-19 infection in humans. Science 369, 1210–1220 (2020).

6. Hadjadj, J. et al. Impaired type I interferon activity and inflammatory responses in severe COVID-19 patients. Science 369, 718–724 (2020).

7. Wilk, A. J. et al. A single-cell atlas of the peripheral immune response in patients with severe COVID-19. Nat Med 26, 1070–1076 (2020).

8. Abers, M. S. et al. An immune-based biomarker signature is associated with mortality in COVID-19 patients. JCI Insight 6, 144455 (2021).

9. Liu, C. et al. Time-resolved systems immunology reveals a late juncture linked to fatal COVID-19. Cell 184, 1836-1857.e22 (2021).

10. Lucas, C. et al. Longitudinal analyses reveal immunological misfiring in severe COVID-19. Nature 584, 463–469 (2020).

11. Schulte-Schrepping, J. et al. Severe COVID-19 Is Marked by a Dysregulated Myeloid Cell Compartment. Cell 182, 1419-1440.e23 (2020).

12. Wilk, A. J. et al. Multi-omic profiling reveals widespread dysregulation of innate immunity and hematopoiesis in COVID-19. J Exp Med 218, e20210582 (2021).

13. Stephenson, E. et al. Single-cell multi-omics analysis of the immune response in COVID-19. Nat Med 27, 904–916 (2021).

14. Talla, A. et al. Longitudinal immune dynamics of mild COVID-19 define signatures of recovery and persistence. bioRxiv 2021.05.26.442666 (2021) doi:10.1101/2021.05.26.442666.

15. Polack, F. P. et al. Safety and Efficacy of the BNT162b2 mRNA Covid-19 Vaccine. New England Journal of Medicine 383, 2603–2615 (2020).

16. Jagannathan, P. et al. Peginterferon Lambda-1a for treatment of outpatients with uncomplicated COVID-19: a randomized placebo-controlled trial. Nature Communications 12, 1967 (2021).

17. Chakraborty, S. et al. Divergent early antibody responses define COVID-19 disease trajectories. bioRxiv 2021.05.25.445649 (2021) doi:10.1101/2021.05.25.445649.

18. Ashburner, M. et al. Gene Ontology: tool for the unification of biology. Nat Genet 25, 25–29 (2000).

19. Lehmann, M. H. et al. CCL2 expression is mediated by type I IFN receptor and recruits NK and T cells to the lung during MVA infection. J Leukoc Biol 99, 1057–1064 (2016).

20. Lee, P. Y. et al. Type I interferon modulates monocyte recruitment and maturation in chronic inflammation. Am J Pathol 175, 2023–2033 (2009).

21. Dengel, L. et al. Differential effects of IFN-α and IFN-γ on upregulation of IFN-inducible chemokines CXCL9, CXCL10 and CXCL11. Cancer Res 68, 2149–2149 (2008).

22. Moll, H. P., Maier, T., Zommer, A., Lavoie, T. & Brostjan, C. The differential activity of interferon-α subtypes is consistent among distinct target genes and cell types. Cytokine 53, 52–59 (2011).

23. xCell: digitally portraying the tissue cellular heterogeneity landscape | Genome Biology | Full Text. https://genomebiology.biomedcentral.com/articles/10.1186/s13059-017-1349-1.

24. Goyal, P. et al. Clinical Characteristics of Covid-19 in New York City. New England Journal of Medicine 382, 2372–2374 (2020).

25. Reyes-Gibby, C. C. et al. Oral microbiome and onset of oral mucositis in patients with squamous cell carcinoma of the head and neck. Cancer 126, 5124–5136 (2020).

26. Pairo-Castineira, E. et al. Genetic mechanisms of critical illness in COVID-19. Nature 591, 92–98 (2021).

27. Matsumiya, T. & Stafforini, D. M. Function and Regulation of Retinoic Acid-Inducible Gene-I. Crit Rev Immunol 30, 489–513 (2010).

28. Thomas, D. A., Du, C., Xu, M., Wang, X. & Ley, T. J. DFF45/ICAD Can Be Directly Processed by Granzyme B during the Induction of Apoptosis. Immunity 12, 621–632 (2000).

29. Zhang, J. H. & Xu, M. DNA fragmentation in apoptosis. Cell Research 10, 205–211 (2000).

30. Arunachalam, P. S. et al. Systems vaccinology of the BNT162b2 mRNA vaccine in humans. Nature 1–10 (2021) doi:10.1038/s41586-021-03791-x.

31. Uhlén, M. et al. Tissue-based map of the human proteome. Science 347, (2015).

32. Jovic, S. et al. The neutrophil-recruiting chemokine GCP-2/CXCL6 is expressed in cystic fibrosis airways and retains its functional properties after binding to extracellular DNA. Mucosal Immunol 9, 112–123 (2016).

33. Sawant, K. V. et al. Chemokine CXCL1 mediated neutrophil recruitment: Role of glycosaminoglycan interactions. Sci Rep 6, 33123 (2016).

34. Renshaw, S. A. et al. Acceleration of human neutrophil apoptosis by TRAIL. J Immunol 170, 1027–1033 (2003).

35. Maecker, H. et al. TWEAK attenuates the transition from innate to adaptive immunity. Cell 123, 931–944 (2005).

36. Patel, H. et al. Proteomic blood profiling in mild, severe and critical COVID-19 patients. Scientific Reports 11, 6357 (2021).

37. Yamada, T. et al. RIG-I triggers a signaling-abortive anti-SARS-CoV-2 defense in human lung cells. Nat Immunol 22, 820–828 (2021).

38. Lin, K. L., Sweeney, S., Kang, B. D., Ramsburg, E. & Gunn, M. D. CCR2 Antagonist Prophylaxis Reduces Pulmonary Immune Pathology and Markedly Improves Survival during Influenza Infection. J Immunol 186, 508–515 (2011).

39. Chen, Y. et al. IP-10 and MCP-1 as biomarkers associated with disease severity of COVID-19. Molecular Medicine 26, 97 (2020).

40. Pamer, E. G. Tipping the balance in favor of protective immunity during influenza virus infection. PNAS 106, 4961–4962 (2009).

41. Lim, J. K. et al. Chemokine Receptor CCR2 is Critical for Monocyte Accumulation and Survival in West Nile Virus Encephalitis. J Immunol 186, 471–478 (2011).

42. Defects in macrophage recruitment and host defense in mice lacking the CCR2 chemokine receptor - PubMed. https://pubmed.ncbi.nlm.nih.gov/9362535/.

43. To, K. K.-W. et al. COVID-19 re-infection by a phylogenetically distinct SARS-coronavirus-2 strain confirmed by whole genome sequencing. Clin Infect Dis ciaa1275 (2020) doi:10.1093/cid/ciaa1275.

44. Birhane, M. et al. COVID-19 Vaccine Breakthrough Infections Reported to CDC — United States, January 1–April 30, 2021. MMWR Morb Mortal Wkly Rep 70, 792–793 (2021).

45. Fujisawa, H. Neutrophils Play an Essential Role in Cooperation with Antibody in both Protection against and Recovery from Pulmonary Infection with Influenza Virus in Mice. Journal of Virology 82, 2772–2783 (2008).

46. Sjöstedt, A., Conlan, J. W. & North, R. J. Neutrophils are critical for host defense against primary infection with the facultative intracellular bacterium Francisella tularensis in mice and participate in defense against reinfection. Infection and Immunity 62, 2779–2783 (1994).

47. Bray, N. L., Pimentel, H., Melsted, P. & Pachter, L. Near-optimal probabilistic RNA-seq quantification. Nat Biotechnol 34, 525–527 (2016).

48. Korotkevich, G., Sukhov, V. & Sergushichev, A. Fast gene set enrichment analysis. bioRxiv 060012 (2019) doi:10.1101/060012.

49. Benjamini, Y. & Hochberg, Y. Controlling the False Discovery Rate: A Practical and Powerful Approach to Multiple Testing. Journal of the Royal Statistical Society: Series B (Methodological) 57, 289–300 (1995).

