## Supplemental information for "Early immune responses have long-term associations with clinical, virologic, and immunologic outcomes in patients with COVID-19"

Supplementary Material

Supplementary Table 1: Baseline characteristics

|  | **Treatment arm** | |  |
| --- | --- | --- | --- |
|  | **Lambda (N=54)** | **Placebo (N=54)** | **Overall (N=108)** |
| **Age in years**, median (range) | 37 (18-65) | 36 (20-71) | 37 (18-71) |
| **Male**, n (%) | 32 (59.3%) | 30 (55.6%) | 62 (57.4%) |
| **Race / Ethnicity**, n (%) |  |  |  |
| Latinx | 30 (56.7%) | 37 (68.3%) | 67 (62.0%) |
| White | 18 (30.0%) | 12 (25.0%) | 30 (27.8%) |
| Asian | 4 (5.0%) | 5 (6.7%) | 9 (8.3%) |
| Native Hawaiian or other Pacific Islander | 2 (3.3%) | 0 (0%) | 2 (1.9%) |
| **BMI (kg/m^2^)**, median (IQR) | 27.6 (25.4-31.1) | 28.5 (24.8-32.3) | 27.7 (24.9-32.0) |
| **Comorbid conditions** |  |  |  |
| Hypertension | 9 (15.0) | 5 (8.3) | 14 (11.7) |
| Diabetes | 4 (6.7) | 8 (13.3) | 12 (10.0) |
| Asthma | 2 (3.3) | 2 (3.3) | 4 (3.3) |
| Heart Disease | 3 (5.0) | 1 (1.7) | 4 (3.3) |
| **Asymptomatic at baseline**, n (%) | 6 (10.0%) | 2 (3.3%) | 8 (6.7%) |
| **Duration of symptoms in days prior to randomization**, median (IQR) ^1^ | 4 (3-6) | 5 (3-5) | 5 (3-6) |
| **Baseline laboratory values**, median (IQR) |  |  |  |
| White blood cell (WBC) count, cells/μl | 5.5 (4.3-6.8) | 5.6 (4.0-7.5) | 5.5 (4.1-7.1) |
| Absolute lymphocyte count (ALC), cells/μl | 1.5 (1.2-1.9) | 1.5 (1.2-2.3) | 1.5 (1.2-2.2) |
| Aspartate aminotransferase, IU/L | 31 (26-41) | 30 (25-39.3) | 30 (25-41) |
| Alanine aminotransferase, IU/L | 32.5 (21-52.3) | 30.5 (23-47.5) | 31.5 (22-50.3) |
| **Baseline oropharyngeal SARS-CoV-2 cycle threshold, median (IQR) ^2^** | 30.9 (26.4-33.8) | 29.3 (26.4-34.3) | 30.3 (26.4-34.3) |
| **Baseline Log10 Viral Load, median (IQR) ^2^** | 4.2 (3.3 - 5.5) | 4.7 (3.2 - 5.5) | 4.4 (3.2 - 5.5) |
| **Baseline SARS-CoV-2 IgG seropositivity**, n (%) | 21 (35.0%) | 28 (46.7%) | 49 (40.8%) |

##### **Supplemental Table 4: ICS antibody panel**

| **ICS Antibody Panel** | | | | | |
| --- | --- | --- | --- | --- | --- |
| **Surface Antibodies** | **Fluorochrome** | **Clone** | **Vendor** | **Catalog** | **Amount Per 50 uL** |
| CCR7 | BV421 | G043H7 | BioLegend | 353208 | 2.5uL |
| CD14  CD19   LIVE/DEAD | BV510  BV510  Aqua | M5E2  HB19 | BioLegend  BioLegend  Invitrogen | 301842  302242  L34965 | 0.5uL  0.5 uL  0.25uL |
| CD45RA | BV605 | HI100 | BioLegend | 304134 | 0.4uL |
| CD4 | BV650 | RPA-T4 | BioLegend | 300536 | 1uL |
| CD8A | BV785 | RPA-T8 | BioLegend | 301046 | 1uL |
| CD107A | FITC | H4A3 | BioLegend | 328606 | 1uL |
| CD3 | APC-H7 | SK7 | BD | 560176 | 2.5uL |
| **Intracellular Antibodies** | **Fluorochrome** | **Clone** | **Vendor** | **Catalog** | **Amount Per 50 uL** |
| IFN-g | PerCP Cy5.5 | 4S.B3 | BioLegend | 502526 | 0.5uL |
| IL-21 | eFluor660 | eBio3A3-N2 | eBioscience | 50-7219-42 | 1.25uL |
| TNF | AF700 | MAb11 | BD | 557996 | 0.5uL |

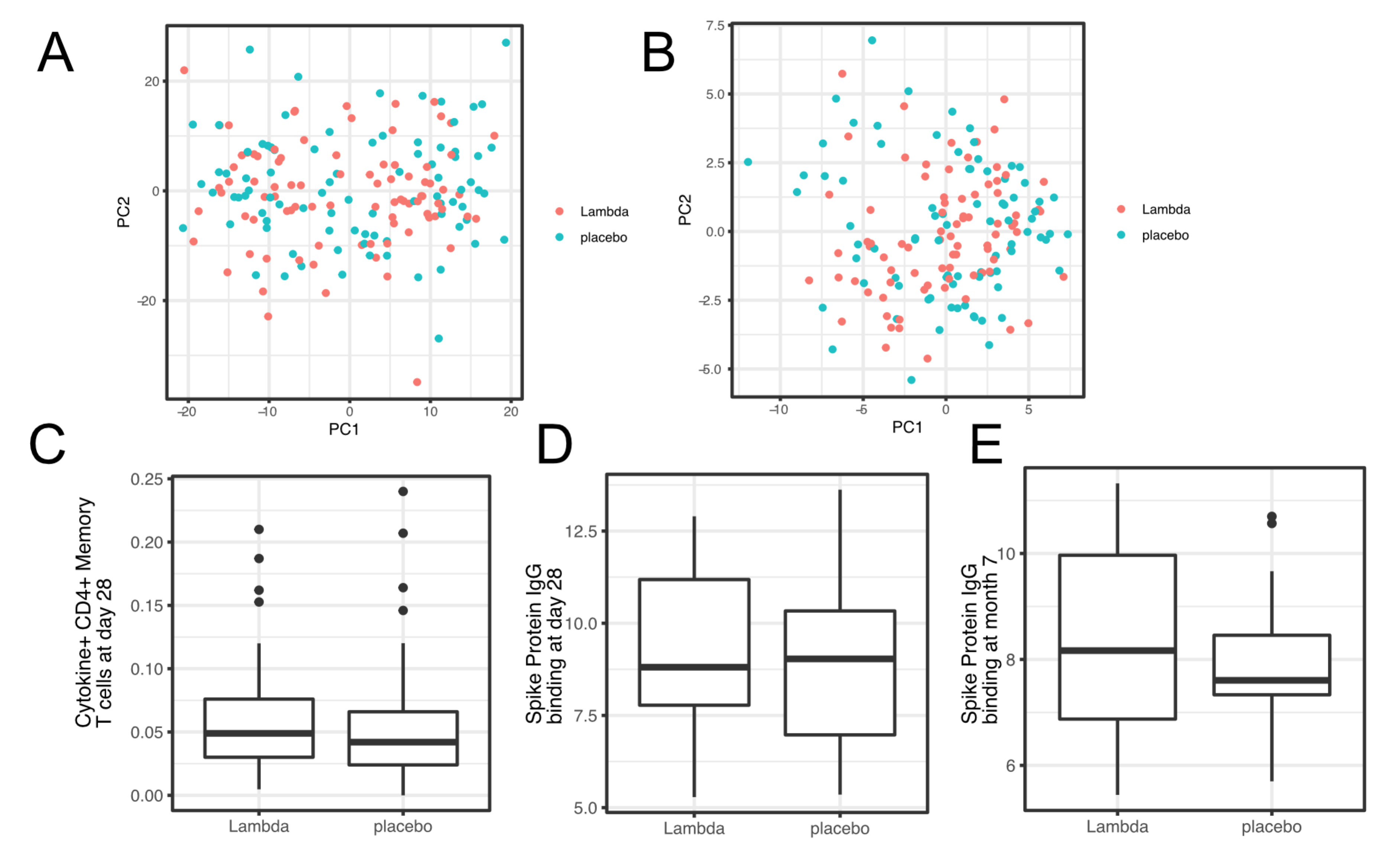

Supplemental Figure 1: Comparing transcriptomics (A), proteomics (B), T cell responses at day 28 after enrollment (C), and antibody responses (at day 28 (D) and month 7 (E) after enrollment) between the two treatment arms.

Supplementary Figure 2

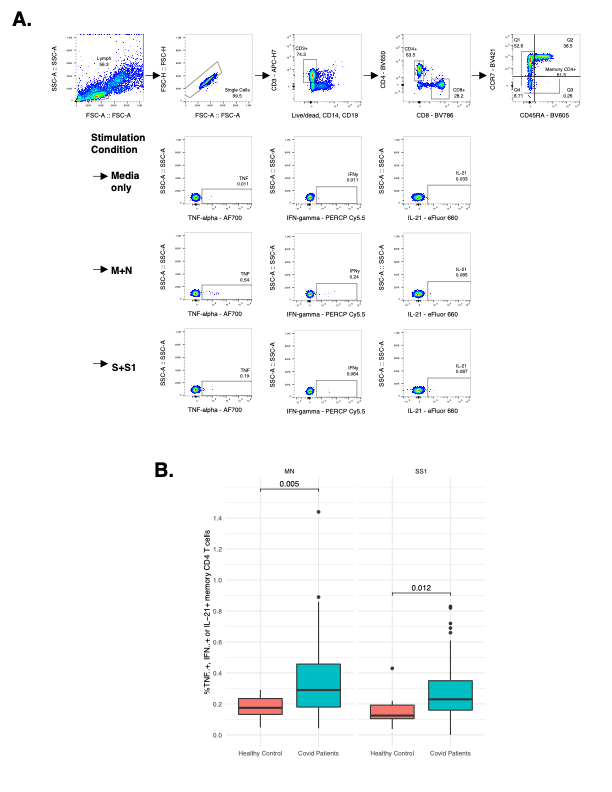

Supplementary Figure 2. Measurement of SARS-CoV-2 specific T cell responses by intracellular cytokine staining. A. Gating strategy to identify SARS-CoV-2 specific memory CD4+ T cells responsive to Membrane (M), Nucleocapsid (N), or Spike (S, S1) immunodominant peptides by detection of intracellular cytokines TNF-alpha, IFN-gamma, or IL-21. B. Comparison of SARS-CoV-2 specific T cell responses producing either TNF-alpha, IFN-gamma, or IL-21 following MN or SS1 stimulation compared to uninfected, healthy age-matched controls.
