## Supplemental table 2 for "Early immune responses have long-term associations with clinical, virologic, and immunologic outcomes in patients with COVID-19"

| gene | overall_p_value_linear_regression | quadratic_term_p_value_linear_regression |
|---|---|---|
| GO_activated_T_cell_proliferation | 0.892575831 | 0.822833174 |
| GO_activation_of_immune_response | 3.24E-05 | 0.000351451 |
| GO_activation_of_innate_immune_response | 0.075158864 | 0.320789006 |
| GO_adaptive_immune_response | 5.75E-05 | 0.000247383 |
| GO_alpha-beta_T_cell_activation | 0.002748293 | 0.194334121 |
| GO_alpha-beta_T_cell_differentiation | 0.012023025 | 0.154950308 |
| GO_alpha-beta_T_cell_lineage_commitment | 0.274129862 | 0.157512516 |
| GO_alpha-beta_T_cell_proliferation | 0.003783833 | 0.317212528 |
| GO_antibacterial_humoral_response | 0.000282968 | 0.02504821 |
| GO_antibacterial_peptide_production | 0.024051683 | 0.432330689 |
| GO_antifungal_humoral_response | 0.001363584 | 0.594429282 |
| GO_antifungal_innate_immune_response | 0.045865787 | 0.322901033 |
| GO_antigen_processing_and_presentation | 0.034479512 | 0.035769227 |
| GO_antimicrobial_humoral_response | 0.012480372 | 0.450585016 |
| GO_antimicrobial_peptide_production | 0.024051683 | 0.432330689 |
| GO_B_cell_activation | 1.19E-05 | 0.000751704 |
| GO_B_cell_chemotaxis | 0.037760087 | 0.210928088 |
| GO_B_cell_differentiation | 0.001429296 | 0.48077151 |
| GO_B_cell_homeostasis | 0.005252107 | 0.014799179 |
| GO_B_cell_lineage_commitment | 0.115640976 | 0.680922813 |
| GO_B_cell_mediated_immunity | 1.48E-07 | 7.29E-05 |
| GO_B_cell_proliferation | 0.601013309 | 0.324755909 |
| GO_B_cell_receptor_signaling_pathway | 4.88E-07 | 0.000131447 |
| GO_B-1_B_cell_differentiation | 0.354281627 | 0.770427794 |
| GO_bone_marrow_development | 0.013381452 | 0.709441482 |
| GO_cellular_extravasation | 0.089775964 | 0.415799327 |
| GO_complement_activation | 2.57E-07 | 0.000743759 |
| GO_defense_response_to_virus | 7.20E-09 | 0.751333503 |
| GO_definitive_hemopoiesis | 0.120992524 | 0.884804085 |
| GO_dendritic_cell_chemotaxis | 0.139004896 | 0.075017418 |
| GO_dendritic_cell_cytokine_production | 8.75E-05 | 0.21899716 |
| GO_dendritic_cell_differentiation | 0.468837165 | 0.749172487 |
| GO_dendritic_cell_migration | 0.144772328 | 0.061278398 |
| GO_diapedesis | 0.24807129 | 0.681343902 |
| GO_embryonic_hemopoiesis | 0.098144985 | 0.472365829 |
| GO_eosinophil_activation | 0.19536512 | 0.072295705 |
| GO_eosinophil_chemotaxis | 0.045123271 | 0.01314166 |
| GO_eosinophil_degranulation | 0.224209645 | 0.103396026 |
| GO_eosinophil_differentiation | 0.329054217 | 0.279737744 |

| | | |
|---|---|---|
| GO_eosinophil_mediated_immunity | 0.224209645 | 0.103396026 |
| GO_eosinophil_migration | 0.079220913 | 0.02557989 |
| GO_erythrocyte_development | 0.181547791 | 0.539299215 |
| GO_erythrocyte_differentiation | 0.884090459 | 0.723010954 |
| GO_erythrocyte_homeostasis | 0.905827262 | 0.708176126 |
| GO_erythrocyte_maturation | 0.027250593 | 0.38536015 |
| GO_establishment_of_lymphocyte_polarity | 0.037316169 | 0.275279593 |
| GO_establishment_of_T_cell_polarity | 0.028778098 | 0.308750114 |
| GO_Fc_receptor_signaling_pathway | 2.12E-06 | 3.73E-05 |
| GO_Fc-gamma_receptor_signaling_pathway | 3.46E-08 | 1.65E-05 |
| GO_gamma-delta_T_cell_activation | 0.592356319 | 0.336146501 |
| GO_gamma-delta_T_cell_differentiation | 0.209499189 | 0.734822114 |
| GO_germinal_center_formation | 0.379165245 | 0.239252823 |
| GO_granulocyte_activation | 0.168884694 | 0.077636201 |
| GO_granulocyte_chemotaxis | 0.884588689 | 0.825656881 |
| GO_granulocyte_differentiation | 0.89191947 | 0.939498294 |
| GO_granulocyte_migration | 0.61621216 | 0.950296855 |
| GO_hemopoiesis | 0.782812351 | 0.847941212 |
| GO_histamine_secretion_by_mast_cell | 0.600362156 | 0.353413324 |
| GO_humoral_immune_response | 1.59E-06 | 0.007568843 |
| GO_hypersensitivity | 0.046449486 | 0.220325311 |
| GO_immature_B_cell_differentiation | 0.180123962 | 0.400607889 |
| GO_immature_T_cell_proliferation | 0.43364825 | 0.370179904 |
| GO_immune_effector_process | 0.054089604 | 0.025894741 |
| GO_immune_response | 0.057984065 | 0.017207129 |
| GO_immune_response_to_tumor_cell | 0.017464965 | 0.247517164 |
| GO_immune_system_development | 0.752879066 | 0.849808823 |
| GO_immunoglobulin_biosynthetic_process | 0.002633362 | 0.00761367 |
| GO_immunoglobulin_production | 3.83E-06 | 0.000694452 |
| GO_immunoglobulin_secretion | 0.184402756 | 0.331596241 |
| GO_immunoglobulin_V(D)J_recombination | 0.586738534 | 0.318216224 |
| GO_immunological_memory_process | 0.686683046 | 0.970736089 |
| GO_immunological_synapse_formation | 0.181659292 | 0.089672594 |
| GO_induction_of_bacterial_agglutination | 0.079917372 | 0.377348427 |
| GO_innate_immune_response | 0.002075282 | 0.184829733 |
| GO_innate_immune_response_in_mucosa | 0.246925326 | 0.414281588 |
| GO_isotype_switching | 0.935738648 | 0.87102227 |
| GO_isotype_switching_to_IgA_isotypes | 0.932936404 | 0.939162224 |
| GO_isotype_switching_to_IgE_isotypes | 0.040552095 | 0.72316054 |
| GO_isotype_switching_to_IgG_isotypes | 0.935050832 | 0.978392087 |
| GO_leukocyte_activation | 0.064114691 | 0.031301244 |
| GO_leukocyte_chemotaxis | 0.979732374 | 0.861140775 |

| | | |
|---|---|---|
| GO_leukocyte_degranulation | 0.188441372 | 0.089432595 |
| GO_leukocyte_differentiation | 0.83384134 | 0.550017819 |
| GO_leukocyte_homeostasis | 0.211422248 | 0.099858057 |
| GO_leukocyte_mediated_cytotoxicity | 0.269841471 | 0.201554601 |
| GO_leukocyte_mediated_immunity | 0.00132105 | 0.00187939 |
| GO_leukocyte_migration | 0.0001699 | 0.026150581 |
| GO_leukocyte_tethering_or_rolling | 0.159582179 | 0.595428783 |
| GO_lymph_node_development | 0.056558014 | 0.258646621 |
| GO_lymphocyte_activation | 0.018594503 | 0.00883176 |
| GO_lymphocyte_anergy | 0.171408249 | 0.446497966 |
| GO_lymphocyte_chemotaxis | 0.148917167 | 0.233971981 |
| GO_lymphocyte_costimulation | 0.00562331 | 0.152721721 |
| GO_lymphocyte_differentiation | 0.624315535 | 0.358540243 |
| GO_lymphocyte_homeostasis | 0.15526605 | 0.101580701 |
| GO_lymphocyte_mediated_immunity | 3.96E-06 | 6.04E-05 |
| GO_lymphocyte_migration | 0.284491822 | 0.320595512 |
| GO_lymphocyte_proliferation | 0.449497156 | 0.884917187 |
| GO_macrophage_activation | 0.311837782 | 0.294491211 |
| GO_macrophage_chemotaxis | 0.678891334 | 0.528101091 |
| GO_macrophage_cytokine_production | 0.336520133 | 0.84869409 |
| GO_macrophage_differentiation | 0.49615987 | 0.906685201 |
| GO_macrophage_migration | 0.856328937 | 0.705224248 |
| GO_marginal_zone_B_cell_differentiation | 0.436065829 | 0.255659405 |
| GO_mast_cell_activation | 0.982282343 | 0.96240271 |
| GO_mast_cell_chemotaxis | 0.01551945 | 0.942365445 |
| GO_mast_cell_cytokine_production | 0.486927296 | 0.676150864 |
| GO_mast_cell_degranulation | 0.882881545 | 0.73913755 |
| GO_mast_cell_differentiation | 0.039188246 | 0.802010268 |
| GO_mast_cell_mediated_immunity | 0.866814832 | 0.614609576 |
| GO_mast_cell_migration | 0.01551945 | 0.942365445 |
| GO_mature_B_cell_differentiation | 0.744955622 | 0.51610356 |
| GO_MDA-5_signaling_pathway | 5.68E-10 | 0.531222341 |
| GO_megakaryocyte_development | 0.229573692 | 0.5601489 |
| GO_megakaryocyte_differentiation | 0.087974739 | 0.697967881 |
| GO_memory_T_cell_differentiation | 0.369275741 | 0.791029416 |
| GO_microglia_differentiation | 0.070177094 | 0.081840783 |
| GO_microglial_cell_activation | 0.430060583 | 0.208248607 |
| GO_microglial_cell_migration | 0.017859668 | 0.574941116 |
| GO_monocyte_activation | 0.361631982 | 0.273279642 |
| GO_monocyte_chemotaxis | 0.430923576 | 0.320279712 |
| GO_monocyte_differentiation | 0.47716424 | 0.845264155 |
| GO_monocyte_extravasation | 0.048162139 | 0.245916753 |

| | | |
|---|---|---|
| GO_mononuclear_cell_differentiation | 0.47716424 | 0.845264155 |
| GO_mononuclear_cell_migration | 0.600847098 | 0.552411972 |
| GO_mucosal_immune_response | 0.590408187 | 0.881807117 |
| GO_myeloid_cell_development | 0.166304562 | 0.968674941 |
| GO_myeloid_cell_differentiation | 0.740458567 | 0.851223417 |
| GO_myeloid_cell_homeostasis | 0.978792404 | 0.969245053 |
| GO_myeloid_dendritic_cell_activation | 0.068485465 | 0.150651417 |
| GO_myeloid_dendritic_cell_chemotaxis | 0.049753898 | 0.186748516 |
| GO_myeloid_leukocyte_activation | 0.22628582 | 0.102974271 |
| GO_myeloid_leukocyte_differentiation | 0.879164525 | 0.765726101 |
| GO_myeloid_leukocyte_mediated_immunity | 0.172186627 | 0.080642767 |
| GO_myeloid_leukocyte_migration | 0.705188553 | 0.921513858 |
| GO_natural_killer_cell_activation | 0.762269704 | 0.866598339 |
| GO_natural_killer_cell_chemotaxis | 0.008891326 | 0.796473241 |
| GO_natural_killer_cell_degranulation | 0.199253903 | 0.108183452 |
| GO_natural_killer_cell_differentiation | 0.7111982 | 0.533645222 |
| GO_natural_killer_cell_proliferation | 0.146578979 | 0.483034654 |
| GO_negative_T_cell_selection | 0.856803258 | 0.584138785 |
| GO_negative_thymic_T_cell_selection | 0.86839347 | 0.596989532 |
| GO_neutrophil_activation | 0.167751388 | 0.077107178 |
| GO_neutrophil_apoptotic_process | 0.573890595 | 0.308388594 |
| GO_neutrophil_chemotaxis | 0.939952568 | 0.793840005 |
| GO_neutrophil_clearance | 0.026393933 | 0.691209695 |
| GO_neutrophil_degranulation | 0.157819223 | 0.076962156 |
| GO_neutrophil_differentiation | 0.053068741 | 0.021771301 |
| GO_neutrophil_extravasation | 0.055581048 | 0.360534599 |
| GO_neutrophil_homeostasis | 0.464887279 | 0.217486649 |
| GO_neutrophil_mediated_cytotoxicity | 0.013899567 | 0.496413374 |
| GO_neutrophil_mediated_immunity | 0.148788824 | 0.078620598 |
| GO_neutrophil_migration | 0.705173441 | 0.986454451 |
| GO_NK_T_cell_activation | 0.038066671 | 0.260018472 |
| GO_NK_T_cell_differentiation | 0.020012534 | 0.013373293 |
| GO_NK_T_cell_proliferation | 0.000752873 | 0.419670956 |
| GO_opsonization | 0.136886271 | 0.17575026 |
| GO_osteoclast_development | 0.404734856 | 0.421359568 |
| GO_osteoclast_differentiation | 0.906519634 | 0.859812045 |
| GO_osteoclast_fusion | 0.983005416 | 0.902577736 |
| GO_peripheral_tolerance_induction | 0.00108049 | 0.467750927 |
| GO_Peyer's_patch_development | 0.453654819 | 0.553070837 |
| GO_platelet_formation | 0.077388301 | 0.305644521 |
| GO_positive_T_cell_selection | 0.803456813 | 0.612673515 |
| GO_positive_thymic_T_cell_selection | 0.854496474 | 0.773854134 |

| | | |
|---|---|---|
| GO_pre-B_cell_differentiation | 0.428161444 | 0.277993382 |
| GO_primitive_hemopoiesis | 0.273436817 | 0.530304086 |
| GO_pro-B_cell_differentiation | 0.008420049 | 0.133128589 |
| GO_regulatory_T_cell_differentiation | 0.017986512 | 0.552855942 |
| GO_response_to_interferon-gamma | 3.98E-05 | 0.516768435 |
| GO_response_to_type_I_interferon | 1.39E-08 | 0.372165051 |
| GO_RIG-I_signaling_pathway | 1.15E-08 | 0.914911182 |
| GO_spleen_development | 0.003723017 | 0.231102065 |
| GO_T_cell_activation | 0.2200628 | 0.285031387 |
| GO_T_cell_anergy | 0.171408249 | 0.446497966 |
| GO_T_cell_chemotaxis | 0.005807448 | 0.346210188 |
| GO_T_cell_costimulation | 0.005818178 | 0.165930083 |
| GO_T_cell_cytokine_production | 0.003551284 | 0.323293179 |
| GO_T_cell_differentiation | 0.249027089 | 0.270273795 |
| GO_T_cell_differentiation_in_thymus | 0.795994823 | 0.520493227 |
| GO_T_cell_extravasation | 0.191483346 | 0.265317823 |
| GO_T_cell_homeostasis | 0.806369752 | 0.726047441 |
| GO_T_cell_lineage_commitment | 0.39498853 | 0.769173736 |
| GO_T_cell_mediated_cytotoxicity | 0.2769928 | 0.280559767 |
| GO_T_cell_mediated_immunity | 0.021501205 | 0.169499607 |
| GO_T_cell_migration | 0.115890652 | 0.722850137 |
| GO_T_cell_proliferation | 0.287834558 | 0.906768255 |
| GO_T_cell_receptor_signaling_pathway | 0.093936666 | 0.404614687 |
| GO_T_cell_receptor_V(D)J_recombination | 0.682702893 | 0.758343136 |
| GO_T_cell_selection | 0.945618894 | 0.804936245 |
| GO_T_cell_tolerance_induction | 0.900285138 | 0.651970858 |
| GO_T-helper_1_cell_cytokine_production | 0.636170044 | 0.941888544 |
| GO_T-helper_1_cell_differentiation | 0.966236032 | 0.793592319 |
| GO_T-helper_1_type_immune_response | 0.820291295 | 0.531385854 |
| GO_T-helper_17_cell_differentiation | 0.219589748 | 0.489518409 |
| GO_T-helper_17_cell_lineage_commitment | 0.749687713 | 0.448585087 |
| GO_T-helper_17_type_immune_response | 0.246359507 | 0.505589914 |
| GO_T-helper_2_cell_cytokine_production | 2.83E-07 | 0.702248666 |
| GO_T-helper_2_cell_differentiation | 0.672701889 | 0.482278613 |
| GO_T-helper_cell_differentiation | 0.496740179 | 0.308480472 |
| GO_T-helper_cell_lineage_commitment | 0.399822895 | 0.242918928 |
| GO_thymic_T_cell_selection | 0.840787189 | 0.659725707 |
| GO_thymocyte_migration | 0.436432476 | 0.204570234 |
| GO_thymus_development | 0.029036109 | 0.86658034 |
| GO_tolerance_induction | 0.012637662 | 0.17272127 |
| GO_tolerance_induction_to_self_antigen | 0.526603281 | 0.45618318 |
| GO_type_2_immune_response | 0.042021765 | 0.34489055 |

| | | |
|---|---|---|
| GO_type_I_interferon_signaling_pathway | 2.53E-08 | 0.352675627 |
| GO_V(D)J_recombination | 0.728956932 | 0.961062366 |
| olink_IL8_P10145_OID00471 | 0.166268238 | 0.334906454 |
| olink_VEGFA_P15692_OID00472 | 0.430429378 | 0.955685697 |
| olink_CD8A_P01732_OID05124 | 0.675840617 | 0.560303079 |
| olink_MCP-3_P80098_OID00474 | 0.368839352 | 0.235317631 |
| olink_GDNF_P39905_OID00475 | 0.544062073 | 0.392438147 |
| olink_CDCP1_Q9H5V8_OID00476 | 0.762794246 | 0.970322982 |
| olink_CD244_Q9BZW8_OID00477 | 0.061764452 | 0.056729831 |
| olink_IL7_P13232_OID00478 | 0.879623964 | 0.66277487 |
| olink_OPG_O00300_OID00479 | 0.012237651 | 0.529687292 |
| olink_LAP TGF-beta-1_P01137_OID00480 | 0.178731623 | 0.086617234 |
| olink_uPA_P00749_OID00481 | 0.477290333 | 0.389335488 |
| olink_IL6_P05231_OID00482 | 0.024021821 | 0.057056853 |
| olink_IL-17C_Q9P0M4_OID00483 | 0.937922005 | 0.730202298 |
| olink_MCP-1_P13500_OID00484 | 0.001475547 | 0.679450866 |
| olink_IL-17A_Q16552_OID00485 | 0.805397741 | 0.518940326 |
| olink_CXCL11_O14625_OID00486 | 7.02E-06 | 0.886354547 |
| olink_AXIN1_O15169_OID00487 | 0.326425797 | 0.923590993 |
| olink_TRAIL_P50591_OID00488 | 0.002557208 | 0.129821827 |
| olink_IL-20RA_Q9UHF4_OID00489 | 0.065492839 | 0.091933447 |
| olink_CXCL9_Q07325_OID00490 | 0.55054001 | 0.695890712 |
| olink_CST5_P28325_OID00491 | 0.323143322 | 0.617587061 |
| olink_IL-2RB_P14784_OID00492 | 0.268291685 | 0.161705392 |
| olink_IL-1 alpha_P01583_OID00493 | 0.857506708 | 0.898300404 |
| olink_OSM_P13725_OID00494 | 0.774001471 | 0.530922688 |
| olink_IL2_P60568_OID00495 | 0.721741359 | 0.421027914 |
| olink_CXCL1_P09341_OID00496 | 0.000312155 | 0.042128918 |
| olink_TSLP_Q969D9_OID00497 | 0.95124218 | 0.767364553 |
| olink_CCL4_P13236_OID00498 | 0.71049609 | 0.540881146 |
| olink_CD6_P30203_OID00499 | 0.644900282 | 0.349842928 |
| olink_SCF_P21583_OID00500 | 0.006755758 | 0.231914293 |
| olink_IL18_Q14116_OID00501 | 0.736456849 | 0.919324568 |
| olink_SLAMF1_Q13291_OID00502 | 0.48234082 | 0.569328595 |
| olink_TGF-alpha_P01135_OID00503 | 0.353546809 | 0.920223649 |
| olink_MCP-4_Q99616_OID00504 | 0.616526521 | 0.43346188 |
| olink_CCL11_P51671_OID00505 | 0.54364976 | 0.701177288 |
| olink_TNFSF14_O43557_OID00506 | 0.817479309 | 0.715820603 |
| olink_FGF-23_Q9GZV9_OID00507 | 0.077185386 | 0.042086963 |
| olink_IL-10RA_Q13651_OID00508 | 0.106815493 | 0.207077701 |
| olink_FGF-5_P12034_OID00509 | 0.622050805 | 0.85453822 |
| olink_MMP-1_P03956_OID00510 | 0.295723115 | 0.372677365 |

| | | |
|---|---|---|
| olink_LIF-R_P42702_OID00511 | 0.640887827 | 0.352871415 |
| olink_FGF-21_Q9NSA1_OID00512 | 0.455232811 | 0.667141519 |
| olink_CCL19_Q99731_OID00513 | 0.003559047 | 0.951515113 |
| olink_IL-15RA_Q13261_OID00514 | 0.115203218 | 0.769068278 |
| olink_IL-10RB_Q08334_OID00515 | 0.043505835 | 0.060492535 |
| olink_IL-22 RA1_Q8N6P7_OID00516 | 0.559682183 | 0.659882707 |
| olink_IL-18R1_Q13478_OID00517 | 0.917238582 | 0.951055773 |
| olink_PD-L1_Q9NZQ7_OID00518 | 0.874940215 | 0.745774059 |
| olink_Beta-NGF_P01138_OID00519 | 0.971634034 | 0.853220635 |
| olink_CXCL5_P42830_OID00520 | 0.033499855 | 0.140794607 |
| olink_TRANCE_O14788_OID00521 | 0.20651343 | 0.710177566 |
| olink_HGF_P14210_OID00522 | 0.906545581 | 0.661007275 |
| olink_IL-12B_P29460_OID00523 | 0.379511238 | 0.199110137 |
| olink_IL-24_Q13007_OID00524 | 0.631359626 | 0.462522175 |
| olink_IL13_P35225_OID00525 | 0.8829249 | 0.698229556 |
| olink_ARTN_Q5T4W7_OID00526 | 0.816490467 | 0.675366254 |
| olink_MMP-10_P09238_OID00527 | 0.863691617 | 0.62743781 |
| olink_IL10_P22301_OID00528 | 0.182864366 | 0.686812363 |
| olink_TNF_P01375_OID05548 | 0.301711662 | 0.931739314 |
| olink_CCL23_P55773_OID00530 | 0.926397522 | 0.751249183 |
| olink_CD5_P06127_OID00531 | 0.012607917 | 0.474124869 |
| olink_CCL3_P10147_OID00532 | 0.838979838 | 0.684834366 |
| olink_Flt3L_P49771_OID00533 | 0.022668577 | 0.109545764 |
| olink_CXCL6_P80162_OID00534 | 0.000149859 | 0.050819019 |
| olink_CXCL10_P02778_OID00535 | 6.70E-05 | 0.522088658 |
| olink_4E-BP1_Q13541_OID00536 | 0.571207517 | 0.558228684 |
| olink_IL-20_Q9NYY1_OID00537 | 0.978364013 | 0.845505131 |
| olink_SIRT2_Q8IXJ6_OID00538 | 0.542850684 | 0.720278146 |
| olink_CCL28_Q9NRJ3_OID00539 | 0.8672411 | 0.937612175 |
| olink_DNER_Q8NFT8_OID01213 | 0.487114499 | 0.669581437 |
| olink_EN-RAGE_P80511_OID00541 | 0.713163682 | 0.81748453 |
| olink_CD40_P25942_OID00542 | 0.858141776 | 0.580952953 |
| olink_IL33_O95760_OID00543 | 0.520819107 | 0.507822617 |
| olink_IFN-gamma_P01579_OID05547 | 0.00085468 | 0.100609212 |
| olink_FGF-19_O95750_OID00545 | 0.606853163 | 0.513462455 |
| olink_IL4_P05112_OID00546 | 0.307333256 | 0.308930293 |
| olink_LIF_P15018_OID00547 | 0.846089198 | 0.570080367 |
| olink_NRTN_Q99748_OID00548 | 0.658573579 | 0.744983817 |
| olink_MCP-2_P80075_OID00549 | 1.02E-07 | 0.88077081 |
| olink_CASP-8_Q14790_OID00550 | 0.835861189 | 0.613755951 |
| olink_CCL25_O15444_OID00551 | 0.069822452 | 0.021622099 |
| olink_CX3CL1_P78423_OID00552 | 0.217114613 | 0.303906725 |

| | | |
|---|---|---|
| olink_TNFRSF9_Q07011_OID00553 | 0.023393906 | 0.165283623 |
| olink_NT-3_P20783_OID00554 | 0.801075058 | 0.507431105 |
| olink_TWEAK_O43508_OID00555 | 0.110217872 | 0.253995247 |
| olink_CCL20_P78556_OID00556 | 0.557485531 | 0.2894123 |
| olink_ST1A1_P50225_OID00557 | 0.4517008 | 0.574936782 |
| olink_STAMBP_O95630_OID00558 | 0.777398838 | 0.885220743 |
| olink_IL5_P05113_OID00559 | 0.694875704 | 0.481391149 |
| olink_ADA_P00813_OID00560 | 0.88246517 | 0.978312175 |
| olink_TNFB_P01374_OID00561 | 0.292192822 | 0.861660448 |
| olink_CSF-1_P09603_OID00562 | 0.069603162 | 0.941452234 |
| olink_PPP1R9B_Q96SB3_OID00936 | 0.956281833 | 0.832772038 |
| olink_GLB1_P16278_OID00937 | 0.748566883 | 0.467328311 |
| olink_PSIP1_O75475_OID00938 | 0.53913263 | 0.39430347 |
| olink_ZBTB16_Q05516_OID00939 | 0.468579333 | 0.218958143 |
| olink_IRAK4_Q9NWZ3_OID00940 | 0.498454971 | 0.349361178 |
| olink_TPSAB1_Q15661_OID00941 | 0.570671718 | 0.315319151 |
| olink_HCLS1_P14317_OID00942 | 0.943475291 | 0.741672417 |
| olink_CNTNAP2_Q9UHC6_OID00943 | 0.110724489 | 0.166143817 |
| olink_CLEC4G_Q6UXB4_OID00944 | 0.272356463 | 0.568174439 |
| olink_IRF9_Q00978_OID00945 | 0.628385176 | 0.87123383 |
| olink_EDAR_Q9UNE0_OID00946 | 0.693260013 | 0.393445519 |
| olink_IL6_P05231_OID00947 | 0.023055944 | 0.045737019 |
| olink_DGKZ_Q13574_OID00948 | 0.107317587 | 0.138410963 |
| olink_CLEC4C_Q8WTT0_OID00949 | 0.181123289 | 0.354264217 |
| olink_IRAK1_P51617_OID00950 | 0.612600394 | 0.610388137 |
| olink_CLEC4A_Q9UMR7_OID00951 | 0.000530583 | 0.001772081 |
| olink_PRDX1_Q06830_OID00952 | 0.758774899 | 0.547887314 |
| olink_PRDX3_P30048_OID00953 | 0.19572631 | 0.529154557 |
| olink_FGF2_P09038_OID00954 | 0.374169013 | 0.999930592 |
| olink_PRDX5_P30044_OID00955 | 0.703323706 | 0.405848622 |
| olink_DPP10_Q8N608_OID00956 | 0.542587741 | 0.642714136 |
| olink_TRIM5_Q9C035_OID00957 | 0.720948543 | 0.799240372 |
| olink_DCTN1_Q14203_OID00958 | 0.416029021 | 0.257163081 |
| olink_ITGA6_P23229_OID00959 | 0.182683047 | 0.549002503 |
| olink_CDSN_Q15517_OID00960 | 0.319392006 | 0.250073748 |
| olink_GALNT3_Q14435_OID00961 | 0.910175336 | 0.991870995 |
| olink_FXYD5_Q96DB9_OID00962 | 0.863237716 | 0.700802062 |
| olink_TRAF2_Q12933_OID00963 | 0.662885981 | 0.405849109 |
| olink_TRIM21_P19474_OID00964 | 0.512480501 | 0.693586489 |
| olink_LILRB4_Q8NHJ6_OID00965 | 0.739067211 | 0.854478892 |
| olink_NTF4_P34130_OID00966 | 0.720029201 | 0.984813049 |
| olink_KRT19_P08727_OID00967 | 0.428124344 | 0.519449079 |

| | | |
|---|---|---|
| olink_ITM2A_O43736_OID00968 | 0.1672493 | 0.182958024 |
| olink_HNMT_P50135_OID00969 | 0.622820863 | 0.557879785 |
| olink_CCL11_P51671_OID00970 | 0.959094435 | 0.956712495 |
| olink_MILR1_Q7Z6M3_OID00971 | 0.189897843 | 0.43361069 |
| olink_EGLN1_Q9GZT9_OID00972 | 0.012824005 | 0.127016628 |
| olink_NFATC3_Q12968_OID00973 | 0.507742347 | 0.421219696 |
| olink_LY75_O60449_OID00974 | 0.604649585 | 0.768083437 |
| olink_EIF5A_P63241_OID00975 | 0.078667021 | 0.156484157 |
| olink_EIF4G1_Q04637_OID00976 | 0.89349573 | 0.644539552 |
| olink_CD28_P10747_OID00977 | 0.701966269 | 0.900164794 |
| olink_PTH1R_Q03431_OID00978 | 0.175850738 | 0.822508388 |
| olink_BIRC2_Q13490_OID00979 | 0.18336303 | 0.673600577 |
| olink_HSD11B1_P28845_OID00980 | 0.679586179 | 0.390265907 |
| olink_NF2_P35240_OID00981 | 0.608460645 | 0.917552002 |
| olink_PLXNA4_Q9HCM2_OID00982 | 0.111676145 | 0.039471854 |
| olink_SH2B3_Q9UQQ2_OID00983 | 0.371901369 | 0.264591971 |
| olink_FCRL3_Q96P31_OID00984 | 0.586085421 | 0.591866802 |
| olink_CKAP4_Q07065_OID00985 | 0.995902533 | 0.989192939 |
| olink_JUN_P05412_OID00986 | 0.240366187 | 0.15640872 |
| olink_HEXIM1_O94992_OID00987 | 0.545406284 | 0.646700083 |
| olink_CLEC4D_Q8WXI8_OID00988 | 0.599031065 | 0.650738715 |
| olink_PRKCQ_Q04759_OID00989 | 0.238897826 | 0.229988977 |
| olink_MGMT_P16455_OID00990 | 0.47391226 | 0.869227458 |
| olink_TREM1_Q9NP99_OID00991 | 0.006060195 | 0.170630339 |
| olink_CXADR_P78310_OID00992 | 0.539814611 | 0.467665194 |
| olink_IL10_P22301_OID00993 | 0.089979921 | 0.170949665 |
| olink_SRPK2_P78362_OID00994 | 0.06277937 | 0.208517952 |
| olink_KLRD1_Q13241_OID00995 | 0.303501929 | 0.623980429 |
| olink_BACH1_O14867_OID00996 | 0.389132862 | 0.246253467 |
| olink_PIK3AP1_Q6ZUJ8_OID00997 | 0.745643298 | 0.882878363 |
| olink_SPRY2_O43597_OID00998 | 0.386985007 | 0.953465154 |
| olink_STC1_P52823_OID00999 | 0.538761143 | 0.766814588 |
| olink_ARNT_P27540_OID01000 | 0.518500154 | 0.511013309 |
| olink_FAM3B_P58499_OID01001 | 0.163492563 | 0.087340817 |
| olink_SH2D1A_O60880_OID01002 | 0.000152907 | 0.001344289 |
| olink_ICA1_Q05084_OID01003 | 0.369069385 | 0.187127129 |
| olink_DFFA_O00273_OID01004 | 0.995877564 | 0.933118418 |
| olink_DCBLD2_Q96PD2_OID01005 | 0.063999586 | 0.997443036 |
| olink_FCRL6_Q6DN72_OID01006 | 0.019011547 | 0.041128108 |
| olink_NCR1_O76036_OID01007 | 0.165915989 | 0.77136478 |
| olink_CXCL12_P48061_OID01008 | 0.007477298 | 0.083974837 |
| olink_AREG_P15514_OID01009 | 0.355979581 | 0.663274688 |

| | | |
|---|---|---|
| olink_IFNLR1_Q8IU57_OID01010 | 0.168499268 | 0.779232935 |
| olink_DAPP1_Q9UN19_OID01011 | 0.887390656 | 0.626059098 |
| olink_PADI2_Q9Y2J8_OID01012 | 0.132564682 | 0.263841884 |
| olink_SIT1_Q9Y3P8_OID01013 | 0.013313387 | 0.223286855 |
| olink_MASP1_P48740_OID01014 | 0.093711489 | 0.108121731 |
| olink_LAMP3_Q9UQV4_OID01015 | 0.685922779 | 0.393643561 |
| olink_CLEC7A_Q9BXN2_OID01016 | 0.825933918 | 0.5371622 |
| olink_CLEC6A_Q6EIG7_OID01017 | 0.030550322 | 0.014832767 |
| olink_DDX58_O95786_OID01018 | 0.596665175 | 0.482758574 |
| olink_IL12RB1_P42701_OID01019 | 0.617255107 | 0.824200615 |
| olink_TANK_Q92844_OID01020 | 0.499210585 | 0.96723725 |
| olink_ITGA11_Q9UKX5_OID01021 | 0.017548487 | 0.269847266 |
| olink_KPNA1_P52294_OID01022 | 0.342840011 | 0.174239313 |
| olink_LAG3_P18627_OID01023 | 0.234917328 | 0.362283005 |
| olink_IL5_P05113_OID01024 | 0.525705116 | 0.277511408 |
| olink_CD83_Q01151_OID01025 | 0.0711711 | 0.181019342 |
| olink_ITGB6_P18564_OID01026 | 0.200426127 | 0.144906562 |
| olink_BTN3A2_P78410_OID01027 | 0.096497802 | 0.17919958 |
| IgG_antibody_value | 1.03E-13 | 0.411870531 |

| overall_FDR_linear_regression | overall_p_value_mixed_model | overall_FDR_mixed_model |
|---|---|---|
| 0.951519517 | 0.59588626 | 0.72506903 |
| 0.000343355 | 1.16E-06 | 1.23E-05 |
| 0.200620906 | 0.04152874 | 0.11150046 |
| 0.000573169 | 3.46E-06 | 3.35E-05 |
| 0.01634511 | 0.00243306 | 0.01085273 |
| 0.056608409 | 0.00937617 | 0.03454916 |
| 0.446990037 | 0.29393946 | 0.44090918 |
| 0.021028187 | 0.00082709 | 0.00445053 |
| 0.002339659 | 4.06E-11 | 1.53E-09 |
| 0.089599127 | 0.0224848 | 0.0699298 |
| 0.00906382 | 1.44E-06 | 1.47E-05 |
| 0.141350017 | 0.04999915 | 0.12372052 |
| 0.119270965 | 0.02518063 | 0.07621637 |
| 0.057122233 | 6.17E-06 | 5.51E-05 |
| 0.089599127 | 0.0224848 | 0.0699298 |
| 0.000134386 | 2.50E-08 | 3.69E-07 |
| 0.125287393 | 0.03809189 | 0.10518409 |
| 0.009317912 | 0.00372563 | 0.01574808 |
| 0.028261337 | 0.004213 | 0.01732463 |
| 0.278630716 | 0.1407229 | 0.26502812 |
| 3.34E-06 | 1.88E-11 | 1.44E-09 |
| 0.763084314 | 0.25662528 | 0.40275912 |
| 9.20E-06 | 2.12E-11 | 1.44E-09 |
| 0.538571622 | 0.3011338 | 0.44773841 |
| 0.059688319 | 0.01025268 | 0.03737266 |
| 0.22543742 | 0.12998065 | 0.25349234 |
| 5.45E-06 | 2.89E-11 | 1.53E-09 |
| 8.10E-07 | 3.62E-09 | 7.67E-08 |
| 0.286828432 | 0.04177156 | 0.11150046 |
| 0.32445058 | 0.08704238 | 0.19160628 |
| 0.000824399 | 6.41E-05 | 0.0004625 |
| 0.640870157 | 0.08910856 | 0.19323229 |
| 0.331606887 | 0.0980658 | 0.20648637 |
| 0.424222026 | 0.39420217 | 0.5375723 |
| 0.242855108 | 0.06479652 | 0.14894297 |
| 0.374173874 | 0.16168409 | 0.28399433 |
| 0.140337513 | 0.04742603 | 0.11998078 |
| 0.393819014 | 0.16931411 | 0.28842956 |
| 0.511694402 | 0.37276196 | 0.51368416 |

| | | |
|---|---|---|
| 0.393819014 | 0.16931411 | 0.28842956 |
| 0.206583765 | 0.09292147 | 0.19936949 |
| 0.36013158 | 0.11345135 | 0.23029945 |
| 0.948973308 | 0.88427142 | 0.93095656 |
| 0.957352511 | 0.94339303 | 0.9635595 |
| 0.09724159 | 0.01063161 | 0.03793806 |
| 0.12524932 | 0.04559036 | 0.11620401 |
| 0.10147671 | 0.03206055 | 0.09133215 |
| 3.26E-05 | 1.65E-07 | 1.99E-06 |
| 1.13E-06 | 1.16E-10 | 3.92E-09 |
| 0.760639365 | 0.34918157 | 0.49321896 |
| 0.385334097 | 0.53823422 | 0.66591752 |
| 0.561297022 | 0.29312789 | 0.44090918 |
| 0.355922358 | 0.14495458 | 0.26706305 |
| 0.948973308 | 0.98951314 | 0.98951314 |
| 0.951519517 | 0.63681554 | 0.74958496 |
| 0.779462396 | 0.73942934 | 0.82956991 |
| 0.890514721 | 0.9367306 | 0.9635595 |
| 0.763084314 | 0.57109893 | 0.69892613 |
| 2.66E-05 | 1.50E-10 | 4.63E-09 |
| 0.141859241 | 0.00149644 | 0.00724704 |
| 0.36013158 | 0.2599315 | 0.40477685 |
| 0.611366155 | 0.4530163 | 0.58840048 |
| 0.158072205 | 0.03698071 | 0.10360712 |
| 0.163856627 | 0.0493402 | 0.12306358 |
| 0.071332807 | 0.00189221 | 0.0087871 |
| 0.865172892 | 0.93674329 | 0.9635595 |
| 0.015941247 | 0.00070956 | 0.00394327 |
| 5.17E-05 | 3.07E-08 | 4.34E-07 |
| 0.363444968 | 0.04353225 | 0.11265216 |
| 0.760639365 | 0.5474625 | 0.67487195 |
| 0.825480684 | 0.61700711 | 0.74389166 |
| 0.36013158 | 0.15958538 | 0.28328696 |
| 0.206809077 | 0.09578822 | 0.2029513 |
| 0.012791283 | 0.00057702 | 0.00343177 |
| 0.424222026 | 0.00014302 | 0.00098944 |
| 0.967120127 | 0.6270493 | 0.74389166 |
| 0.967120127 | 0.85899194 | 0.91860653 |
| 0.130925334 | 0.00329566 | 0.01414212 |
| 0.967120127 | 0.83411089 | 0.89482149 |
| 0.178154756 | 0.06032674 | 0.14103975 |
| 0.988098613 | 0.80304183 | 0.8781651 |

| | | |
|---|---|---|
| 0.369257948 | 0.16421193 | 0.28694765 |
| 0.926794145 | 0.62759002 | 0.74389166 |
| 0.385334097 | 0.18571957 | 0.30869789 |
| 0.446990037 | 0.35862415 | 0.50237019 |
| 0.008956721 | 0.00068468 | 0.00386844 |
| 0.001515688 | 5.25E-06 | 4.95E-05 |
| 0.346870502 | 0.36378426 | 0.5075015 |
| 0.162484464 | 0.01806389 | 0.059453 |
| 0.073296936 | 0.0194824 | 0.06230691 |
| 0.355922358 | 0.03847442 | 0.10518409 |
| 0.334323971 | 0.04937064 | 0.12306358 |
| 0.029785969 | 0.00135192 | 0.0067397 |
| 0.786776827 | 0.60472254 | 0.73214621 |
| 0.345348521 | 0.14313106 | 0.26691984 |
| 5.17E-05 | 1.16E-08 | 1.97E-07 |
| 0.452782759 | 0.08796044 | 0.192378 |
| 0.624506294 | 0.14408947 | 0.26691984 |
| 0.489412075 | 0.44859401 | 0.58516489 |
| 0.823616657 | 0.21754452 | 0.347866 |
| 0.518546932 | 0.44836946 | 0.58516489 |
| 0.665592571 | 0.82567122 | 0.89140937 |
| 0.936955821 | 0.43898293 | 0.57904752 |
| 0.611366155 | 0.41392133 | 0.55595625 |
| 0.988098613 | 0.7439211 | 0.82956991 |
| 0.065240924 | 0.04300373 | 0.1121405 |
| 0.657642842 | 0.28455189 | 0.4306388 |
| 0.948973308 | 0.87217331 | 0.92648659 |
| 0.127738611 | 0.01503781 | 0.05097817 |
| 0.943542905 | 0.81110131 | 0.87847714 |
| 0.065240924 | 0.04300373 | 0.1121405 |
| 0.861911112 | 0.79194642 | 0.86883443 |
| 1.92E-07 | 2.69E-10 | 7.59E-09 |
| 0.399105034 | 0.21935801 | 0.34911908 |
| 0.224236366 | 0.02828682 | 0.08411606 |
| 0.549054721 | 0.47475105 | 0.61194147 |
| 0.188809799 | 0.0765542 | 0.17073602 |
| 0.611226327 | 0.39643975 | 0.5375723 |
| 0.071734441 | 0.008376 | 0.0312029 |
| 0.544858852 | 0.53145396 | 0.66480772 |
| 0.611226327 | 0.14030853 | 0.26502812 |
| 0.647034709 | 0.94366299 | 0.9635595 |
| 0.145776474 | 0.06502525 | 0.14894297 |

| | | |
|---|---|---|
| 0.647034709 | 0.94366299 | 0.9635595 |
| 0.763084314 | 0.10048354 | 0.2089811 |
| 0.760639365 | 0.05063201 | 0.12437862 |
| 0.355922358 | 0.15784846 | 0.28312501 |
| 0.861911112 | 0.80982464 | 0.87847714 |
| 0.988098613 | 0.92495543 | 0.9635595 |
| 0.185732581 | 0.05726002 | 0.13574228 |
| 0.149261694 | 0.01693489 | 0.05651568 |
| 0.395416973 | 0.19978519 | 0.32332571 |
| 0.948973308 | 0.80868245 | 0.87847714 |
| 0.355922358 | 0.15169547 | 0.27499874 |
| 0.832625319 | 0.94225332 | 0.9635595 |
| 0.872552572 | 0.22608397 | 0.35814237 |
| 0.042864265 | 0.00257787 | 0.01120384 |
| 0.379477939 | 0.19521388 | 0.31969809 |
| 0.834242872 | 0.27209607 | 0.41363483 |
| 0.333491772 | 0.00203886 | 0.00934018 |
| 0.936955821 | 0.98689852 | 0.98951314 |
| 0.943542905 | 0.97368232 | 0.97946085 |
| 0.355922358 | 0.1437295 | 0.26691984 |
| 0.751154099 | 0.36788443 | 0.50903192 |
| 0.968522555 | 0.96508346 | 0.97583324 |
| 0.096210144 | 0.01402734 | 0.04852316 |
| 0.346870502 | 0.13510389 | 0.25875831 |
| 0.156437419 | 0.00424172 | 0.01732463 |
| 0.161042523 | 0.01907837 | 0.06159588 |
| 0.638043675 | 0.36571928 | 0.50810999 |
| 0.061194196 | 0.00053177 | 0.00321912 |
| 0.334323971 | 0.12388396 | 0.24416664 |
| 0.832625319 | 0.74240599 | 0.82956991 |
| 0.125287393 | 0.03172829 | 0.09115162 |
| 0.077093738 | 0.01356902 | 0.04742163 |
| 0.0054303 | 0.00018717 | 0.00124414 |
| 0.322253096 | 0.1616833 | 0.28399433 |
| 0.58886316 | 0.86879072 | 0.9261637 |
| 0.957352511 | 0.33069539 | 0.47720828 |
| 0.988098613 | 0.94794026 | 0.96385971 |
| 0.007630961 | 8.53E-05 | 0.0006022 |
| 0.625158469 | 0.50514394 | 0.63896939 |
| 0.204958079 | 0.0613204 | 0.14238094 |
| 0.901893574 | 0.62394313 | 0.74389166 |
| 0.936955821 | 0.71074181 | 0.81952883 |

| | | |
|---|---|---|
| 0.611226327 | 0.43834211 | 0.57904752 |
| 0.446990037 | 0.09557223 | 0.2029513 |
| 0.041368066 | 0.00548463 | 0.02161964 |
| 0.071734441 | 0.00755102 | 0.02844216 |
| 0.000408831 | 1.91E-05 | 0.00015793 |
| 8.10E-07 | 7.53E-09 | 1.50E-07 |
| 8.10E-07 | 8.07E-09 | 1.52E-07 |
| 0.021028187 | 0.0104812 | 0.03779921 |
| 0.392638364 | 0.14986777 | 0.27462255 |
| 0.355922358 | 0.03847442 | 0.10518409 |
| 0.029884279 | 0.00063542 | 0.00365097 |
| 0.029884279 | 0.00138901 | 0.00682427 |
| 0.020404834 | 0.00061064 | 0.00356911 |
| 0.424222026 | 0.13011111 | 0.25349234 |
| 0.896485864 | 0.94964349 | 0.96385971 |
| 0.371830337 | 0.11921531 | 0.23772935 |
| 0.902176059 | 0.76635521 | 0.84348837 |
| 0.579658492 | 0.56815282 | 0.69783987 |
| 0.446990037 | 0.01700472 | 0.05651568 |
| 0.081897847 | 0.00078091 | 0.0042698 |
| 0.278630716 | 0.0369683 | 0.10360712 |
| 0.454765835 | 0.07035944 | 0.15901234 |
| 0.234150954 | 0.07498062 | 0.16833398 |
| 0.823616657 | 0.26762484 | 0.40867036 |
| 0.9714085 | 0.87455962 | 0.92648659 |
| 0.956729348 | 0.48616811 | 0.62428405 |
| 0.798746833 | 0.5200553 | 0.65538568 |
| 0.986608478 | 0.88980655 | 0.93388365 |
| 0.914732727 | 0.76129597 | 0.8406493 |
| 0.392638364 | 0.05460944 | 0.13223285 |
| 0.864435832 | 0.94145317 | 0.9635595 |
| 0.424222026 | 0.0586033 | 0.13796195 |
| 5.64E-06 | 3.04E-09 | 6.86E-08 |
| 0.823270543 | 0.5026109 | 0.63896939 |
| 0.665592571 | 0.35677451 | 0.50185294 |
| 0.584223971 | 0.49360386 | 0.63144041 |
| 0.931460318 | 0.72778312 | 0.82956991 |
| 0.611366155 | 0.17108501 | 0.2899891 |
| 0.10147671 | 0.08949106 | 0.19323229 |
| 0.057122233 | 0.00174149 | 0.008315 |
| 0.694624561 | 0.25540654 | 0.40271078 |
| 0.133134378 | 0.00683523 | 0.02603532 |

| | | |
|---|---|---|
| 9.51E-07 | 1.39E-08 | 2.25E-07 |
| 0.852125517 | 0.67559432 | 0.78703255 |
| 0.59190873 | 0.00171306 | 0.01212316 |
| 0.8999887 | 0.05858966 | 0.19600906 |
| 0.92222507 | 0.35722473 | 0.614293 |
| 0.839598761 | 0.04174506 | 0.14492626 |
| 0.907579462 | 0.57638369 | 0.74657211 |
| 0.935694275 | 0.58998389 | 0.7538683 |
| 0.385161249 | 0.0208293 | 0.08844534 |
| 0.960469887 | 0.96836633 | 0.97640303 |
| 0.136092398 | 1.06E-07 | 3.92E-06 |
| 0.59190873 | 0.17291022 | 0.41318805 |
| 0.907579462 | 0.0015532 | 0.01143158 |
| 0.184167295 | 0.0003422 | 0.0033139 |
| 0.980407645 | 0.86214686 | 0.93675455 |
| 0.030166744 | 7.84E-06 | 0.00018029 |
| 0.960469887 | 0.7038659 | 0.84353414 |
| 0.000645788 | 2.71E-11 | 2.50E-09 |
| 0.800831289 | 0.4658235 | 0.69949845 |
| 0.047052622 | 0.00233099 | 0.01588526 |
| 0.385161249 | 0.09476425 | 0.27677176 |
| 0.907579462 | 0.01682288 | 0.0771106 |
| 0.800831289 | 0.03806304 | 0.140072 |
| 0.747964018 | 0.22396976 | 0.4630386 |
| 0.960469887 | 0.84615334 | 0.93675455 |
| 0.941061751 | 0.41016647 | 0.65993936 |
| 0.92222507 | 0.6973059 | 0.84353414 |
| 0.009572761 | 1.31E-05 | 0.00026737 |
| 0.980407645 | 0.75985363 | 0.87932747 |
| 0.92222507 | 0.3796315 | 0.64092599 |
| 0.912781938 | 0.39733127 | 0.65863923 |
| 0.095619954 | 2.59E-07 | 7.94E-06 |
| 0.930650719 | 0.89498693 | 0.94511367 |
| 0.907579462 | 0.27773173 | 0.53231915 |
| 0.839598761 | 0.22791862 | 0.46596696 |
| 0.907579462 | 0.77029422 | 0.88033625 |
| 0.907579462 | 0.02114997 | 0.08844534 |
| 0.960469887 | 0.34243136 | 0.59938058 |
| 0.402075885 | 0.10261616 | 0.29048266 |
| 0.467009332 | 0.22375941 | 0.4630386 |
| 0.907579462 | 0.57539278 | 0.74657211 |
| 0.785407209 | 0.06256176 | 0.2046343 |

| | | |
|---|---|---|
| 0.912781938 | 0.66542078 | 0.81084386 |
| 0.907579462 | 0.94163715 | 0.96256242 |
| 0.059533143 | 7.49E-09 | 3.44E-07 |
| 0.47105316 | 0.1084824 | 0.29556515 |
| 0.296484211 | 0.03613934 | 0.13570692 |
| 0.907579462 | 0.32443888 | 0.59696753 |
| 0.969953443 | 0.34529533 | 0.59938058 |
| 0.960469887 | 0.7328305 | 0.86343711 |
| 0.987738465 | 0.97109649 | 0.97640303 |
| 0.237075899 | 0.03962518 | 0.1421922 |
| 0.622925755 | 0.01958919 | 0.08581929 |
| 0.968047756 | 0.71058583 | 0.84353414 |
| 0.841326117 | 0.33614605 | 0.59938058 |
| 0.907579462 | 0.56680635 | 0.74657211 |
| 0.960469887 | 0.61107956 | 0.76790371 |
| 0.960469887 | 0.93447129 | 0.96057384 |
| 0.960469887 | 0.46873586 | 0.69949845 |
| 0.59190873 | 0.18159231 | 0.41766232 |
| 0.785407209 | 0.02403005 | 0.09825621 |
| 0.974040823 | 0.30561931 | 0.57381584 |
| 0.136092398 | 2.70E-05 | 0.00045146 |
| 0.960469887 | 0.43992282 | 0.68347929 |
| 0.184167295 | 0.00106405 | 0.0081577 |
| 0.005626984 | 1.94E-05 | 0.00035767 |
| 0.004108543 | 1.62E-10 | 9.93E-09 |
| 0.907579462 | 0.40335929 | 0.65993936 |
| 0.989115266 | 0.92599745 | 0.95721085 |
| 0.907579462 | 0.2022328 | 0.44298613 |
| 0.960469887 | 0.8652524 | 0.93675455 |
| 0.907579462 | 0.61906593 | 0.76964954 |
| 0.92222507 | 0.49269109 | 0.69949845 |
| 0.960469887 | 0.61348829 | 0.76790371 |
| 0.907579462 | 0.4793742 | 0.69949845 |
| 0.019657647 | 0.00012334 | 0.00141842 |
| 0.907579462 | 0.47519987 | 0.69949845 |
| 0.785407209 | 0.27241135 | 0.52876821 |
| 0.960469887 | 0.7588201 | 0.87932747 |
| 0.92222507 | 0.29026974 | 0.55061477 |
| 1.88E-05 | 1.17E-15 | 2.16E-13 |
| 0.960469887 | 0.57599367 | 0.74657211 |
| 0.385161249 | 0.02704454 | 0.10817816 |
| 0.644340143 | 0.00446242 | 0.02831326 |

| | | |
|---|---|---|
| 0.184167295 | 9.59E-05 | 0.00117599 |
| 0.960469887 | 0.80437004 | 0.90800054 |
| 0.467009332 | 0.08661913 | 0.25706321 |
| 0.907579462 | 0.42232261 | 0.66989103 |
| 0.907579462 | 0.4420328 | 0.68347929 |
| 0.941061751 | 0.4124621 | 0.65993936 |
| 0.92222507 | 0.53063335 | 0.72220431 |
| 0.960469887 | 0.65072493 | 0.79822258 |
| 0.785407209 | 0.15368855 | 0.37704923 |
| 0.385161249 | 0.01411089 | 0.06913191 |
| 0.980407645 | 0.99596359 | 0.99596359 |
| 0.930650719 | 0.76758807 | 0.88033625 |
| 0.907579462 | 0.08452837 | 0.25706321 |
| 0.907579462 | 0.50506125 | 0.70402477 |
| 0.907579462 | 0.44865247 | 0.68793379 |
| 0.907579462 | 0.8942276 | 0.94511367 |
| 0.980407645 | 0.4959362 | 0.69949845 |
| 0.467009332 | 0.16704969 | 0.4044361 |
| 0.747964018 | 0.17680913 | 0.41522541 |
| 0.907579462 | 0.7367371 | 0.86343711 |
| 0.92222507 | 0.31095315 | 0.57793313 |
| 0.184167295 | 5.38E-05 | 0.00077961 |
| 0.467009332 | 0.27300533 | 0.52876821 |
| 0.59190873 | 0.00057353 | 0.00502519 |
| 0.907579462 | 0.70837311 | 0.84353414 |
| 0.013946751 | 0.00039555 | 0.00363905 |
| 0.935694275 | 0.8673857 | 0.93675455 |
| 0.610400695 | 0.12741768 | 0.32562296 |
| 0.839598761 | 0.01530676 | 0.07221652 |
| 0.92222507 | 0.63329986 | 0.78206157 |
| 0.907579462 | 0.04481941 | 0.15271799 |
| 0.92222507 | 0.91354752 | 0.95507241 |
| 0.890108603 | 0.21071465 | 0.45083135 |
| 0.59190873 | 0.10159879 | 0.29048266 |
| 0.800831289 | 0.17827613 | 0.41522541 |
| 0.968047756 | 0.87566186 | 0.93675455 |
| 0.960469887 | 0.86079978 | 0.93675455 |
| 0.92222507 | 0.60441294 | 0.76697918 |
| 0.907579462 | 0.26699614 | 0.52825042 |
| 0.930650719 | 0.12287936 | 0.32299718 |
| 0.92222507 | 0.56380712 | 0.74657211 |
| 0.8999887 | 0.23327772 | 0.47168241 |

| | | |
|---|---|---|
| 0.59190873 | 0.37967898 | 0.64092599 |
| 0.907579462 | 0.18972109 | 0.43097136 |
| 0.980407645 | 0.32845233 | 0.5983686 |
| 0.602434537 | 0.19482988 | 0.43717924 |
| 0.136092398 | 0.00506055 | 0.03103802 |
| 0.907579462 | 0.13527234 | 0.34096041 |
| 0.907579462 | 0.42980609 | 0.67593437 |
| 0.402075885 | 0.00098537 | 0.00788295 |
| 0.961422306 | 0.58021637 | 0.74657211 |
| 0.92222507 | 0.20644849 | 0.44690025 |
| 0.59190873 | 0.2159235 | 0.45666578 |
| 0.59190873 | 0.19867917 | 0.44044539 |
| 0.92222507 | 0.89888528 | 0.94511367 |
| 0.907579462 | 0.49021935 | 0.69949845 |
| 0.467009332 | 0.14240352 | 0.35408443 |
| 0.839598761 | 0.4085576 | 0.65993936 |
| 0.907579462 | 0.46553381 | 0.69949845 |
| 0.995902533 | 0.34329602 | 0.59938058 |
| 0.680421205 | 0.01427724 | 0.06913191 |
| 0.907579462 | 0.51886609 | 0.71782977 |
| 0.907579462 | 0.1178332 | 0.31422186 |
| 0.680421205 | 0.57232315 | 0.74657211 |
| 0.907579462 | 0.00019621 | 0.00200574 |
| 0.092922987 | 5.51E-05 | 0.00077961 |
| 0.907579462 | 0.06634152 | 0.20689558 |
| 0.447467715 | 0.01344117 | 0.06913191 |
| 0.385161249 | 0.01412032 | 0.06913191 |
| 0.785407209 | 0.08549859 | 0.25706321 |
| 0.842358194 | 0.7872572 | 0.89416867 |
| 0.930650719 | 0.8721614 | 0.93675455 |
| 0.842358194 | 0.33927952 | 0.59938058 |
| 0.907579462 | 0.01718225 | 0.0771106 |
| 0.907579462 | 0.23902051 | 0.47804102 |
| 0.59190873 | 0.03423104 | 0.13121897 |
| 0.005626984 | 0.00068098 | 0.00569546 |
| 0.839598761 | 0.38430829 | 0.64284295 |
| 0.995902533 | 0.94792304 | 0.96363448 |
| 0.385161249 | 0.01167407 | 0.0631773 |
| 0.174906232 | 0.00429619 | 0.02823209 |
| 0.59190873 | 0.06585557 | 0.20689558 |
| 0.098273055 | 0.0069283 | 0.04112283 |
| 0.839598761 | 0.48114839 | 0.69949845 |

| | | |
|---|---|---|
| 0.59190873 | 0.1053461 | 0.29369217 |
| 0.960469887 | 0.53380319 | 0.72220431 |
| 0.530258727 | 0.02855527 | 0.11179084 |
| 0.136092398 | 0.00014832 | 0.00160534 |
| 0.453760892 | 7.54E-05 | 0.00099139 |
| 0.92222507 | 0.53164562 | 0.72220431 |
| 0.960469887 | 0.85879322 | 0.93675455 |
| 0.224850369 | 0.00740972 | 0.04260591 |
| 0.907579462 | 0.49267386 | 0.69949845 |
| 0.907579462 | 0.00900895 | 0.05023172 |
| 0.907579462 | 0.49801249 | 0.69949845 |
| 0.169943247 | 1.75E-06 | 4.59E-05 |
| 0.83003371 | 0.92398406 | 0.95721085 |
| 0.680421205 | 0.12663524 | 0.32562296 |
| 0.907579462 | 0.83914532 | 0.93675455 |
| 0.385161249 | 0.1092306 | 0.29556515 |
| 0.614640124 | 0.06339215 | 0.2046343 |
| 0.455271682 | 0.04018475 | 0.1421922 |
| 1.03E-13 | 2.62E-14 | 2.62E-14 |
