## Supplemental table 3 for "Early immune responses have long-term associations with clinical, virologic, and immunologic outcomes in patients with COVID-19"

| variable | severity_t | severity_p | viral_t |
|---|---|---|---|
| GO_activated_T_cell_proliferation | 2.36661607 | 0.01905727 | -1.6092771 |
| GO_activation_of_immune_response | 1.12769181 | 0.26101167 | -3.3090401 |
| GO_activation_of_innate_immune_response | 1.56608706 | 0.11915557 | -0.6389275 |
| GO_adaptive_immune_response | 0.45689283 | 0.64832125 | -2.7581777 |
| GO_alpha-beta_T_cell_activation | 3.0082914 | 0.0030194 | -0.0547637 |
| GO_alpha-beta_T_cell_differentiation | 2.68991449 | 0.00784715 | -0.1547694 |
| GO_alpha-beta_T_cell_lineage_commitment | 0.12167559 | 0.90329716 | -1.1596208 |
| GO_alpha-beta_T_cell_proliferation | 1.96875944 | 0.05057812 | 0.80916762 |
| GO_antibacterial_humoral_response | -0.2576365 | 0.79699365 | -1.1432958 |
| GO_antibacterial_peptide_production | 0.00460161 | 0.99633376 | -1.6282676 |
| GO_antifungal_humoral_response | 0.7601319 | 0.44821041 | -0.7016251 |
| GO_antifungal_innate_immune_response | -0.4039941 | 0.68671599 | -1.8682742 |
| GO_antigen_processing_and_presentation | 1.44105756 | 0.15137595 | 0.57115286 |
| GO_antimicrobial_humoral_response | 0.35680101 | 0.7216756 | -1.4900212 |
| GO_antimicrobial_peptide_production | 0.00460161 | 0.99633376 | -1.6282676 |
| GO_B_cell_activation | -0.3903496 | 0.69675824 | -3.7835965 |
| GO_B_cell_chemotaxis | 1.51614098 | 0.13130879 | -2.7033705 |
| GO_B_cell_differentiation | -0.3440699 | 0.73121156 | -2.7047512 |
| GO_B_cell_homeostasis | 0.72978294 | 0.46650986 | 0.47179165 |
| GO_B_cell_lineage_commitment | -1.0778388 | 0.2826063 | -0.0744752 |
| GO_B_cell_mediated_immunity | -0.0588741 | 0.95312033 | -3.6282538 |
| GO_B_cell_proliferation | 0.4467604 | 0.65560683 | -3.0160223 |
| GO_B_cell_receptor_signaling_pathway | -1.2247788 | 0.22232366 | -3.3732604 |
| GO_B-1_B_cell_differentiation | -1.7248681 | 0.08633717 | -0.1451576 |
| GO_bone_marrow_development | -0.8087913 | 0.41974607 | -1.1899101 |
| GO_cellular_extravasation | 2.04762505 | 0.04210856 | -2.6222967 |
| GO_complement_activation | 0.25931381 | 0.79570143 | -4.0585761 |
| GO_defense_response_to_virus | 3.25254822 | 0.00137522 | 1.43379855 |
| GO_definitive_hemopoiesis | 0.03978148 | 0.96831321 | 0.74752236 |
| GO_dendritic_cell_chemotaxis | 0.72430404 | 0.46985748 | 0.58634205 |
| GO_dendritic_cell_cytokine_production | 2.77832179 | 0.00606764 | 0.18446127 |
| GO_dendritic_cell_differentiation | 1.87868395 | 0.0619698 | 0.15861508 |
| GO_dendritic_cell_migration | 0.83715238 | 0.40366188 | 0.2181635 |
| GO_diapedesis | 0.27015656 | 0.78736181 | -0.081605 |
| GO_embryonic_hemopoiesis | 1.04764518 | 0.29626356 | -0.5251838 |
| GO_eosinophil_activation | 0.77571455 | 0.43897666 | 0.70977368 |
| GO_eosinophil_chemotaxis | 0.70133449 | 0.48403663 | -1.480817 |
| GO_eosinophil_degranulation | 0.43193666 | 0.66632531 | 0.27622326 |
| GO_eosinophil_differentiation | 1.00370313 | 0.31692354 | 0.51459597 |
| GO_eosinophil_mediated_immunity | 0.43193666 | 0.66632531 | 0.27622326 |
| GO_eosinophil_migration | 0.68319232 | 0.49539925 | -1.4666746 |

| | | | |
|---|---|---|---|
| GO_erythrocyte_development | 0.49164648 | 0.62359249 | 0.79369524 |
| GO_erythrocyte_differentiation | 1.48551255 | 0.13922808 | 0.71658385 |
| GO_erythrocyte_homeostasis | 1.4464801 | 0.14985195 | 0.79554938 |
| GO_erythrocyte_maturation | 0.13698108 | 0.89120503 | 0.46166985 |
| GO_establishment_of_lymphocyte_polarity | 0.25129784 | 0.80188209 | -0.7827538 |
| GO_establishment_of_T_cell_polarity | 0.11868138 | 0.90566548 | -0.6642568 |
| GO_Fc_receptor_signaling_pathway | 0.31427749 | 0.75368896 | -2.5071722 |
| GO_Fc-gamma_receptor_signaling_pathway | 0.04641345 | 0.96303425 | -3.0129284 |
| GO_gamma-delta_T_cell_activation | -0.8159854 | 0.41563057 | -0.4837646 |
| GO_gamma-delta_T_cell_differentiation | -1.2943088 | 0.19728383 | -1.5933931 |
| GO_germinal_center_formation | 1.89913612 | 0.05921086 | -0.906788 |
| GO_granulocyte_activation | 1.50684126 | 0.13367518 | -0.6916411 |
| GO_granulocyte_chemotaxis | 1.56369244 | 0.11971721 | -1.4246738 |
| GO_granulocyte_differentiation | 1.31903729 | 0.1888999 | 0.30680758 |
| GO_granulocyte_migration | 1.64268764 | 0.10226395 | -1.6229436 |
| GO_hemopoiesis | 1.87061103 | 0.06308796 | -0.881074 |
| GO_histamine_secretion_by_mast_cell | 0.23177497 | 0.81698673 | 0.46363609 |
| GO_humoral_immune_response | 0.32166381 | 0.74809571 | -3.7531979 |
| GO_hypersensitivity | 0.68261834 | 0.49576107 | 0.65516952 |
| GO_immature_B_cell_differentiation | 0.2943851 | 0.76881658 | -1.4433662 |
| GO_immature_T_cell_proliferation | 0.96915186 | 0.3338224 | -2.7090229 |
| GO_immune_effector_process | 2.49836355 | 0.01340992 | -1.197104 |
| GO_immune_response | 2.23009073 | 0.02702765 | -1.7384369 |
| GO_immune_response_to_tumor_cell | 0.9199281 | 0.35889126 | -0.6395855 |
| GO_immune_system_development | 1.83112112 | 0.06880268 | -1.0280355 |
| GO_immunoglobulin_biosynthetic_process | 1.62981468 | 0.10496008 | -3.1164232 |
| GO_immunoglobulin_production | -0.0147608 | 0.98824 | -2.9624897 |
| GO_immunoglobulin_secretion | 0.67696622 | 0.49933158 | -0.5497911 |
| GO_immunoglobulin_V(D)J_recombination | 0.74926686 | 0.45471397 | -0.1852737 |
| GO_immunological_memory_process | 0.2179773 | 0.82770369 | -1.4185266 |
| GO_immunological_synapse_formation | -0.7714289 | 0.44150516 | 0.98656898 |
| GO_induction_of_bacterial_agglutination | 0.10168321 | 0.91912589 | -1.7613937 |
| GO_innate_immune_response | 2.88131852 | 0.00446174 | -0.5702267 |
| GO_innate_immune_response_in_mucosa | 0.13010723 | 0.89663275 | -0.0231616 |
| GO_isotype_switching | 1.02180189 | 0.30830141 | -1.2457281 |
| GO_isotype_switching_to_IgA_isotypes | -2.061167 | 0.04078333 | -0.1020207 |
| GO_isotype_switching_to_IgE_isotypes | -0.3986575 | 0.69063721 | -1.6759318 |
| GO_isotype_switching_to_IgG_isotypes | -0.9124247 | 0.36281493 | 0.79994229 |
| GO_leukocyte_activation | 1.57275233 | 0.11760324 | -1.5235444 |
| GO_leukocyte_chemotaxis | 1.87953507 | 0.06185288 | -1.8393845 |
| GO_leukocyte_degranulation | 1.56182309 | 0.1201571 | -0.7597753 |
| GO_leukocyte_differentiation | 1.60315049 | 0.11072561 | -1.0861418 |

| | | | |
|---|---|---|---|
| GO_leukocyte_homeostasis | 1.39324026 | 0.16533468 | -0.0984503 |
| GO_leukocyte_mediated_cytotoxicity | 1.0110106 | 0.31342329 | -0.2151103 |
| GO_leukocyte_mediated_immunity | 1.4366169 | 0.15263283 | -2.1297651 |
| GO_leukocyte_migration | 1.29052177 | 0.19859166 | -3.7985144 |
| GO_leukocyte_tethering_or_rolling | 0.09967577 | 0.92071715 | -2.712137 |
| GO_lymph_node_development | 1.05507779 | 0.29286098 | 0.2197517 |
| GO_lymphocyte_activation | 0.80701925 | 0.4207635 | -1.9576194 |
| GO_lymphocyte_anergy | -4.4362023 | 1.62E-05 | 0.83350306 |
| GO_lymphocyte_chemotaxis | 2.18369521 | 0.03033081 | -2.195787 |
| GO_lymphocyte_costimulation | 0.91178108 | 0.36315274 | 0.28169862 |
| GO_lymphocyte_differentiation | 0.98570085 | 0.32565649 | -1.034396 |
| GO_lymphocyte_homeostasis | 0.95216748 | 0.34234047 | 0.3600984 |
| GO_lymphocyte_mediated_immunity | 0.40676545 | 0.68468305 | -3.4842218 |
| GO_lymphocyte_migration | 1.8942005 | 0.05986707 | -2.1135675 |
| GO_lymphocyte_proliferation | 2.20322887 | 0.0288997 | -0.839632 |
| GO_macrophage_activation | 2.32120996 | 0.02144156 | -2.6413858 |
| GO_macrophage_chemotaxis | 1.79520549 | 0.07436651 | -1.9852261 |
| GO_macrophage_cytokine_production | 0.31516997 | 0.75301245 | -0.7097357 |
| GO_macrophage_differentiation | 1.05909949 | 0.29103096 | -1.9591127 |
| GO_macrophage_migration | 1.71677804 | 0.08780949 | -2.4470358 |
| GO_marginal_zone_B_cell_differentiation | 1.56598518 | 0.11917942 | 0.421242 |
| GO_mast_cell_activation | 2.02126784 | 0.04479349 | -1.1140872 |
| GO_mast_cell_chemotaxis | 0.57760105 | 0.56428448 | -3.021089 |
| GO_mast_cell_cytokine_production | 1.99071066 | 0.04808724 | -1.5706199 |
| GO_mast_cell_degranulation | 1.61668926 | 0.10776739 | -0.9416639 |
| GO_mast_cell_differentiation | -0.224292 | 0.82279481 | -0.8557947 |
| GO_mast_cell_mediated_immunity | 1.69795398 | 0.0913144 | -0.8017935 |
| GO_mast_cell_migration | 0.57760105 | 0.56428448 | -3.021089 |
| GO_mature_B_cell_differentiation | -0.2770435 | 0.78207742 | -0.056881 |
| GO_MDA-5_signaling_pathway | 3.35064955 | 0.0009899 | 1.96478311 |
| GO_megakaryocyte_development | 1.58422832 | 0.11496807 | -0.5395623 |
| GO_megakaryocyte_differentiation | 1.54911613 | 0.12318121 | -0.468989 |
| GO_memory_T_cell_differentiation | 1.2961366 | 0.19665487 | -2.3269477 |
| GO_microglia_differentiation | 1.27777429 | 0.20304095 | -1.1640913 |
| GO_microglial_cell_activation | 2.62897229 | 0.00933539 | -2.5287581 |
| GO_microglial_cell_migration | 2.45019947 | 0.01527321 | -1.9759922 |
| GO_monocyte_activation | 1.97403734 | 0.04996947 | -3.4545429 |
| GO_monocyte_chemotaxis | 1.6189115 | 0.10728792 | -1.6885845 |
| GO_monocyte_differentiation | -0.6952546 | 0.48782859 | -1.8907175 |
| GO_monocyte_extravasation | -0.0421473 | 0.96642991 | -0.4136278 |
| GO_mononuclear_cell_differentiation | -0.6952546 | 0.48782859 | -1.8907175 |
| GO_mononuclear_cell_migration | 1.83879348 | 0.06765998 | -1.6604927 |

| | | | |
|---|---|---|---|
| GO_mucosal_immune_response | 0.78014444 | 0.43637193 | -0.5489568 |
| GO_myeloid_cell_development | 1.2159801 | 0.2256491 | 0.01051144 |
| GO_myeloid_cell_differentiation | 1.85748446 | 0.06494198 | -0.4875687 |
| GO_myeloid_cell_homeostasis | 1.57074102 | 0.11806997 | 0.64992532 |
| GO_myeloid_dendritic_cell_activation | 1.74748305 | 0.08232792 | -0.7730578 |
| GO_myeloid_dendritic_cell_chemotaxis | -1.5555573 | 0.12164088 | 1.45813178 |
| GO_myeloid_leukocyte_activation | 1.7161545 | 0.08792381 | -0.9996547 |
| GO_myeloid_leukocyte_differentiation | 1.54445552 | 0.12430529 | -1.1824434 |
| GO_myeloid_leukocyte_mediated_immunity | 1.59561493 | 0.11239994 | -0.7537422 |
| GO_myeloid_leukocyte_migration | 1.77941329 | 0.07692774 | -2.0287018 |
| GO_natural_killer_cell_activation | 1.6020779 | 0.11096271 | -0.6452212 |
| GO_natural_killer_cell_chemotaxis | 0.75636643 | 0.45045828 | -0.0514081 |
| GO_natural_killer_cell_degranulation | 1.50405237 | 0.13439129 | -0.0929497 |
| GO_natural_killer_cell_differentiation | 0.3151923 | 0.75299552 | -1.0025414 |
| GO_natural_killer_cell_proliferation | 0.74640706 | 0.45643466 | 0.49449493 |
| GO_negative_T_cell_selection | -0.9372431 | 0.34994009 | 1.92647738 |
| GO_negative_thymic_T_cell_selection | -0.8541339 | 0.39421196 | 1.85630613 |
| GO_neutrophil_activation | 1.50135966 | 0.13508553 | -0.6929244 |
| GO_neutrophil_apoptotic_process | 1.98790875 | 0.04839928 | -0.9470371 |
| GO_neutrophil_chemotaxis | 1.48684616 | 0.1388757 | -1.1782841 |
| GO_neutrophil_clearance | 1.5227232 | 0.12965377 | 0.72403706 |
| GO_neutrophil_degranulation | 1.51278629 | 0.13215861 | -0.7490679 |
| GO_neutrophil_differentiation | -0.2366482 | 0.81320968 | -0.1645834 |
| GO_neutrophil_extravasation | 2.83672033 | 0.00510228 | -1.886504 |
| GO_neutrophil_homeostasis | 2.0586146 | 0.04103035 | -0.5939479 |
| GO_neutrophil_mediated_cytotoxicity | 0.55799637 | 0.57756824 | 0.12620556 |
| GO_neutrophil_mediated_immunity | 1.52167954 | 0.12991509 | -0.7587663 |
| GO_neutrophil_migration | 1.6518038 | 0.10038848 | -1.4603873 |
| GO_NK_T_cell_activation | 3.38368541 | 0.00088469 | -0.9832066 |
| GO_NK_T_cell_differentiation | 0.55302156 | 0.58096256 | -0.0300625 |
| GO_NK_T_cell_proliferation | 3.01168166 | 0.00298756 | -0.217326 |
| GO_opsonization | 1.1013089 | 0.27229215 | -1.3813794 |
| GO_osteoclast_development | 1.54777781 | 0.12350318 | -1.2003995 |
| GO_osteoclast_differentiation | 1.84986562 | 0.06603872 | -1.2506749 |
| GO_osteoclast_fusion | 0.70333814 | 0.48279052 | -0.4411764 |
| GO_peripheral_tolerance_induction | 3.31484654 | 0.00111704 | -1.1035249 |
| GO_Peyer's_patch_development | -1.4614834 | 0.14569681 | -1.8592917 |
| GO_platelet_formation | 1.44736086 | 0.14960554 | -1.1462652 |
| GO_positive_T_cell_selection | -0.7674689 | 0.44384894 | 0.9844477 |
| GO_positive_thymic_T_cell_selection | -1.6997508 | 0.09097503 | 2.40631371 |
| GO_pre-B_cell_differentiation | 0.18389056 | 0.85431467 | -1.7672882 |
| GO_primitive_hemopoiesis | 0.97705036 | 0.3299085 | 0.60157242 |

| | | | |
|---|---|---|---|
| GO_pro-B_cell_differentiation | 0.88603208 | 0.37683019 | -1.2122133 |
| GO_regulatory_T_cell_differentiation | 2.30483289 | 0.02236371 | -0.1790851 |
| GO_response_to_interferon-gamma | 3.31326522 | 0.00112299 | 0.4128698 |
| GO_response_to_type_I_interferon | 2.93442685 | 0.00379526 | 1.52978772 |
| GO_RIG-I_signaling_pathway | 2.85175069 | 0.00487759 | 2.19815214 |
| GO_spleen_development | 0.80213551 | 0.42357506 | -2.1756068 |
| GO_T_cell_activation | 1.51408713 | 0.13182857 | -0.0116945 |
| GO_T_cell_anergy | -4.4362023 | 1.62E-05 | 0.83350306 |
| GO_T_cell_chemotaxis | 1.94723713 | 0.05312534 | -1.2623337 |
| GO_T_cell_costimulation | 0.84838311 | 0.39739696 | 0.31091366 |
| GO_T_cell_cytokine_production | 3.22477287 | 0.00150736 | -0.795189 |
| GO_T_cell_differentiation | 1.41837238 | 0.15788096 | -0.1083581 |
| GO_T_cell_differentiation_in_thymus | -0.2410605 | 0.80979366 | 0.29819037 |
| GO_T_cell_extravasation | 2.74566978 | 0.00667706 | -0.1120446 |
| GO_T_cell_homeostasis | 1.20296649 | 0.23063301 | 0.1993114 |
| GO_T_cell_lineage_commitment | 0.27193148 | 0.78599896 | -1.9253181 |
| GO_T_cell_mediated_cytotoxicity | 0.35149347 | 0.7256459 | 0.44626222 |
| GO_T_cell_mediated_immunity | 1.70789436 | 0.08944969 | -0.0610962 |
| GO_T_cell_migration | 1.69259534 | 0.09233263 | -0.864185 |
| GO_T_cell_proliferation | 2.51414089 | 0.0128452 | 0.17769135 |
| GO_T_cell_receptor_signaling_pathway | 1.19735888 | 0.23280475 | 1.63882976 |
| GO_T_cell_receptor_V(D)J_recombination | -1.2598569 | 0.2094179 | 2.10107888 |
| GO_T_cell_selection | -1.3418252 | 0.1814112 | 1.52134523 |
| GO_T_cell_tolerance_induction | -0.2830682 | 0.777463 | 2.15715849 |
| GO_T-helper_1_cell_cytokine_production | 3.07530628 | 0.00244461 | -1.3114019 |
| GO_T-helper_1_cell_differentiation | 3.42487236 | 0.00076816 | -0.8013282 |
| GO_T-helper_1_type_immune_response | 3.72691495 | 0.00026229 | -1.5383116 |
| GO_T-helper_17_cell_differentiation | 2.43192051 | 0.01603844 | -1.8323366 |
| GO_T-helper_17_cell_lineage_commitment | 1.32556424 | 0.18673185 | -2.1725368 |
| GO_T-helper_17_type_immune_response | 2.28432331 | 0.02356748 | -1.7397377 |
| GO_T-helper_2_cell_cytokine_production | 3.20766936 | 0.00159451 | 0.50637404 |
| GO_T-helper_2_cell_differentiation | 1.5826791 | 0.11532105 | -0.8020908 |
| GO_T-helper_cell_differentiation | 3.26445755 | 0.00132193 | -1.8663017 |
| GO_T-helper_cell_lineage_commitment | 0.46731426 | 0.64086312 | -1.2563536 |
| GO_thymic_T_cell_selection | -1.4400629 | 0.15165679 | 1.96595373 |
| GO_thymocyte_migration | 2.63290599 | 0.00923215 | -2.4982453 |
| GO_thymus_development | 0.83260574 | 0.40621503 | -3.0398539 |
| GO_tolerance_induction | 1.88561086 | 0.0610236 | 1.44382305 |
| GO_tolerance_induction_to_self_antigen | 2.18457922 | 0.03026474 | -0.3953979 |
| GO_type_2_immune_response | 2.29360668 | 0.02301576 | -0.6018234 |
| GO_type_I_interferon_signaling_pathway | 2.90271353 | 0.00418133 | 1.49418969 |
| GO_V(D)J_recombination | -0.5932821 | 0.55376723 | -0.6032119 |

| | | | |
|---|---|---|---|
| olink_IL8_P10145_OID00471 | 1.76830737 | 0.07888016 | -1.5700003 |
| olink_VEGFA_P15692_OID00472 | 2.25278997 | 0.02560657 | -1.8987513 |
| olink_CD8A_P01732_OID05124 | 1.25378602 | 0.21171502 | 3.13669428 |
| olink_MCP-3_P80098_OID00474 | 3.36708785 | 0.00094773 | -3.1360844 |
| olink_GDNF_P39905_OID00475 | 0.70601682 | 0.48118555 | 0.47341762 |
| olink_CDCP1_Q9H5V8_OID00476 | 0.24935899 | 0.80339706 | -1.4359196 |
| olink_CD244_Q9BZW8_OID00477 | -2.1743347 | 0.03112189 | 1.37557787 |
| olink_IL7_P13232_OID00478 | 0.43720663 | 0.66254019 | -0.1939016 |
| olink_OPG_O00300_OID00479 | 0.8916311 | 0.373905 | -0.5093036 |
| olink_LAP TGF-beta-1_P01137_OID00480 | 0.45646078 | 0.64866631 | -0.5923005 |
| olink_uPA_P00749_OID00481 | -1.0218831 | 0.30835048 | 1.19512918 |
| olink_IL6_P05231_OID00482 | 2.55434325 | 0.01155477 | -2.9703566 |
| olink_IL-17C_Q9P0M4_OID00483 | -1.2935507 | 0.19765073 | -0.7774771 |
| olink_MCP-1_P13500_OID00484 | 4.52226821 | 1.17E-05 | -2.5219944 |
| olink_IL-17A_Q16552_OID00485 | -0.0372335 | 0.97034439 | 0.59238133 |
| olink_CXCL11_O14625_OID00486 | 3.78630797 | 0.00021462 | -0.9980461 |
| olink_AXIN1_O15169_OID00487 | 2.46497877 | 0.01473906 | 0.69417831 |
| olink_TRAIL_P50591_OID00488 | 1.34488923 | 0.18052931 | 3.91323207 |
| olink_IL-20RA_Q9UHF4_OID00489 | 1.64609143 | 0.10167173 | 0.16678905 |
| olink_CXCL9_Q07325_OID00490 | 2.18226031 | 0.0305215 | -2.1313945 |
| olink_CST5_P28325_OID00491 | -0.2832077 | 0.77737698 | -0.0258166 |
| olink_IL-2RB_P14784_OID00492 | 0.2361812 | 0.81358858 | 0.01721102 |
| olink_IL-1 alpha_P01583_OID00493 | 0.09095396 | 0.92764085 | -1.0983319 |
| olink_OSM_P13725_OID00494 | 2.79052846 | 0.00589032 | -1.3573749 |
| olink_IL2_P60568_OID00495 | 0.04980904 | 0.96033554 | 0.84283472 |
| olink_CXCL1_P09341_OID00496 | 2.19680136 | 0.02944605 | -0.988065 |
| olink_TSLP_Q969D9_OID00497 | 0.03653349 | 0.97090168 | -1.1966192 |
| olink_CCL4_P13236_OID00498 | 2.24469475 | 0.02613317 | -0.172288 |
| olink_CD6_P30203_OID00499 | -1.8091907 | 0.072264 | 2.01923515 |
| olink_SCF_P21583_OID00500 | 1.60828271 | 0.10970848 | 3.70363409 |
| olink_IL18_Q14116_OID00501 | -0.0408597 | 0.96745771 | -0.3840917 |
| olink_SLAMF1_Q13291_OID00502 | -0.5997145 | 0.54952929 | 0.04952836 |
| olink_TGF-alpha_P01135_OID00503 | 1.91173757 | 0.05766428 | -1.1122619 |
| olink_MCP-4_Q99616_OID00504 | 0.40859531 | 0.68337296 | 0.29201951 |
| olink_CCL11_P51671_OID00505 | 0.98260185 | 0.32725965 | -1.7639061 |
| olink_TNFSF14_O43557_OID00506 | 1.74011989 | 0.08372532 | -2.1529658 |
| olink_FGF-23_Q9GZV9_OID00507 | 0.4273008 | 0.66972418 | 0.80993849 |
| olink_IL-10RA_Q13651_OID00508 | 0.21510872 | 0.82995146 | 0.32578699 |
| olink_FGF-5_P12034_OID00509 | -0.6789775 | 0.49811503 | 0.65839388 |
| olink_MMP-1_P03956_OID00510 | 1.58734707 | 0.11437201 | -1.0640234 |
| olink_LIF-R_P42702_OID00511 | 0.44665503 | 0.65571701 | -0.5251746 |
| olink_FGF-21_Q9NSA1_OID00512 | -1.1338431 | 0.25852518 | -1.4758778 |

| | | | |
|---|---|---|---|
| olink_CCL19_Q99731_OID00513 | 0.59966767 | 0.5495604 | 2.25550295 |
| olink_IL-15RA_Q13261_OID00514 | 0.07679774 | 0.93887863 | 0.93371179 |
| olink_IL-10RB_Q08334_OID00515 | -1.1217369 | 0.26362407 | 2.12344289 |
| olink_IL-22 RA1_Q8N6P7_OID00516 | -0.788186 | 0.43173268 | -0.4272365 |
| olink_IL-18R1_Q13478_OID00517 | 2.75526095 | 0.00653201 | -1.286214 |
| olink_PD-L1_Q9NZQ7_OID00518 | 3.00549973 | 0.00307098 | 0.4958569 |
| olink_Beta-NGF_P01138_OID00519 | -0.4105494 | 0.68194214 | 0.38954721 |
| olink_CXCL5_P42830_OID00520 | 0.28007091 | 0.77977822 | -0.2685073 |
| olink_TRANCE_O14788_OID00521 | -2.6899976 | 0.00788916 | 0.58460533 |
| olink_HGF_P14210_OID00522 | 1.73857525 | 0.08399771 | -1.7563442 |
| olink_IL-12B_P29460_OID00523 | -0.5526072 | 0.58128941 | 0.68137772 |
| olink_IL-24_Q13007_OID00524 | 1.40509296 | 0.1618966 | 0.87434512 |
| olink_IL13_P35225_OID00525 | -0.7422808 | 0.45898553 | -0.5234957 |
| olink_ARTN_Q5T4W7_OID00526 | -0.4796049 | 0.63215128 | 2.21548241 |
| olink_MMP-10_P09238_OID00527 | -1.0293711 | 0.30483036 | 2.00080852 |
| olink_IL10_P22301_OID00528 | 1.40262331 | 0.16263096 | -0.208346 |
| olink_TNF_P01375_OID05548 | 1.08516733 | 0.27944989 | -0.3327545 |
| olink_CCL23_P55773_OID00530 | 0.91586931 | 0.36108843 | -2.7391194 |
| olink_CD5_P06127_OID00531 | -1.7406937 | 0.08362433 | 0.54257641 |
| olink_CCL3_P10147_OID00532 | 2.59648626 | 0.01027905 | -1.0523723 |
| olink_Flt3L_P49771_OID00533 | -0.2353318 | 0.81424659 | 0.38945893 |
| olink_CXCL6_P80162_OID00534 | 1.20577576 | 0.22965134 | 0.94621636 |
| olink_CXCL10_P02778_OID00535 | 2.90444769 | 0.00418975 | -2.0179152 |
| olink_4E-BP1_Q13541_OID00536 | 2.99646305 | 0.00315851 | -1.4823785 |
| olink_IL-20_Q9NYY1_OID00537 | -0.3106805 | 0.75644048 | -0.9179571 |
| olink_SIRT2_Q8IXJ6_OID00538 | 0.42462274 | 0.67167166 | -1.1933123 |
| olink_CCL28_Q9NRJ3_OID00539 | -1.012258 | 0.31291494 | 0.04246998 |
| olink_DNER_Q8NFT8_OID01213 | -1.1649271 | 0.24575052 | 1.32838298 |
| olink_EN-RAGE_P80511_OID00541 | 1.37119832 | 0.17219797 | -1.063398 |
| olink_CD40_P25942_OID00542 | 0.76058569 | 0.44800338 | -1.3672454 |
| olink_IL33_O95760_OID00543 | -0.8541803 | 0.39425869 | -0.6142523 |
| olink_IFN-gamma_P01579_OID05547 | 2.23494529 | 0.02677991 | -1.7801991 |
| olink_FGF-19_O95750_OID00545 | -0.5177645 | 0.6053246 | -0.0719502 |
| olink_IL4_P05112_OID00546 | -0.3443306 | 0.73104149 | 1.18788552 |
| olink_LIF_P15018_OID00547 | 0.20467315 | 0.83808285 | 0.35310463 |
| olink_NRTN_Q99748_OID00548 | -0.2192883 | 0.82669982 | -0.8302809 |
| olink_MCP-2_P80075_OID00549 | 3.64622641 | 0.0003576 | 0.61681975 |
| olink_CASP-8_Q14790_OID00550 | 2.03423331 | 0.04354905 | -2.0607709 |
| olink_CCL25_O15444_OID00551 | 1.62726783 | 0.10561176 | -3.9471303 |
| olink_CX3CL1_P78423_OID00552 | 2.50515231 | 0.01322181 | 0.97009303 |
| olink_TNFRSF9_Q07011_OID00553 | -1.6677663 | 0.09728214 | 2.26557525 |
| olink_NT-3_P20783_OID00554 | 0.42483022 | 0.6715207 | -0.9623831 |

| | | | |
|---|---|---|---|
| olink_TWEAK_O43508_OID00555 | -2.6433272 | 0.00901078 | 2.4651075 |
| olink_CCL20_P78556_OID00556 | 1.27648772 | 0.20359883 | -3.8449724 |
| olink_ST1A1_P50225_OID00557 | 1.75233232 | 0.08159706 | 0.36465355 |
| olink_STAMBP_O95630_OID00558 | 1.77496427 | 0.07777016 | -1.4423717 |
| olink_IL5_P05113_OID00559 | 0.09039011 | 0.92808819 | 2.41777608 |
| olink_ADA_P00813_OID00560 | 1.24062709 | 0.21652626 | -1.8018269 |
| olink_TNFB_P01374_OID00561 | -1.4090286 | 0.16073156 | 1.42621416 |
| olink_CSF-1_P09603_OID00562 | 3.35132795 | 0.00099962 | -0.4876947 |
| olink_PPP1R9B_Q96SB3_OID00936 | 2.5533942 | 0.01158507 | 0.2865664 |
| olink_GLB1_P16278_OID00937 | 0.93763188 | 0.34982084 | -1.108708 |
| olink_PSIP1_O75475_OID00938 | 0.28045726 | 0.77948235 | -3.4566674 |
| olink_ZBTB16_Q05516_OID00939 | 2.19943355 | 0.02925493 | -1.356226 |
| olink_IRAK4_Q9NWZ3_OID00940 | -0.9318758 | 0.3527789 | 1.74223935 |
| olink_TPSAB1_Q15661_OID00941 | -1.7954115 | 0.07444071 | -1.7996123 |
| olink_HCLS1_P14317_OID00942 | 1.85874776 | 0.06486593 | -2.8436141 |
| olink_CNTNAP2_Q9UHC6_OID00943 | -3.3522182 | 0.00099662 | 0.64591755 |
| olink_CLEC4G_Q6UXB4_OID00944 | 1.12486736 | 0.26229893 | 2.13102988 |
| olink_IRF9_Q00978_OID00945 | 0.57455359 | 0.56638535 | -0.9245105 |
| olink_EDAR_Q9UNE0_OID00946 | -0.3469577 | 0.72907065 | -0.1722122 |
| olink_IL6_P05231_OID00947 | 2.46941004 | 0.01456442 | -3.3692204 |
| olink_DGKZ_Q13574_OID00948 | 0.4254106 | 0.6710985 | 2.21440907 |
| olink_CLEC4C_Q8WTT0_OID00949 | -0.1568049 | 0.87559266 | 4.03393614 |
| olink_IRAK1_P51617_OID00950 | 2.21748379 | 0.02797313 | -1.4783495 |
| olink_CLEC4A_Q9UMR7_OID00951 | -0.8029902 | 0.42314971 | -0.3124326 |
| olink_PRDX1_Q06830_OID00952 | 2.58515746 | 0.01060893 | -2.6120192 |
| olink_PRDX3_P30048_OID00953 | 1.68928665 | 0.0930764 | -2.0957516 |
| olink_FGF2_P09038_OID00954 | 0.39139276 | 0.69601811 | 0.44963878 |
| olink_PRDX5_P30044_OID00955 | 3.82112853 | 0.00018862 | -2.1145012 |
| olink_DPP10_Q8N608_OID00956 | -2.6050876 | 0.01003478 | -0.1474695 |
| olink_TRIM5_Q9C035_OID00957 | 2.63813218 | 0.00914414 | -1.7260649 |
| olink_DCTN1_Q14203_OID00958 | 0.58180804 | 0.56149977 | -0.2372621 |
| olink_ITGA6_P23229_OID00959 | -0.7977215 | 0.42619268 | -0.66649 |
| olink_CDSN_Q15517_OID00960 | -0.4758588 | 0.63481212 | 0.19969264 |
| olink_GALNT3_Q14435_OID00961 | 0.54235089 | 0.58831752 | -0.3866115 |
| olink_FXYD5_Q96DB9_OID00962 | 1.00264967 | 0.31751604 | -0.1629257 |
| olink_TRAF2_Q12933_OID00963 | 1.09142331 | 0.27669731 | -0.1401321 |
| olink_TRIM21_P19474_OID00964 | 1.16692722 | 0.24494409 | -2.9848833 |
| olink_LILRB4_Q8NHJ6_OID00965 | 3.31720308 | 0.0011212 | -2.4914302 |
| olink_NTF4_P34130_OID00966 | -1.4880444 | 0.13867124 | -0.326013 |
| olink_KRT19_P08727_OID00967 | 0.9118035 | 0.3632187 | -3.5522935 |
| olink_ITM2A_O43736_OID00968 | -1.8029918 | 0.07323665 | 0.70852877 |
| olink_HNMT_P50135_OID00969 | 0.45396009 | 0.65046142 | -2.9274071 |

| | | | |
|---|---|---|---|
| olink_CCL11_P51671_OID00970 | 1.68492234 | 0.0939172 | -2.1253738 |
| olink_MILR1_Q7Z6M3_OID00971 | -1.5360722 | 0.12645951 | -2.1462657 |
| olink_EGLN1_Q9GZT9_OID00972 | 1.59657565 | 0.11229724 | -2.0828961 |
| olink_NFATC3_Q12968_OID00973 | -1.3173075 | 0.18958432 | -0.7452009 |
| olink_LY75_O60449_OID00974 | -0.1018954 | 0.91896491 | 0.75822804 |
| olink_EIF5A_P63241_OID00975 | -2.0366271 | 0.04330554 | 0.31479916 |
| olink_EIF4G1_Q04637_OID00976 | 1.95661829 | 0.05210103 | -2.340216 |
| olink_CD28_P10747_OID00977 | 0.06811931 | 0.94577415 | -1.8808311 |
| olink_PTH1R_Q03431_OID00978 | -1.6928183 | 0.09240049 | 0.4623959 |
| olink_BIRC2_Q13490_OID00979 | -0.5448284 | 0.58661617 | 0.73346697 |
| olink_HSD11B1_P28845_OID00980 | 0.96952022 | 0.33372211 | -0.2728242 |
| olink_NF2_P35240_OID00981 | -0.6094117 | 0.54310008 | 1.73994234 |
| olink_PLXNA4_Q9HCM2_OID00982 | 0.17368863 | 0.86232569 | 0.41832445 |
| olink_SH2B3_Q9UQQ2_OID00983 | 2.26999216 | 0.02451823 | 1.09978077 |
| olink_FCRL3_Q96P31_OID00984 | -1.0571199 | 0.29202104 | 0.31933833 |
| olink_CKAP4_Q07065_OID00985 | 1.71497427 | 0.08825036 | -1.33999 |
| olink_JUN_P05412_OID00986 | 2.87065514 | 0.00464018 | -0.3114773 |
| olink_HEXIM1_O94992_OID00987 | 3.29226474 | 0.00121863 | -2.8400537 |
| olink_CLEC4D_Q8WXI8_OID00988 | 1.4488066 | 0.14931262 | -2.1360524 |
| olink_PRKCQ_Q04759_OID00989 | 0.46492138 | 0.64260823 | 1.82618917 |
| olink_MGMT_P16455_OID00990 | -0.2649395 | 0.79139084 | -1.3306202 |
| olink_TREM1_Q9NP99_OID00991 | 0.59320386 | 0.55386692 | -2.9638612 |
| olink_CXADR_P78310_OID00992 | -1.0120067 | 0.31303472 | -1.4756925 |
| olink_IL10_P22301_OID00993 | 0.38813218 | 0.69842465 | -2.0944191 |
| olink_SRPK2_P78362_OID00994 | 1.29534349 | 0.1970333 | -1.7606126 |
| olink_KLRD1_Q13241_OID00995 | 0.1409887 | 0.88805296 | -0.2103963 |
| olink_BACH1_O14867_OID00996 | 2.54599645 | 0.01182365 | -0.6717222 |
| olink_PIK3AP1_Q6ZUJ8_OID00997 | 0.10804195 | 0.91409529 | -1.7189389 |
| olink_SPRY2_O43597_OID00998 | -0.6214618 | 0.53516402 | -0.6150703 |
| olink_STC1_P52823_OID00999 | 1.86473213 | 0.06401681 | -3.0349884 |
| olink_ARNT_P27540_OID01000 | 1.92741192 | 0.05566721 | -0.7969364 |
| olink_FAM3B_P58499_OID01001 | -0.3678427 | 0.71346775 | 0.75111095 |
| olink_SH2D1A_O60880_OID01002 | -0.2059092 | 0.83711877 | -0.3704281 |
| olink_ICA1_Q05084_OID01003 | 0.66726587 | 0.50554586 | -1.4090757 |
| olink_DFFA_O00273_OID01004 | 2.49498121 | 0.01359216 | -2.4575599 |
| olink_DCBLD2_Q96PD2_OID01005 | -1.5053992 | 0.13415683 | -2.4668493 |
| olink_FCRL6_Q6DN72_OID01006 | -1.336141 | 0.18336553 | 0.27943235 |
| olink_NCR1_O76036_OID01007 | -0.3976446 | 0.69141241 | -0.0210474 |
| olink_CXCL12_P48061_OID01008 | -0.7610522 | 0.4477255 | 0.70600702 |
| olink_AREG_P15514_OID01009 | 0.34247237 | 0.73243665 | -3.5434876 |
| olink_IFNLR1_Q8IU57_OID01010 | 1.03423085 | 0.30256024 | -1.6449036 |
| olink_DAPP1_Q9UN19_OID01011 | 0.85427386 | 0.39420704 | -0.0545704 |

| | | | |
|---|---|---|---|
| olink_PADI2_Q9Y2J8_OID01012 | -0.0293104 | 0.97665289 | -0.1010687 |
| olink_SIT1_Q9Y3P8_OID01013 | -1.3735765 | 0.1714594 | 0.19811264 |
| olink_MASP1_P48740_OID01014 | -1.1194495 | 0.26459531 | -1.889531 |
| olink_LAMP3_Q9UQV4_OID01015 | 2.73009422 | 0.00702803 | -0.5745998 |
| olink_CLEC7A_Q9BXN2_OID01016 | -0.1109379 | 0.91180209 | 2.73361987 |
| olink_CLEC6A_Q6EIG7_OID01017 | -1.08709 | 0.27860196 | 0.25483259 |
| olink_DDX58_O95786_OID01018 | 3.36283626 | 0.00096147 | -4.1352428 |
| olink_IL12RB1_P42701_OID01019 | 0.26210209 | 0.79357367 | -0.8273517 |
| olink_TANK_Q92844_OID01020 | 0.60802721 | 0.54401568 | -0.9164605 |
| olink_ITGA11_Q9UKX5_OID01021 | -0.3851063 | 0.70066069 | 1.68484549 |
| olink_KPNA1_P52294_OID01022 | -1.9473449 | 0.05321184 | 0.03508771 |
| olink_LAG3_P18627_OID01023 | 2.48150666 | 0.01409703 | 0.2128076 |
| olink_IL5_P05113_OID01024 | 0.12965158 | 0.89700195 | 1.40074273 |
| olink_CD83_Q01151_OID01025 | -1.3614957 | 0.17523606 | -2.0545527 |
| olink_ITGB6_P18564_OID01026 | -0.4926128 | 0.62294908 | -1.0052191 |
| olink_BTN3A2_P78410_OID01027 | 1.32021581 | 0.18861388 | -2.0245663 |

| viral_p | tcell_t | tcell_p | Ab_28days_t | Ab_28days_p | Ab_7M_t |
|---|---|---|---|---|---|
| 0.10955198 | 3.50134717 | 0.00062661 | 2.285656016 | 0.024044241 | 1.113087151 |
| 0.00115954 | 4.04049789 | 8.88E-05 | 3.396685596 | 0.000927953 | 2.206877888 |
| 0.52379523 | 3.4096549 | 0.00085641 | 1.860555867 | 0.065275456 | 0.333584645 |
| 0.00649819 | 2.42594323 | 0.01657945 | 2.519720359 | 0.013070124 | 2.337392183 |
| 0.95639588 | 1.73526485 | 0.08495913 | 0.393083911 | 0.694960811 | 0.582322095 |
| 0.87720062 | 1.30851048 | 0.19290723 | 0.467573952 | 0.640944415 | 0.023538928 |
| 0.24795263 | -0.238491 | 0.81185928 | -0.3820336 | 0.703117822 | 0.129684027 |
| 0.41963557 | 1.5510659 | 0.12320988 | -0.984427169 | 0.32690282 | 0.991630869 |
| 0.25464514 | 2.28146902 | 0.0240742 | 1.144491286 | 0.25471674 | 1.395710115 |
| 0.10546054 | 2.17933907 | 0.03102852 | 0.08732975 | 0.93055619 | 0.485548155 |
| 0.48394517 | 1.68897075 | 0.09351649 | 0.958004016 | 0.340003041 | 1.18629392 |
| 0.06357548 | 1.10234655 | 0.27225797 | 0.074960104 | 0.940372322 | 1.097704763 |
| 0.56870754 | 2.18172584 | 0.03084794 | -0.326260328 | 0.744800478 | 1.4862242 |
| 0.13821222 | 2.93773513 | 0.00388464 | 1.330694349 | 0.185833806 | 1.436351689 |
| 0.10546054 | 2.17933907 | 0.03102852 | 0.08732975 | 0.93055619 | 0.485548155 |
| 0.00021898 | 2.43962457 | 0.01599055 | 4.166532605 | 5.89E-05 | 1.782298671 |
| 0.00761546 | 3.09834876 | 0.00236493 | 2.997147245 | 0.003317789 | 1.094183814 |
| 0.0075853 | 1.52914109 | 0.12855206 | 2.269929746 | 0.025011716 | -0.303586963 |
| 0.63772639 | 1.4966817 | 0.13679326 | 1.411500505 | 0.160706396 | -0.220019732 |
| 0.94072648 | 0.65155821 | 0.51578573 | -0.510816491 | 0.610425918 | 0.223558452 |
| 0.00038476 | 2.8514238 | 0.0050317 | 3.320088984 | 0.00119499 | 2.101534381 |
| 0.00298541 | 2.69048581 | 0.00802953 | 3.936503077 | 0.000139677 | 2.407211708 |
| 0.00093437 | 1.27588297 | 0.20417169 | 3.737088284 | 0.000287455 | 1.538892595 |
| 0.88477146 | 0.0731789 | 0.94177127 | 3.711981482 | 0.000314229 | 1.153006528 |
| 0.23586664 | 1.70910502 | 0.08971245 | -0.522822685 | 0.60206972 | 0.805464831 |
| 0.00958836 | 3.94068973 | 0.00012935 | 1.230250993 | 0.221029575 | 0.294425853 |
| 7.74E-05 | 3.13286746 | 0.00212038 | 3.544295226 | 0.000563578 | 2.019184083 |
| 0.15360579 | 2.15194319 | 0.03316775 | 2.230104272 | 0.027615883 | 1.603110219 |
| 0.45585939 | 1.06544546 | 0.28856228 | -1.816164554 | 0.07186251 | 0.702850171 |
| 0.55848241 | 0.65500916 | 0.5135682 | -1.370384239 | 0.173146602 | -0.411094396 |
| 0.85388816 | 2.3038897 | 0.0227447 | 3.236954671 | 0.001565241 | 1.203632133 |
| 0.8741747 | 1.50118198 | 0.13562665 | 1.703061313 | 0.091167048 | 0.616013281 |
| 0.82758324 | 0.94977601 | 0.34391083 | -0.870872682 | 0.385577136 | -0.293251073 |
| 0.93506407 | -0.590094 | 0.5561068 | -1.447290952 | 0.150444702 | -2.098952044 |
| 0.60019161 | 1.70527007 | 0.09042712 | -1.341780211 | 0.18222202 | 0.781273737 |
| 0.47889067 | 2.67149137 | 0.00847384 | 1.484300517 | 0.140373226 | 0.644184747 |
| 0.14064675 | 2.68792285 | 0.00808823 | 0.591164264 | 0.555531603 | 1.842779206 |
| 0.78273757 | 2.52711432 | 0.0126452 | 1.18401163 | 0.238768082 | 0.380503932 |
| 0.60755446 | 1.03721316 | 0.30147717 | -1.01098219 | 0.314076173 | -0.224640989 |
| 0.78273757 | 2.52711432 | 0.0126452 | 1.18401163 | 0.238768082 | 0.380503932 |
| 0.14445214 | 3.05461549 | 0.00271225 | 0.599511333 | 0.549971707 | 1.616375446 |

| | | | | | |
|---|---|---|---|---|---|
| 0.42856324 | 0.4880369 | 0.6263096 | -0.676087478 | 0.500296971 | 0.11949252 |
| 0.47468879 | 1.52021969 | 0.13077727 | -0.46737206 | 0.641088408 | 0.618897336 |
| 0.42748755 | 1.52993598 | 0.12835525 | -0.511323067 | 0.610072293 | 0.588739601 |
| 0.64495308 | 0.43390362 | 0.66504597 | -0.703240256 | 0.483279633 | 0.16884111 |
| 0.43494335 | 1.03403964 | 0.30295286 | -0.882827526 | 0.379109398 | -0.305168784 |
| 0.50749416 | 0.85613794 | 0.3934274 | -1.354938737 | 0.178003687 | -0.445059169 |
| 0.0131808 | 2.82077764 | 0.00550838 | 3.101945387 | 0.002401336 | 1.336739754 |
| 0.00301432 | 2.55340039 | 0.01177049 | 3.227652716 | 0.001612748 | 1.557050806 |
| 0.62922277 | -0.1324255 | 0.89484349 | 2.576108183 | 0.011214936 | -0.116546514 |
| 0.11307058 | 0.40340798 | 0.68728165 | -0.08958402 | 0.928768385 | -1.555808237 |
| 0.36589978 | 2.61473889 | 0.00993685 | 3.456865322 | 0.000758586 | 1.110911663 |
| 0.49017782 | 3.21478973 | 0.00163077 | 2.244161343 | 0.02667084 | 0.218328144 |
| 0.15622373 | 2.52825271 | 0.01260614 | 0.548221112 | 0.584567055 | 1.042153802 |
| 0.75939381 | 1.4558256 | 0.14774541 | -1.140738753 | 0.25626922 | -0.332321389 |
| 0.10659503 | 2.90239659 | 0.00432165 | 0.6984539 | 0.486256077 | 0.832968942 |
| 0.37961604 | 2.90451193 | 0.00429428 | 0.296368036 | 0.767465641 | 0.603699806 |
| 0.64354656 | 2.06783303 | 0.04055098 | 0.822796576 | 0.412269424 | -0.417826622 |
| 0.00024485 | 3.69123269 | 0.00032201 | 3.303907324 | 0.001259922 | 2.165769319 |
| 0.51331136 | 0.73291949 | 0.46486891 | -1.434758728 | 0.153978761 | 0.997018343 |
| 0.15089709 | 0.21518291 | 0.82994707 | -2.542991455 | 0.012273513 | -0.48940335 |
| 0.00749268 | 1.07439786 | 0.2845469 | 0.125135306 | 0.900627584 | 1.572898447 |
| 0.23305904 | 4.37648437 | 2.38E-05 | 3.579722305 | 0.000498916 | 1.584121614 |
| 0.08408189 | 4.45048467 | 1.77E-05 | 3.439354204 | 0.000804604 | 2.090543149 |
| 0.52336837 | 1.60234985 | 0.11139838 | 2.356917312 | 0.020060494 | 1.015154589 |
| 0.30550436 | 3.08953141 | 0.00243146 | 0.180562059 | 0.857018482 | 0.690217003 |
| 0.002175 | 4.59792947 | 9.65E-06 | 3.681680951 | 0.000349693 | 1.554531205 |
| 0.00352347 | 2.35616617 | 0.0198925 | 2.473518976 | 0.014790162 | 2.309845043 |
| 0.58323895 | 1.28591561 | 0.2006576 | -0.572301373 | 0.568197439 | 2.459299798 |
| 0.85325195 | 1.78810524 | 0.0759856 | 2.075396414 | 0.04010441 | 0.384573979 |
| 0.15800656 | 1.37605123 | 0.17106748 | 2.322708711 | 0.021893649 | 0.433905718 |
| 0.32536237 | -0.338678 | 0.73537465 | -2.081255178 | 0.039555171 | 0.313258758 |
| 0.08010606 | 2.96234019 | 0.00360474 | -0.300881267 | 0.764030101 | 1.372663031 |
| 0.56933394 | 4.1248332 | 6.43E-05 | 3.159460461 | 0.00200426 | 1.98971027 |
| 0.98155059 | 1.51442469 | 0.13223879 | 0.946373729 | 0.345875723 | 0.7472734 |
| 0.21470834 | 1.5009003 | 0.13569944 | 0.10552537 | 0.916136611 | 1.435394519 |
| 0.91886963 | -1.1968491 | 0.23344752 | 1.066891507 | 0.288180531 | 1.216224563 |
| 0.09572928 | 1.68112973 | 0.09503288 | 1.097102858 | 0.274811519 | 1.77343108 |
| 0.4249453 | -0.8453858 | 0.3993803 | -1.978108145 | 0.050227191 | 0.235782886 |
| 0.12962158 | 3.78977671 | 0.00022572 | 2.523412171 | 0.012940734 | 0.649866556 |
| 0.06773547 | 2.98752391 | 0.00333756 | 0.77495909 | 0.439901005 | 1.138941745 |
| 0.44852104 | 3.24380761 | 0.00148421 | 2.273926602 | 0.024762649 | 0.157464411 |
| 0.27907057 | 2.72998815 | 0.00717238 | 0.661634919 | 0.509484345 | 0.212709178 |

| | | | | | |
|---|---|---|---|---|---|
| 0.92169952 | 2.57847701 | 0.01098702 | 1.212704358 | 0.227644845 | 0.283452895 |
| 0.82995873 | 1.5710574 | 0.11849297 | 1.077181485 | 0.283578032 | 1.555974279 |
| 0.03473989 | 4.27970858 | 3.51E-05 | 3.618955305 | 0.000435503 | 1.272303676 |
| 0.00020726 | 4.12196089 | 6.50E-05 | 2.056824938 | 0.041888918 | 1.777235466 |
| 0.0074258 | 3.51894242 | 0.00058974 | 0.30976694 | 0.757279981 | 0.654709798 |
| 0.82634819 | -1.4485206 | 0.14977317 | -1.40464841 | 0.162730715 | -0.455550246 |
| 0.05203646 | 2.79411769 | 0.0059562 | 1.976030249 | 0.050465143 | 1.161365765 |
| 0.40581934 | 0.02945791 | 0.97654258 | -3.424961138 | 0.00084438 | -1.005917101 |
| 0.02956498 | 3.56182871 | 0.00050826 | 1.826005214 | 0.070356587 | 1.776049993 |
| 0.77854323 | 0.55933583 | 0.57685251 | -0.308497319 | 0.758243332 | 0.138835879 |
| 0.30253171 | 1.79429756 | 0.07498715 | 0.005250574 | 0.99581946 | -0.020285121 |
| 0.71925445 | 2.37270409 | 0.01905841 | 1.208882051 | 0.229104658 | 0.211312089 |
| 0.00063883 | 3.22722217 | 0.0015664 | 3.698869246 | 0.000329136 | 2.514677978 |
| 0.03612283 | 2.6430726 | 0.00918017 | 0.68068941 | 0.497390347 | 1.35083455 |
| 0.40238338 | 2.69937307 | 0.00782898 | 1.682884736 | 0.095019341 | 1.860637976 |
| 0.00908636 | 3.84924292 | 0.00018159 | 2.56080636 | 0.011693412 | 0.779574546 |
| 0.04885007 | 3.49073264 | 0.00064988 | 2.336983347 | 0.021111481 | 1.739279035 |
| 0.47891417 | 2.64835135 | 0.00904508 | 2.410464901 | 0.017462783 | 0.963812525 |
| 0.05185971 | 2.83958738 | 0.00521115 | 0.356667979 | 0.72197202 | 0.14544742 |
| 0.01549943 | 3.37244455 | 0.00097053 | 2.380371329 | 0.018883342 | 2.297865031 |
| 0.67415086 | -1.031758 | 0.30401683 | -1.017891297 | 0.310794628 | -1.315570945 |
| 0.26693364 | 3.07798256 | 0.00252121 | 1.586883947 | 0.115192603 | -0.15081676 |
| 0.00293862 | 2.5408041 | 0.01218265 | 0.928028368 | 0.355271366 | 1.454058199 |
| 0.11827141 | 2.62319065 | 0.00970546 | 2.491547218 | 0.014096297 | 0.254866252 |
| 0.34780227 | 2.45792765 | 0.01523181 | 1.596745594 | 0.112974627 | -0.347968378 |
| 0.39340728 | -0.5810049 | 0.56219878 | -2.051646602 | 0.042398471 | -0.147306258 |
| 0.42387666 | 2.61987263 | 0.00979571 | 1.724407818 | 0.087230805 | -0.194510169 |
| 0.00293862 | 2.5408041 | 0.01218265 | 0.928028368 | 0.355271366 | 1.454058199 |
| 0.95471183 | -0.9232617 | 0.35750596 | 1.295276455 | 0.197732049 | -0.544579163 |
| 0.0511932 | 1.67665489 | 0.09590716 | 2.559564271 | 0.011733044 | 1.850129544 |
| 0.59025851 | 2.00697551 | 0.04673445 | -0.741195483 | 0.460035083 | 0.527468963 |
| 0.63972396 | 2.03886774 | 0.04340066 | -0.833836499 | 0.406043755 | 0.590240057 |
| 0.02123749 | 1.75366609 | 0.08174053 | 2.337667801 | 0.021074604 | 0.499137924 |
| 0.24614179 | 2.04738237 | 0.04254573 | 0.795143276 | 0.428113258 | -0.47805643 |
| 0.0124273 | 4.20972954 | 4.62E-05 | 2.790096507 | 0.006138627 | 1.310276193 |
| 0.04989694 | 2.27197865 | 0.02465718 | 1.894683876 | 0.060561035 | 3.944241094 |
| 0.00070785 | 3.47637459 | 0.00068266 | 2.378666159 | 0.018966815 | -0.16704362 |
| 0.09327175 | 3.32298739 | 0.00114433 | 1.119620733 | 0.265130152 | 1.856317168 |
| 0.06049273 | 1.08888474 | 0.27813048 | 0.248643674 | 0.804065025 | -0.768107454 |
| 0.6797076 | -0.473358 | 0.63671671 | 0.327308433 | 0.744009691 | -0.89211348 |
| 0.06049273 | 1.08888474 | 0.27813048 | 0.248643674 | 0.804065025 | -0.768107454 |
| 0.09879863 | 3.44606534 | 0.00075703 | 1.055057417 | 0.293536424 | 1.696392783 |

| | | | | | |
|---|---|---|---|---|---|
| 0.58381002 | 2.04158834 | 0.04312591 | 1.264141759 | 0.208649673 | 0.972584931 |
| 0.9916265 | 1.62734291 | 0.10597894 | -0.02446447 | 0.980523101 | 0.352500728 |
| 0.6265312 | 2.62741379 | 0.00959166 | 0.025120294 | 0.980001089 | 0.34795083 |
| 0.51668428 | 1.81533326 | 0.07167593 | -0.305028879 | 0.760877017 | 0.617467738 |
| 0.44064316 | 2.0822523 | 0.03919319 | 1.604147295 | 0.111332409 | -0.048593771 |
| 0.14678912 | -1.7137863 | 0.0888463 | -5.651397847 | 1.11E-07 | -1.206973696 |
| 0.31900613 | 3.38121114 | 0.00094243 | 2.282498811 | 0.024235792 | 0.251079888 |
| 0.23880618 | 2.9964191 | 0.00324763 | 0.355387541 | 0.722928432 | -0.167741347 |
| 0.45212584 | 3.34582206 | 0.00106075 | 2.238477192 | 0.027049519 | 0.231691934 |
| 0.0441679 | 3.194687 | 0.00174008 | 0.700423474 | 0.485030063 | 1.157857683 |
| 0.51971967 | 1.55958159 | 0.12118282 | -0.465946051 | 0.642105845 | 1.142122781 |
| 0.95906525 | 0.37179108 | 0.71062704 | -0.419190207 | 0.675832846 | 0.756887895 |
| 0.92606128 | 1.8185742 | 0.0711767 | 0.604335757 | 0.546770917 | -0.552059589 |
| 0.3176151 | 3.46115178 | 0.00071911 | 0.798891926 | 0.425944688 | 1.94319155 |
| 0.62164354 | 0.43818053 | 0.66195062 | -1.748989698 | 0.082871182 | 1.056528045 |
| 0.05583979 | -2.6980835 | 0.0078578 | -4.76653852 | 5.35E-06 | -1.325342472 |
| 0.06527223 | -2.144965 | 0.03373263 | -4.69731953 | 7.13E-06 | -1.210759571 |
| 0.48937423 | 3.19805351 | 0.00172131 | 2.244411022 | 0.026654313 | 0.209981914 |
| 0.34506602 | 0.74438208 | 0.45792963 | 0.933070263 | 0.35267303 | -0.150317103 |
| 0.24045494 | 2.20935205 | 0.02882335 | 0.493290344 | 0.622716566 | 1.033738679 |
| 0.47011367 | 2.26336976 | 0.02519667 | 1.768701553 | 0.07950586 | 0.968674299 |
| 0.45493 | 3.26713799 | 0.00137537 | 2.289109865 | 0.023836216 | 0.18848208 |
| 0.86948223 | -0.1424665 | 0.88692241 | -3.052851281 | 0.002796721 | -1.979264644 |
| 0.06106176 | 2.77306014 | 0.00633309 | 1.476475051 | 0.142457798 | -0.372471479 |
| 0.55339642 | 2.2926325 | 0.02340394 | 1.264993613 | 0.208345193 | 0.677883614 |
| 0.89972967 | 2.3939797 | 0.01803094 | 1.142347741 | 0.255602744 | -0.128094226 |
| 0.44912277 | 3.30995515 | 0.00119474 | 2.290530787 | 0.023751094 | 0.185523233 |
| 0.14616928 | 2.66020922 | 0.00874815 | 0.710600741 | 0.478722051 | 0.85388109 |
| 0.32700894 | 0.50094614 | 0.61721884 | 1.572534519 | 0.11848171 | -0.557727355 |
| 0.97605512 | 1.77482493 | 0.07816393 | 0.824389252 | 0.411367755 | -0.013117834 |
| 0.8282347 | 2.01512806 | 0.04586215 | 2.407197755 | 0.017612277 | 0.030940482 |
| 0.16911272 | 2.60953922 | 0.01008166 | -0.606626304 | 0.545254517 | 0.485119335 |
| 0.2317809 | 2.66109279 | 0.00872638 | 2.207932343 | 0.029165941 | 0.955315796 |
| 0.21290206 | 3.41714441 | 0.00083502 | 1.01407052 | 0.3126065 | 0.29553147 |
| 0.6596884 | 1.43952523 | 0.15229954 | 0.550310025 | 0.583138262 | 1.1070199 |
| 0.27147715 | 3.24911199 | 0.00145878 | 2.426577071 | 0.016741898 | 1.495206975 |
| 0.06484547 | 0.97806061 | 0.32978094 | -0.972199425 | 0.332923351 | 0.715595041 |
| 0.25341846 | 2.2531208 | 0.02585238 | 0.416570096 | 0.677743304 | 0.879388544 |
| 0.32640053 | -1.7653667 | 0.07974647 | -2.619235858 | 0.009959672 | -0.664744576 |
| 0.01726874 | -1.9677989 | 0.05112563 | -3.37058023 | 0.001011947 | -1.385359653 |
| 0.07911047 | 0.36089734 | 0.71873617 | -1.93039005 | 0.05593788 | 0.212463501 |
| 0.54832097 | 1.46186608 | 0.14608475 | -0.704293777 | 0.482625838 | 0.233984044 |

| | | | | | |
|---|---|---|---|---|---|
| 0.2272403 | 1.0480917 | 0.2964554 | 0.084239241 | 0.933007771 | -1.441492469 |
| 0.85810036 | 1.72979718 | 0.08593522 | 2.022287869 | 0.045388853 | 1.988723268 |
| 0.68026174 | 3.33496751 | 0.00109974 | 2.14072073 | 0.03433682 | 1.960443026 |
| 0.12806891 | 2.31151308 | 0.02230757 | 2.228402948 | 0.027732223 | 2.000575363 |
| 0.02939273 | 1.31267388 | 0.19150371 | 1.09754528 | 0.27461898 | 0.868958185 |
| 0.03107069 | 1.76138437 | 0.08042061 | -0.598052318 | 0.550941537 | 0.766889743 |
| 0.99068409 | 1.79819227 | 0.07436473 | -0.743844599 | 0.458436703 | 0.157534401 |
| 0.40581934 | 0.02945791 | 0.97654258 | -3.424961138 | 0.00084438 | -1.005917101 |
| 0.20868879 | 3.23439479 | 0.00153035 | 1.69327791 | 0.093018803 | 0.599661539 |
| 0.75627606 | 0.45381344 | 0.65068646 | -0.362535717 | 0.717594812 | 0.008495804 |
| 0.42769649 | 2.94785115 | 0.00376723 | 1.559818739 | 0.121458288 | 0.343326183 |
| 0.91384909 | 1.21076396 | 0.22808578 | -1.113849486 | 0.267588476 | -0.22982105 |
| 0.76594964 | -0.8490313 | 0.39735589 | -4.521146499 | 1.46E-05 | -1.295534561 |
| 0.91093027 | 4.22487566 | 4.35E-05 | 1.887783471 | 0.061490499 | 0.992183728 |
| 0.84227542 | 2.364055 | 0.01949069 | 0.868679909 | 0.386770825 | -0.036124125 |
| 0.05598577 | -0.3409644 | 0.73365654 | -0.792464092 | 0.429667128 | 0.251313987 |
| 0.65601883 | 0.40386715 | 0.68694476 | 0.071840063 | 0.942849791 | 1.519989006 |
| 0.95135982 | 1.6985467 | 0.09169126 | 1.025760951 | 0.30708487 | 1.698076522 |
| 0.38879635 | 2.09063097 | 0.03842229 | 1.084145393 | 0.28049197 | 0.463469798 |
| 0.85919299 | 2.19365769 | 0.02995884 | 0.503910851 | 0.615255666 | 1.510287602 |
| 0.10323847 | 1.43807919 | 0.15270871 | -3.022680772 | 0.003068722 | 0.940548866 |
| 0.03722121 | -3.3896373 | 0.00091614 | -5.323906185 | 4.86E-07 | -1.996575402 |
| 0.130172 | -2.0535567 | 0.04193483 | -3.520734884 | 0.000610872 | -1.020485554 |
| 0.03250485 | -1.0106444 | 0.31398137 | -1.290766593 | 0.199286721 | 0.540995407 |
| 0.19162454 | 1.59822005 | 0.1123148 | 2.178531295 | 0.031337969 | -0.409669984 |
| 0.42414512 | 3.39897166 | 0.00088782 | 2.358284733 | 0.019990123 | 0.194107702 |
| 0.1259727 | 2.17807033 | 0.03112488 | 2.084978641 | 0.03920948 | -0.656844047 |
| 0.06878395 | -0.1846109 | 0.85380923 | 1.115061291 | 0.267070983 | -0.006456567 |
| 0.03130548 | 0.05250805 | 0.95820093 | 1.529538079 | 0.12878503 | 0.374330792 |
| 0.08385239 | -0.37504 | 0.70821492 | 0.722415168 | 0.471456571 | 0.136257507 |
| 0.61329996 | 2.06002902 | 0.04130252 | 0.311263344 | 0.756145043 | 0.946840507 |
| 0.42370518 | 1.98473583 | 0.04918611 | 2.584885542 | 0.010948491 | 0.00161222 |
| 0.06385256 | 2.04151181 | 0.04313362 | 2.273563654 | 0.024785177 | -0.241979874 |
| 0.2108422 | -0.1511571 | 0.88007576 | 0.659459188 | 0.510875146 | 0.253024171 |
| 0.05105651 | -2.0088319 | 0.04653458 | -4.080304959 | 8.17E-05 | -1.253980012 |
| 0.01350415 | 0.42549054 | 0.67115165 | 0.887164006 | 0.376780105 | 0.219355958 |
| 0.00277114 | 2.60733836 | 0.01014352 | -1.619895734 | 0.107901789 | -0.67987035 |
| 0.15076868 | 1.56592985 | 0.11968893 | -0.457870333 | 0.647880554 | 1.936340418 |
| 0.69308261 | 2.32274605 | 0.02167693 | 0.456677195 | 0.648735564 | 0.248468897 |
| 0.54815428 | 3.33789648 | 0.00108909 | 2.866947548 | 0.004903145 | 1.069164647 |
| 0.13712048 | 2.37224826 | 0.01908098 | 2.30490954 | 0.022904631 | 2.055656824 |
| 0.54723268 | 0.38207309 | 0.70300352 | -2.803173875 | 0.005910131 | -0.476926614 |

| | | | | | |
|---|---|---|---|---|---|
| 0.11841553 | 6.48443892 | 1.52E-09 | 5.118043526 | 1.20E-06 | 3.155191826 |
| 0.05942008 | 5.90589089 | 2.66E-08 | 3.366780916 | 0.001024749 | 3.75168047 |
| 0.00203837 | 2.64231073 | 0.00919981 | 0.648891749 | 0.517658685 | 0.35408239 |
| 0.00204236 | 6.61336387 | 7.89E-10 | 6.369396212 | 3.72E-09 | 1.873211007 |
| 0.6365687 | 1.1612996 | 0.24755427 | 2.624161425 | 0.009824687 | 0.33247932 |
| 0.15300212 | 2.28818855 | 0.02366877 | 5.057427632 | 1.56E-06 | 1.2860152 |
| 0.17089936 | -0.0093685 | 0.99253886 | 0.166696095 | 0.867892094 | 0.182808073 |
| 0.84650197 | -0.0735299 | 0.94149252 | 1.478435104 | 0.141933427 | -0.431298796 |
| 0.61124998 | 2.27460885 | 0.02449438 | 3.775112942 | 0.000250989 | 2.728540387 |
| 0.55449606 | 2.97103204 | 0.00351038 | 1.049987484 | 0.295851547 | 1.666980842 |
| 0.23382736 | 1.60092709 | 0.11171342 | 3.414501524 | 0.000874435 | 0.69168629 |
| 0.00343918 | 6.38949166 | 2.46E-09 | 7.040133571 | 1.33E-10 | 2.391829779 |
| 0.43803989 | 0.9637197 | 0.33689694 | 0.233434887 | 0.815824804 | -0.904827607 |
| 0.01265915 | 6.30586274 | 3.74E-09 | 3.655230876 | 0.000383718 | 3.115248133 |
| 0.55444207 | -0.6858755 | 0.4939592 | 0.521040321 | 0.603306924 | -1.576971002 |
| 0.31978304 | 6.03500588 | 1.42E-08 | 4.428023482 | 2.12E-05 | 3.346509875 |
| 0.4885898 | 2.74037817 | 0.00696116 | 2.50464193 | 0.013610703 | 2.158791165 |
| 0.00013503 | -0.4152873 | 0.67858606 | -1.362318173 | 0.175670442 | -0.464353924 |
| 0.86774929 | 0.27275794 | 0.78545301 | -0.375524294 | 0.707939121 | -0.317027659 |
| 0.03460333 | 5.79958645 | 4.44E-08 | 4.684902246 | 7.51E-06 | 1.586944007 |
| 0.97943618 | -0.2330345 | 0.81608495 | 0.688791166 | 0.492295412 | 0.261041208 |
| 0.98628999 | -1.8295084 | 0.06951365 | -0.189767308 | 0.84981479 | -0.294381599 |
| 0.2737305 | -0.4088405 | 0.68329977 | 0.722572078 | 0.471360493 | -1.707308579 |
| 0.17659781 | 2.76033549 | 0.00657123 | 5.065271076 | 1.51E-06 | 1.279287233 |
| 0.40059497 | 0.42520388 | 0.67136008 | -0.111487298 | 0.911417765 | -2.31526964 |
| 0.32463152 | 5.90145755 | 2.72E-08 | 3.223503356 | 0.00163437 | 1.552660453 |
| 0.23324747 | 0.92157679 | 0.35838131 | 2.140352081 | 0.03436724 | -0.565854876 |
| 0.86343158 | 4.95608178 | 2.10E-06 | 3.875800678 | 0.000174462 | 3.127108955 |
| 0.04515349 | -2.5907701 | 0.01062032 | 0.638906633 | 0.524111518 | -0.597736207 |
| 0.00029332 | -2.9844241 | 0.00336943 | -4.237002811 | 4.49E-05 | -1.725296226 |
| 0.70142687 | 2.76448244 | 0.00649274 | 2.321655987 | 0.021952331 | 3.053641484 |
| 0.96056079 | 1.73598289 | 0.08483162 | 4.018703917 | 0.000102978 | 1.907243881 |
| 0.26771506 | 1.8601715 | 0.06502124 | 3.629780251 | 0.000419396 | -0.198405334 |
| 0.77065479 | 2.65375924 | 0.00890855 | 0.90297718 | 0.368362199 | 2.474066041 |
| 0.07968047 | 2.53235888 | 0.01246617 | 2.240493668 | 0.026914647 | 2.135959245 |
| 0.03283866 | 2.59131747 | 0.01060426 | 5.772094816 | 6.35E-08 | 2.209230645 |
| 0.41919368 | -1.8616016 | 0.06481777 | 0.600877301 | 0.5490645 | -0.846290438 |
| 0.74501659 | -1.914003 | 0.05772054 | -0.691104135 | 0.490846077 | -2.85780258 |
| 0.5112433 | 0.76370806 | 0.44636411 | 0.722985434 | 0.471107442 | -0.344594321 |
| 0.28894153 | 1.91294189 | 0.05785749 | 0.337168811 | 0.736583601 | -0.014424887 |
| 0.60019802 | 2.33041059 | 0.02125569 | 2.48957888 | 0.014170613 | 1.531148311 |
| 0.14196681 | 2.64777186 | 0.00905982 | 2.542102598 | 0.012303122 | 2.127206541 |

| | | | | | |
|---|---|---|---|---|---|
| 0.0254746 | 0.77759698 | 0.43815722 | 0.561639824 | 0.57541768 | 2.668057848 |
| 0.35187731 | 2.49481592 | 0.01379935 | 1.886594636 | 0.061651834 | 0.953828745 |
| 0.03527415 | -0.2495421 | 0.80331802 | -0.164076262 | 0.869949465 | 0.660807114 |
| 0.66978872 | -2.0826892 | 0.03915267 | 0.765941539 | 0.445227707 | -1.027649868 |
| 0.20024998 | 3.18244142 | 0.00180997 | 4.048010125 | 9.23E-05 | 3.233834653 |
| 0.6206844 | 5.37924814 | 3.17E-07 | 4.334044168 | 3.08E-05 | 2.998964177 |
| 0.69739595 | 0.22343608 | 0.8235314 | -0.057311034 | 0.954393552 | -0.451346225 |
| 0.78865915 | 1.65863089 | 0.09949477 | 2.925372876 | 0.004121163 | 1.210937983 |
| 0.55964697 | -2.5943834 | 0.01051465 | -2.909070372 | 0.00432696 | -1.663514661 |
| 0.08096709 | 3.85826878 | 0.00017565 | 6.923015307 | 2.40E-10 | 2.788996676 |
| 0.49663 | -0.4726107 | 0.63724851 | 2.908727069 | 0.004331394 | 1.710116916 |
| 0.38325741 | 0.00152585 | 0.99878478 | -0.019019951 | 0.984857043 | -0.837971499 |
| 0.6013628 | 1.11410927 | 0.26719738 | 0.259733669 | 0.795518048 | -2.951192451 |
| 0.02815695 | -0.7462532 | 0.45680251 | -0.588661038 | 0.557204376 | -0.253979739 |
| 0.04712567 | 0.35782566 | 0.72102852 | -0.550008443 | 0.583344439 | 1.43611625 |
| 0.83522715 | 2.67795296 | 0.00832026 | 1.313267935 | 0.191619291 | 0.22210895 |
| 0.73976073 | 2.58551933 | 0.01077558 | 3.70364372 | 0.000323632 | 1.887447308 |
| 0.0068686 | 4.21282351 | 4.56E-05 | 1.053532828 | 0.294231311 | 2.396384679 |
| 0.58818598 | -1.1113439 | 0.26838117 | 2.32261416 | 0.021898914 | 1.202536108 |
| 0.29423557 | 6.48565553 | 1.51E-09 | 5.117552477 | 1.20E-06 | 2.620377239 |
| 0.69746111 | -0.4886925 | 0.62584652 | 0.218372033 | 0.827513304 | -0.303077247 |
| 0.34548308 | 5.33274518 | 3.93E-07 | 1.636450115 | 0.104387506 | 1.929546089 |
| 0.04529239 | 5.78484068 | 4.76E-08 | 6.570550952 | 1.39E-09 | 3.762127 |
| 0.14023138 | 3.78732796 | 0.00022774 | 3.650395768 | 0.000390267 | 2.068931696 |
| 0.36004046 | -0.4239153 | 0.67229729 | 0.622960368 | 0.534502621 | -0.814173135 |
| 0.23453583 | 1.53382508 | 0.12739574 | 1.701025241 | 0.091549932 | 1.669945933 |
| 0.96617766 | -0.4636585 | 0.64363362 | -0.077911372 | 0.938029405 | -0.879967099 |
| 0.18596715 | -3.1484449 | 0.00201789 | -2.815825033 | 0.005696494 | -1.525248226 |
| 0.28922403 | 2.83978416 | 0.00520812 | 4.457332692 | 1.89E-05 | 1.60870705 |
| 0.17349037 | 5.15115995 | 8.90E-07 | 3.14343341 | 0.002108293 | 3.114888969 |
| 0.53993209 | 0.26552778 | 0.79100499 | 0.734598545 | 0.464029131 | -1.137108786 |
| 0.07696528 | 3.95444484 | 0.00012286 | 4.286643288 | 3.71E-05 | 1.684483973 |
| 0.9427325 | -2.2823907 | 0.02401823 | -1.423437722 | 0.15722588 | -0.38696278 |
| 0.23666113 | -0.5950072 | 0.5528274 | 1.503324195 | 0.135404734 | -0.918073286 |
| 0.72448065 | -0.4898225 | 0.62504868 | 2.301288569 | 0.023115256 | -0.381511601 |
| 0.40763279 | 0.30748383 | 0.75894549 | -0.553639157 | 0.580864588 | -0.813612356 |
| 0.53824141 | 5.33281387 | 3.92E-07 | 1.806189706 | 0.073416158 | 2.32785062 |
| 0.04096422 | 5.21924404 | 6.56E-07 | 2.853450485 | 0.005102132 | 1.202496211 |
| 0.00011877 | 1.3063085 | 0.19365261 | 0.974533141 | 0.331768744 | 0.890682801 |
| 0.33348282 | 2.55102562 | 0.01184723 | 0.358513309 | 0.720594441 | 1.378315772 |
| 0.02483573 | 0.49707436 | 0.61993923 | -0.913172513 | 0.362998218 | -0.233509291 |
| 0.33732776 | 0.58226441 | 0.56135263 | 0.228122289 | 0.819942693 | -0.519501584 |

| | | | | | |
|---|---|---|---|---|---|
| 0.01476722 | -4.385932 | 2.30E-05 | -2.584257754 | 0.010967357 | -2.381130615 |
| 0.00017444 | 3.66737748 | 0.00035058 | 6.080378186 | 1.49E-08 | 1.989216684 |
| 0.71585763 | -0.1269527 | 0.89916532 | 0.186044428 | 0.852726687 | -0.761774373 |
| 0.15117693 | 4.34026178 | 2.76E-05 | 2.821500487 | 0.005602966 | 2.301503471 |
| 0.01675347 | -1.3731892 | 0.17195326 | -1.72131842 | 0.087791738 | -2.433720454 |
| 0.07347914 | 1.14551747 | 0.2540066 | 3.49048875 | 0.00067708 | 1.675489555 |
| 0.15577941 | -2.857806 | 0.0049373 | -1.578056365 | 0.117207298 | -0.802804954 |
| 0.62644217 | 5.10585112 | 1.09E-06 | 4.07532457 | 8.33E-05 | 3.490445186 |
| 0.77481977 | 2.4226807 | 0.01672269 | 2.89479307 | 0.004514898 | 3.369670517 |
| 0.26924095 | 1.69881811 | 0.09163995 | 0.493984151 | 0.622227955 | 1.697667543 |
| 0.00070268 | 2.291784 | 0.02345431 | 2.148522461 | 0.033698471 | 4.408087225 |
| 0.17696222 | 3.14604197 | 0.0020334 | 1.124550855 | 0.263042629 | 0.84555083 |
| 0.08341244 | 0.43530692 | 0.66402971 | 0.337198427 | 0.736561333 | 0.635188682 |
| 0.07383001 | -0.9326629 | 0.35264687 | -0.618076768 | 0.537705864 | 0.00952734 |
| 0.00505065 | 3.23214458 | 0.00154157 | 5.035888769 | 1.71E-06 | 2.925471024 |
| 0.51926976 | -1.40576 | 0.16207587 | -0.566011498 | 0.572451778 | 0.143894368 |
| 0.03463385 | -0.0115628 | 0.99079137 | -0.109309795 | 0.913140894 | 1.123780087 |
| 0.3566304 | 0.91598518 | 0.36129601 | 1.968128952 | 0.051378721 | 1.78979506 |
| 0.86349111 | 1.23421543 | 0.21925109 | 0.999320116 | 0.319667355 | 2.134805533 |
| 0.00094724 | 7.00673096 | 1.03E-10 | 7.706595683 | 4.30E-12 | 3.03211913 |
| 0.02823216 | 1.2957051 | 0.19727191 | -0.455379344 | 0.649666146 | -0.430791845 |
| 8.52E-05 | -1.3887697 | 0.16717292 | -3.499596805 | 0.000656464 | -1.469978978 |
| 0.14130503 | 3.88316795 | 0.00016022 | 5.117633463 | 1.20E-06 | 2.662091692 |
| 0.75512374 | -2.4665716 | 0.01488476 | -2.594418937 | 0.01066556 | 0.267053917 |
| 0.00986885 | 5.07755722 | 1.23E-06 | 5.101130269 | 1.29E-06 | 3.180921282 |
| 0.0376984 | 1.2627415 | 0.20884289 | 1.364271488 | 0.175056722 | 0.136016435 |
| 0.65358715 | -1.2661655 | 0.20761834 | 0.179100267 | 0.858163554 | -0.625128397 |
| 0.03604184 | 5.38312971 | 3.12E-07 | 5.695832006 | 9.03E-08 | 3.368929902 |
| 0.88294943 | -0.6937231 | 0.48903907 | 0.089806303 | 0.928592117 | -0.528221695 |
| 0.08629057 | 2.81501129 | 0.00560256 | 3.427157141 | 0.000838194 | 3.097185517 |
| 0.81276079 | 2.71577722 | 0.00747068 | 2.101863314 | 0.037674299 | 2.265373291 |
| 0.50607 | -0.620278 | 0.53611274 | 0.206162981 | 0.837015907 | -0.68274063 |
| 0.84197774 | -0.1854923 | 0.8531193 | -2.570203242 | 0.011397452 | -0.747534451 |
| 0.69956403 | 0.49485083 | 0.62150392 | 0.26608885 | 0.79063126 | 0.053598056 |
| 0.87078514 | 1.18998642 | 0.23612491 | 1.519512994 | 0.131285918 | 1.253712093 |
| 0.88873411 | 2.04585081 | 0.04269844 | 1.916026785 | 0.057760533 | 1.688593956 |
| 0.00328838 | 2.33353202 | 0.02108621 | 4.364687574 | 2.73E-05 | 3.275206588 |
| 0.01375575 | 6.47442725 | 1.60E-09 | 6.226105246 | 7.43E-09 | 5.603498418 |
| 0.74484591 | -1.7053811 | 0.09040637 | 1.105652151 | 0.271107419 | 0.368498281 |
| 0.00050365 | 6.79126292 | 3.16E-10 | 3.776843319 | 0.000249439 | 2.639277436 |
| 0.47966099 | 2.38165803 | 0.01861985 | 1.131660317 | 0.260052599 | 2.965233789 |
| 0.00392297 | 2.93793117 | 0.00388233 | 6.095999956 | 1.39E-08 | 4.637865138 |

| | | | | | |
|---|---|---|---|---|---|
| 0.03511023 | 2.19856527 | 0.02959967 | 2.365548729 | 0.019619948 | 2.152183145 |
| 0.03337826 | -0.3456356 | 0.73015062 | 3.373464661 | 0.001002328 | 2.549792514 |
| 0.03887164 | 2.00319758 | 0.04714344 | 3.283053338 | 0.001348471 | 2.694617386 |
| 0.45725736 | -0.5395187 | 0.59041122 | -0.719999303 | 0.472937222 | 0.827653268 |
| 0.44944397 | -1.2074994 | 0.22933566 | -0.960902415 | 0.338549636 | 1.299305014 |
| 0.75332949 | -1.5569111 | 0.12181565 | -2.146353608 | 0.03387489 | -1.121667883 |
| 0.02052275 | 7.14238034 | 5.03E-11 | 4.402382982 | 2.35E-05 | 4.289450288 |
| 0.06183494 | -1.2468306 | 0.21460269 | 2.020672589 | 0.045558479 | 0.982801677 |
| 0.64443356 | 0.9413129 | 0.34821354 | -0.226017873 | 0.82157526 | 0.568273013 |
| 0.46436057 | 1.05952597 | 0.29123843 | 0.851312422 | 0.396305683 | 1.329323562 |
| 0.78534462 | 1.75311188 | 0.08183599 | -0.558795287 | 0.577351459 | 3.130301378 |
| 0.08381633 | -1.622913 | 0.1069238 | -1.21242664 | 0.227750684 | -2.142511795 |
| 0.67627795 | -0.9732103 | 0.3321766 | -0.83560071 | 0.40505417 | 0.798320606 |
| 0.2731005 | 1.85773197 | 0.06536956 | 0.209320576 | 0.834555904 | 1.077257075 |
| 0.74989179 | 0.04545576 | 0.96381072 | -2.057208252 | 0.041851409 | 1.464538194 |
| 0.18217252 | 6.23685361 | 5.27E-09 | 5.476258863 | 2.46E-07 | 4.216447733 |
| 0.75584838 | 3.3371082 | 0.00109195 | 2.065403735 | 0.041056306 | 3.564040147 |
| 0.00510455 | 5.94858722 | 2.17E-08 | 4.709695797 | 6.78E-06 | 4.04332264 |
| 0.03421551 | 1.82597902 | 0.0700469 | 4.697740479 | 7.12E-06 | 2.259154576 |
| 0.06970943 | -1.5895418 | 0.11426013 | -0.296429311 | 0.767418966 | 0.555233722 |
| 0.18523118 | 2.46097522 | 0.01510864 | 4.272673388 | 3.91E-05 | 3.407595551 |
| 0.00350864 | 3.26478552 | 0.001386 | 5.291676619 | 5.60E-07 | 2.218908698 |
| 0.14201655 | -0.0211769 | 0.98313556 | -1.576095016 | 0.117658713 | 1.096507209 |
| 0.03781858 | 4.32568433 | 2.92E-05 | 2.821467724 | 0.005603502 | 2.276681469 |
| 0.08023877 | 2.80975069 | 0.00568975 | 3.049042363 | 0.002829786 | 1.742896853 |
| 0.83362945 | 3.18977219 | 0.00176782 | 2.118994 | 0.036169956 | 2.674649121 |
| 0.50274157 | 3.74083293 | 0.00026951 | 4.987527159 | 2.11E-06 | 3.757013942 |
| 0.08758404 | 2.23688709 | 0.02692152 | 3.483866326 | 0.000692451 | 0.728745623 |
| 0.53939314 | 0.93652562 | 0.35066269 | 0.942696679 | 0.347745977 | 2.153102762 |
| 0.0028137 | 3.53800763 | 0.00055211 | 5.800191565 | 5.57E-08 | 3.562369411 |
| 0.4266839 | -1.1100129 | 0.26895227 | -0.847367749 | 0.398491198 | 1.090303466 |
| 0.45370314 | -2.0097391 | 0.04643719 | -3.628034506 | 0.000421955 | -1.525633091 |
| 0.71155967 | 1.38627477 | 0.16793155 | 0.481837202 | 0.63080657 | 1.60863875 |
| 0.16077785 | 2.03193071 | 0.04410799 | 0.839991948 | 0.402597375 | 1.735325519 |
| 0.0150692 | 5.13557814 | 9.54E-07 | 6.277087495 | 5.81E-09 | 3.943611457 |
| 0.0146983 | 0.23180674 | 0.81703655 | 0.261151222 | 0.794427315 | 0.917902446 |
| 0.7802785 | -0.6501314 | 0.51670408 | 2.351881219 | 0.020321554 | 1.369233801 |
| 0.98323439 | 1.16246069 | 0.2470842 | -0.325473477 | 0.745394329 | 1.591116967 |
| 0.48122348 | 0.23958682 | 0.8110113 | 0.346660767 | 0.729458427 | 0.47458021 |
| 0.00051948 | 4.97512649 | 1.94E-06 | 3.608825038 | 0.000451105 | 0.775008215 |
| 0.10197777 | 2.11323071 | 0.03640761 | 3.151237434 | 0.002057025 | 1.576903052 |
| 0.95654964 | 2.77014509 | 0.00638694 | 1.376971522 | 0.171105906 | 0.228126629 |

| | | | | | |
|---|---|---|---|---|---|
| 0.91962409 | 0.15108299 | 0.88013412 | 1.692225868 | 0.093219736 | 0.179734062 |
| 0.8432116 | 0.22451412 | 0.82269424 | 1.842830384 | 0.067842589 | 0.295539993 |
| 0.06065253 | -2.4632609 | 0.01501684 | -0.803820042 | 0.423103703 | 0.177047657 |
| 0.56637923 | 3.38099746 | 0.00094311 | 0.882707387 | 0.379174057 | 3.90125614 |
| 0.00697899 | -0.1713286 | 0.86422008 | -0.227514655 | 0.820414003 | 1.265296714 |
| 0.79918379 | 0.69048468 | 0.49106617 | 1.749212136 | 0.082832564 | 3.577669186 |
| 5.74E-05 | 6.81803189 | 2.75E-10 | 4.697063922 | 7.14E-06 | 5.333458331 |
| 0.40928559 | 4.95315805 | 2.13E-06 | 4.432870454 | 2.08E-05 | 2.678392037 |
| 0.36082213 | -0.8377347 | 0.40364953 | -0.940566284 | 0.348832532 | -0.127708379 |
| 0.0939926 | -2.9083603 | 0.00424488 | -2.523238343 | 0.0129468 | -0.907816491 |
| 0.97205409 | -1.4618824 | 0.1460803 | -3.505629368 | 0.000643136 | -0.541685053 |
| 0.83175135 | 3.14186493 | 0.00206061 | 2.317975606 | 0.022158575 | 3.348719823 |
| 0.16325192 | -0.7504783 | 0.45426312 | -1.706386168 | 0.09054461 | -1.428509989 |
| 0.04156939 | -0.1511996 | 0.8800423 | 1.495157024 | 0.137520674 | 2.406996874 |
| 0.31632834 | 1.44868883 | 0.14972622 | 0.788806756 | 0.431793652 | 0.118094064 |
| 0.04459618 | 5.25101478 | 5.69E-07 | 3.217726598 | 0.001664922 | 3.137237582 |

| Ab_7M_p |
|---|
| 0.268570863 |
| 0.029810921 |
| 0.739452118 |
| 0.021587821 |
| 0.56177416 |
| 0.981271367 |
| 0.897099537 |
| 0.323978579 |
| 0.166161217 |
| 0.628441908 |
| 0.238560823 |
| 0.275198122 |
| 0.140638815 |
| 0.154294117 |
| 0.628441908 |
| 0.077999527 |
| 0.276730892 |
| 0.762128087 |
| 0.826343263 |
| 0.823596564 |
| 0.038327522 |
| 0.018071606 |
| 0.127262986 |
| 0.251894632 |
| 0.422628398 |
| 0.769095815 |
| 0.04637825 |
| 0.112338213 |
| 0.483923853 |
| 0.681959334 |
| 0.231819712 |
| 0.539406893 |
| 0.769990711 |
| 0.03856034 |
| 0.436647381 |
| 0.521058485 |
| 0.068581662 |
| 0.704448509 |
| 0.822756752 |
| 0.704448509 |
| 0.109437271 |

| |
|---|
| 0.905145712 |
| 0.537513465 |
| 0.557478714 |
| 0.866292283 |
| 0.760926954 |
| 0.657322387 |
| 0.184603891 |
| 0.122891924 |
| 0.907473462 |
| 0.123187188 |
| 0.269501315 |
| 0.827657013 |
| 0.300071249 |
| 0.740402574 |
| 0.407019477 |
| 0.547528896 |
| 0.677047466 |
| 0.032914775 |
| 0.321370745 |
| 0.625721016 |
| 0.119175233 |
| 0.116597673 |
| 0.039326952 |
| 0.312694472 |
| 0.491795299 |
| 0.123491227 |
| 0.023133787 |
| 0.015790308 |
| 0.701440613 |
| 0.665372743 |
| 0.754793162 |
| 0.173194109 |
| 0.049592038 |
| 0.456804228 |
| 0.154565874 |
| 0.227010679 |
| 0.079466428 |
| 0.814125239 |
| 0.517397823 |
| 0.257684418 |
| 0.875223933 |
| 0.83202441 |

| |
|---|
| 0.77746656 |
| 0.1231477 |
| 0.206471592 |
| 0.078834327 |
| 0.514288147 |
| 0.649786803 |
| 0.248497538 |
| 0.317093852 |
| 0.079030847 |
| 0.889883296 |
| 0.983859855 |
| 0.833111135 |
| 0.013650515 |
| 0.180061518 |
| 0.065989521 |
| 0.437642211 |
| 0.08533039 |
| 0.337666503 |
| 0.884675819 |
| 0.023836061 |
| 0.191587193 |
| 0.880450454 |
| 0.149333239 |
| 0.79939526 |
| 0.728658845 |
| 0.883212635 |
| 0.846205311 |
| 0.149333239 |
| 0.587360581 |
| 0.067504662 |
| 0.599137732 |
| 0.556476759 |
| 0.618873703 |
| 0.633744035 |
| 0.193364355 |
| 0.000156153 |
| 0.867702338 |
| 0.066609026 |
| 0.444390452 |
| 0.374659544 |
| 0.444390452 |
| 0.093192794 |

| |
|---|
| 0.333309864 |
| 0.725269035 |
| 0.72867198 |
| 0.538451595 |
| 0.961348372 |
| 0.230536464 |
| 0.802312281 |
| 0.867154949 |
| 0.817291854 |
| 0.249919194 |
| 0.256366799 |
| 0.451051063 |
| 0.58224615 |
| 0.055049985 |
| 0.293493504 |
| 0.188339532 |
| 0.229088799 |
| 0.834146113 |
| 0.880843512 |
| 0.303968082 |
| 0.335247427 |
| 0.850913902 |
| 0.050775701 |
| 0.710398515 |
| 0.499546843 |
| 0.898353995 |
| 0.853227072 |
| 0.39538875 |
| 0.57838518 |
| 0.989562188 |
| 0.975384029 |
| 0.628744876 |
| 0.341921525 |
| 0.768253889 |
| 0.271171418 |
| 0.138282824 |
| 0.476053702 |
| 0.381481822 |
| 0.507876738 |
| 0.169292218 |
| 0.832215486 |
| 0.815517262 |

| |
|---|
| 0.152840873 |
| 0.049702867 |
| 0.052969249 |
| 0.048385882 |
| 0.387131302 |
| 0.445110588 |
| 0.875168935 |
| 0.317093852 |
| 0.550205872 |
| 0.993239806 |
| 0.732136407 |
| 0.818741032 |
| 0.198377112 |
| 0.323710321 |
| 0.971261652 |
| 0.802131849 |
| 0.131943164 |
| 0.092873334 |
| 0.644122274 |
| 0.134397138 |
| 0.349399134 |
| 0.048826981 |
| 0.310174266 |
| 0.589818291 |
| 0.683000359 |
| 0.846519509 |
| 0.512920962 |
| 0.99486242 |
| 0.709019624 |
| 0.891915432 |
| 0.346200391 |
| 0.998717126 |
| 0.809334332 |
| 0.80081405 |
| 0.21302622 |
| 0.826858716 |
| 0.498293739 |
| 0.055895319 |
| 0.804325428 |
| 0.287792706 |
| 0.042649482 |
| 0.634545311 |

| |
|---|
| 0.002167758 |
| 0.000307053 |
| 0.72408737 |
| 0.064214227 |
| 0.740283727 |
| 0.201665019 |
| 0.855350863 |
| 0.667260082 |
| 0.007620854 |
| 0.098919162 |
| 0.490876244 |
| 0.018799223 |
| 0.367920139 |
| 0.002451167 |
| 0.118234715 |
| 0.001186353 |
| 0.033468677 |
| 0.643491188 |
| 0.751940924 |
| 0.115956524 |
| 0.794644181 |
| 0.769129519 |
| 0.091137553 |
| 0.204013045 |
| 0.022821846 |
| 0.123937695 |
| 0.572870032 |
| 0.002363605 |
| 0.551484482 |
| 0.087831733 |
| 0.002956528 |
| 0.059608848 |
| 0.843165719 |
| 0.015192228 |
| 0.035338009 |
| 0.029641284 |
| 0.399586652 |
| 0.005275343 |
| 0.73118586 |
| 0.98852224 |
| 0.129164206 |
| 0.036078277 |

| |
|---|
| 0.009016114 |
| 0.342669786 |
| 0.510387364 |
| 0.306808871 |
| 0.001696803 |
| 0.003484325 |
| 0.652802175 |
| 0.229020739 |
| 0.099612428 |
| 0.006425567 |
| 0.090614829 |
| 0.404218487 |
| 0.004015449 |
| 0.800077979 |
| 0.154360928 |
| 0.824721381 |
| 0.062252635 |
| 0.018581093 |
| 0.232241735 |
| 0.010275257 |
| 0.762515257 |
| 0.056744471 |
| 0.000296154 |
| 0.041357792 |
| 0.417648171 |
| 0.09832923 |
| 0.381169967 |
| 0.130627669 |
| 0.111106825 |
| 0.002453865 |
| 0.258445814 |
| 0.095477955 |
| 0.699677421 |
| 0.360981122 |
| 0.703703369 |
| 0.417967815 |
| 0.022112634 |
| 0.232257108 |
| 0.375422733 |
| 0.171448567 |
| 0.815884745 |
| 0.604658645 |

| |
|---|
| 0.019320573 |
| 0.049647436 |
| 0.448143119 |
| 0.023620813 |
| 0.016876312 |
| 0.097233937 |
| 0.424156588 |
| 0.000742402 |
| 0.00110112 |
| 0.092950848 |
| 2.82E-05 |
| 0.399997133 |
| 0.526882044 |
| 0.992419027 |
| 0.004331444 |
| 0.885898609 |
| 0.264030121 |
| 0.076776998 |
| 0.035434826 |
| 0.003154752 |
| 0.66762735 |
| 0.144979122 |
| 0.009165611 |
| 0.790025361 |
| 0.002001697 |
| 0.89210547 |
| 0.533434292 |
| 0.001103755 |
| 0.598617337 |
| 0.002590324 |
| 0.025836191 |
| 0.496486357 |
| 0.456647468 |
| 0.957371621 |
| 0.213123178 |
| 0.094684197 |
| 0.00148927 |
| 2.17E-07 |
| 0.713348318 |
| 0.009758243 |
| 0.003852059 |
| 1.16E-05 |

| |
|---|
| 0.034000652 |
| 0.012432335 |
| 0.008377093 |
| 0.410008644 |
| 0.197085903 |
| 0.264922775 |
| 4.42E-05 |
| 0.328282605 |
| 0.571234049 |
| 0.187028277 |
| 0.00234054 |
| 0.034792478 |
| 0.42674042 |
| 0.284182069 |
| 0.146455848 |
| 5.80E-05 |
| 0.000581318 |
| 0.000109383 |
| 0.026235394 |
| 0.580082374 |
| 0.000973868 |
| 0.028952324 |
| 0.27571879 |
| 0.025123941 |
| 0.084692828 |
| 0.008853529 |
| 0.000301442 |
| 0.468008315 |
| 0.033926181 |
| 0.000584576 |
| 0.278426956 |
| 0.13053181 |
| 0.111121787 |
| 0.086031616 |
| 0.000156504 |
| 0.361070085 |
| 0.1742596 |
| 0.115013736 |
| 0.636210794 |
| 0.440322232 |
| 0.118250359 |
| 0.820054065 |

| |
|---|
| 0.85775663 |
| 0.7682474 |
| 0.859860152 |
| 0.000181925 |
| 0.208960285 |
| 0.000555374 |
| 6.86E-07 |
| 0.008762392 |
| 0.898658493 |
| 0.366347021 |
| 0.589344964 |
| 0.00117796 |
| 0.156531406 |
| 0.018081593 |
| 0.906250583 |
| 0.002291145 |
